# A scoping review of AI, speech and natural language processing methods for assessment of clinician-patient communication

**DOI:** 10.1101/2024.12.13.24318778

**Authors:** Pierre Albert, Brian McKinstry, Saturnino Luz

## Abstract

**Introduction:** There is growing research interest in applying Artificial Intelligence (AI) methods to medicine and healthcare. Analysis of communication in healthcare has become a target for AI research, particularly in the field of analysis of medical consultations, an area that so far has been dominated by manual rating using measures. This opens new perspectives for automation and large scale appraisal of clinicians’ communication skills. In this scoping review we summarised existing methods and systems for the assessment of patient doctor communication in consultations.

**Methods:** We searched EMBASE, MEDLINE/PubMed, the Cochrane Central Register of Controlled Trials, and the ACM digital library for papers describing methods or systems that employ artificial intelligence or speech and natural language processing (NLP) techniques with a view to automating the assessment of patient-clinician communication, in full or in part. The search covered three main concepts: dyadic communication, clinician-patient interaction, and systematic assessment.

**Results:** We found that while much work has been done which employs AI and machine learning methods in the analysis of patient-clinician communication in medical encounters, this evolving research field is uneven and presents significant challenges to researchers, developers and prospective users. Most of the studies reviewed focused on linguistic analysis of transcribed consultations. Research on non-verbal aspects of these encounters are fewer, and often hindered by lack of methodological standardisation. This is true especially of studies that investigate the effects of acoustic (paralinguistic) features of speech in communication but also affects studies of visual aspects of interaction (gestures, facial expressions, gaze, etc). We also found that most studies employed small data sets, often consisting of interactions with simulated patients (actors).

**Conclusions:** While our results point to promising opportunities for the use of AI, more work is needed for collecting larger, standardised, and more easily available data sets, as well as on better documentation and sharing of methods, protocols and code to improve reproducibility of research in this area.

## 1. Introduction

Clinician-patient communication has been the focus of considerable research efforts by the health community. The assessment of communication skills in medical consultations and teamwork among clinicians has been studied for more than sixty years, and numerous models of the medical consultation have been proposed. Better understanding of the patient’s motivations and expectations (attitude to illness, psychological aspects), and new insights on the sequence of the consultation itself have led to changes in practice, such as taking social history or safety netting during medical consultations, alongside the formalisation of phases and tasks of the consultation. This has led to the creation of guides and assessment tools for learning and training purposes, such as the Calgary Cambridge Guide to the Medical Interview [21].

Changes and discoveries in models naturally led to their integration in the training of health professionals. However, the assessment of doctors’ communication skills is a complex and time consuming process, performed by human experts. While innovations in automated processing have allowed an initial explorations of this domain, the development of automatic assessment of communication skills in clinical settings remains a challenging task.

Clinician-patient communication is a synchronous, usually dyadic communication: a dialogue between two participants interacting dynamically with each other, or tryadic — a clinician interacting with a patient and another person, such as the patient’s carer or a relative.

Sociological studies of the consultation have investigated many general traits of social behaviours of clinicians and patients. This includes the role of the patient, for instance, the definition and discussion of a patient’s “sick role” (normative expectations around illness) [36, 51], the relationship between clinician and patient [9], the influence of the general organisation of the healthcare system [50, 9], social aspects of health and disease [9], and social factors determining the health of individuals, groups, and large populations [9].

This general picture has been refined by actual observations of interaction patterns contrasted with patient expectations. Such patterns have been the subject of investigation over many years. Davis [11], for instance, analysed recordings of medical consultations combined with interviews and questionnaires to identify patterns of communication explaining non-compliance (tension between the patient and the clinician, lack of rapport, seeking information without giving feedback). Regarding patients’ expectations, McKinstry [29] (in a cross-sectional survey) and Elwyn et al. [14] found that patients varied in their desire for involvement in decision making, stressing the need for doctors to determine the level of involvement desired by a patient.

Communication in a dialogue can be divided into different modalities, verbal (speech), paralinguistic (tone, use of silence) and non-verbal (gestures, smiles, showing concern), and between *content*, i.e. the semantics of the interaction, and *content-free* aspects, the form of the interaction. The distinction between verbal and non-verbal aspects of an utterance refers to the distinction between the semantic content, and its paralinguistic content [16].

This review concerns technology that aims to extract meaning from patient-clinician interactions using existing tools, new methods, and their combination in a processing pipeline able to provide data that can be turned into metrics and feedback. While some work exists on the automatic assessment of parts of the communication, this domain is in its infancy and more is still needed for its practical applications, e.g. to teach and train communication skills. Nonetheless recent studies have demonstrated the capacity of current systems produce meaningful results, such as prediction of student’s success based on communication and domain skills [6], identification and assessment of suicidal risk using verbal and nonverbal cues during interviews with adolescents [52], characterisation of semantic similarity of the patient’s and physician’s language [53], etc.

At the acoustic level, speech processing focuses on content-free patterns that may be helpful in structuring the communication, such as *prosody* and *segmentation*. Prosody and assessment of voice quality have been used in clinical training using staged scenarios [55]. Spoken dialogue can be segmented by monitoring *turn taking patterns* or using *vocalisation patterns*. Vocalisation patterns [25, 24] are Markov diagrams encoding transition probabilities between vocalisation events of both participants, providing patterns of interaction. In medical applications, they have been used in the context of mental health to characterise power dynamics during dementia diagnosis disclosure conversations [45].

Semantic processing is the content-rich approach to speech processing. For the analysis of consultations, it first requires the transcription or automatic speech recognition (ASR), and its understanding using different semantic processing. A typical semantic pipeline includes diarisation (datermining who spoke when), ASR, syntactic analysys and semantic interpretation. Variations to this typical architecture presented in this review include the use of machine learning (ML) methods for detection of dialogue acts, analysis gestures, facial expressions and other non-verbal signals which affect communication [23].

## 2. Methods

The reporting of this scoping review follows the recommendations set by the preferred reporting items for systematic reviews and meta-analyses (PRISMA) statement [33].

### 2.1. Eligibility criteria

To be included all studies had to have three main characteristics. First, the studies had to analyse the interaction or communication between clinicians and patients (dyadic) in primary care settings. Second, the analyses had to be done by using automated methodologies including but not limited to machine learning and deep learning methodologies. Third, the studies had to report performance measure for these analyses.

#### 2.1.1. Dyadic clinician-patient interactions

Dyadic refers to the interaction between a clinician and one patient. Studies including a third person (e.g. carer or relatives) were not deemed eligible.

These conversations should occur only in primary care settings and the term clinician refers to any professional who provides care to patients in these settings such as general practitioners, and nurses.

The patient-clinician interaction had to occur in real time (synchronous) through spoken natural language, either face-to-face or remotely using videoconferencing technology. The interactions had to be spontaneous. We included in this category semi-structured interviews and studies that enrolled simulated patients.

#### 2.1.2. Automated analysis

Automated analysis include machine learning and deep learning methodologies that automatically extract features from dyadic communications. Other type of automated analysis are those that describe precise algorithms or specific instructions which needed to be followed to analyse the interaction.

#### 2.1.3. Performance measures

We distinguished three types of measures to assess performance of automated analyses of clinician-patient interactions: 1) intrinsic evaluations, such as F1-score, recall, precision, sensitivity, area under the receiver operating characteristic curve (AUC), 2) medical communication evaluation meeting certain criteria from medical frameworks, and 3) correlation with human assessment, such as the patient’s assessment, as reflected in questionnaires or structured interviews that yield a numerical score (e.g. [48, 13, 15].

### 2.2. Information sources

A systematic search was performed on Cochrane Central Register of Controlled Trials, Cochrane Database of Systematic Reviews, Embase and Medline through Pubmed from their inception to January, 2021. Because the importance of the Association for Computing Machinery (ACM) Digital Library as one of the world’s most comprehensive bibliographical databases in the field of computing we searched this digital library from inception to January 2021. We also included grey literature such as dissertation and theses as our information sources.

The search strategy included terms related to the following terms: 1) dyadic communication, 2) clinician-patient interaction, and 3)automated analysis. We also performed snowballing of included articles. We searched through the references of these articles and assess them against our eligibility criteria.

### 2.3. Article selection

All authors participated in abstract screening. Full-text screening was performed by one author (PA), with random samples assigned to SL and BM for confirmation of selection. A pilot for this last step was performed to homogenise the eligibility criteria of included studies. Borderline papers were identified regarding the interpretation of automation and patient-clinician consultations, and a stricter application of the criteria was advised. Following this, the definitions were clarified and every full text paper was reviewed a second time. Twenty-two studies were rejected and one additional study was included.

### 2.4. Data collection

We extracted the following overall information: 1) general characteristics such as publication date, first author, and location of the study, 2) baseline characteristics such as sample size, inclusion criteria, age, sex, ethnicity, socioeconomic information. We also extracted the following methodology-related data: 1) datasets’ characteristics: language, availability, annotated data, 2) the purpose of the interaction: palliative, risk-benefits of treatment options, etc. 3) the name of the framework or theory used to analyse the interaction, 4) type of input material: transcripts of video-recordings or audio-recordings, semi-structured interview transcripts, video-recordings, audio-recordings, manual annotation of non-linguistic features, etc, 5) features of interest: discourse acts.

In terms of results, we extracted the following information:

1. performance metrics: kappa, accuracy, sensitivity, specificity correlation scores, F-scores, AUC, and error scores;
2. dataset characteristics: language of the dataset, availability, type of collected information (transcription of video- or audio-recordings), input material for preprocessing or cleaning (transcripts, audio, video), preprocessing techniques (e.g. stop-word removal), type of system input information (video, text, or audio), and extracted features;
3. analysis characteristics: theory or framework behind the analysis, machine learning or statistical method (supervised, unsupervised, semisupervised), type of analysis (statistical, machine learning, deep learning).

### 2.5. Critical appraisal of included studies

The search was centred around the three main concepts of the review: dyadic communication, clinician-patient interaction, and systematic assessment.

The concepts grouped under *systematic assessment* of the communication by this review are diverse. Specific terms used in the language processing community are not always used by the medical community, and broader terms needed to be included. In addition, systematic assessment differs from automated assessment, encompassing studies that may have not used computational methods to extract features of the communication. Additional relevant terms were identified during a preliminary search on a subset of studies and reference lists. The final list (figure 1, item 1) includes terms from both medical and speech processing fields.

**Figure 1:**
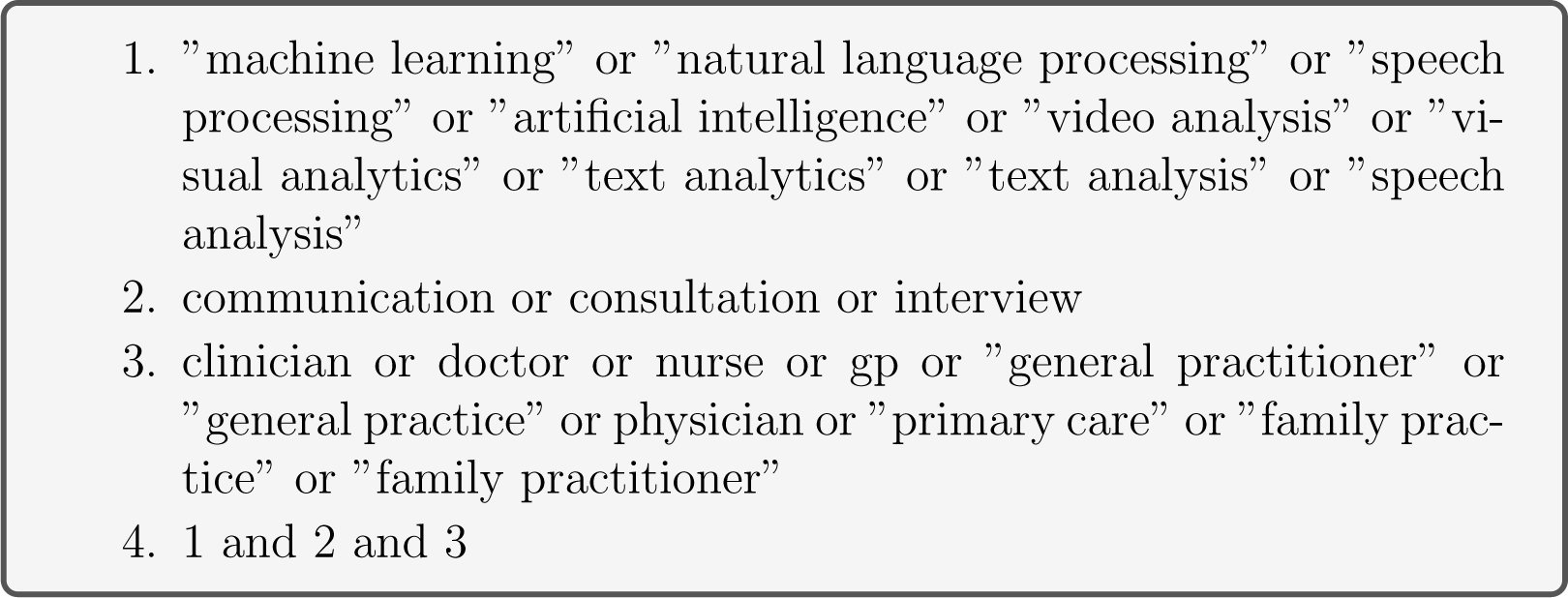
Search terms.

No previous review on a topic similar to this review was found during initial searches. The scoping and search strategy for this review was developed from scratch to identify studies using systematic approaches or automated processing to support the assessment of patient-clinician communication. Search updates were done as the search protocol was refined.

### 2.6. Sources searched

Searches were not restricted by location, date of study, or language. Grey literature (dissertations and theses) was also included for screening.

#### 2.6.1. Medical libraries

Systematic searches were performed in the main electronic databases, using the search strategy presented in figure 1.

Dates and issues of the medical databases searched for the review are:

**Cochrane** Cochrane Central Register of Controlled Trials Issue 1 of 12, January 2021 and Cochrane Database of Systematic Reviews Issue 1 of 12, January 2021. 44 reviews and 41 controlled trials were found.

**Embase** Embase 1980 to 2021 Week 03

**MEDLINE/PubMed** Accessed 2021-01-25

**ACM** Full-Text Collection was searched to retrieve studies with a strong focus set on the language processing aspect that may not have been reported in medical journals. The search was performed on full texts until 2019-08-30, then updated with searches on titles and abstracts until 2021-01-25.

Reference lists from eligible studies identified using the developed search strategy were searched manually for additional studies. Within-paper references were searched using Google Scholar^1^ or DuckDuckGo^2^ when not referenced to find further relevant studies. Search updates were conducted until January 2021, as mentioned in the respective online libraries search protocols.

### 2.7. Inclusion criteria

- Primary research study;
- The study is on clinician and/or patient communication;
- The studied interaction is based on synchronous interactive communication using spoken natural language (face-to-face or remotely), spontaneous (including semi-structured interviews), staged or not (e.g. a simulated patients acted a predefined scenario);
- Direct signals processing or their interpretation (e.g. speech or transcripts). Secondary interpretation, such as studies that extract patterns from manual annotations were included;
- Automated analysis is used. Therefore statistical analyses only based on manual annotations were discarded. Automation includes manual analysis in which a precise algorithmic methodology was described and used (following objective instructions, e.g. if … then … else …);
- Study must report evaluation measures. The measures can be classified into three types of evaluation: technical evaluation (e.g. standard NLP metrics), medical communication evaluation (e.g. using medical frameworks), and correlation with assessment (e.g. patient’s assessment).

Secondary sources were screened for the identification of additional material. Some studies in foreign language were included (French and German).

#### 2.7.1. Exclusion criteria

- Studies based on asynchronous communication: clinical narratives, medical notes (discharge summaries, nursing notes), speech notes using ASR;
- Automatic analysis of medical expert systems (diagnosis systems) and electronic health records without an interactive component;
- Studies using patient interviews or focus group discussions by researchers that were conducted after the interaction with clinicians for qualitative studies;
- Studies without a strong focus on communication between a clinician and a patient (e.g. team communication in presence of a patient);
- Studies reporting manual annotation and observation of the results whithout automation;
- Opinion and prospective papers;
- Studies with no full text available, or full text not in English, French or German.

### 2.8. Screening Procedure

A search of the main medical databases was conducted using the search strategy described in Figure 1. Results were automatically merged and duplicates removed using a specific tool^3^, then screened for relevance using the title, keywords, and abstracts. Relevance was established where studies discussed analysis of communication in a primary care setting or in a clinical setting similar to primary care (e.g. consultation with a surgeon). Full texts of identified studies were retrieved, and eligibility was screened against review inclusion and exclusion criteria outlined above.

#### 2.8.1. Updates

The search was updated four times — every 6 months — from July 2018 until January 2021. Retrieved results were merged and filtered, and previously screened references were discarded using the aforementioned automated tool. Potentially relevant studies uncovered during article screening (retrieved using Google Scholar) were also screened for eligibility.

#### 2.8.2. Results from the search

Due to the heterogeneity of the systems, aspects of communication, and interventions, a meta-analysis was not attempted. A detailed visualisation of the result of the search and screening procedure is provided in figure 2, formatted in accordance with the PRISMA flow diagram for screening [32]. We can group studies by themes according to the type of communication investigated. The first and largest group of studies explored *verbal communication*: the semantic content of the interaction. In this group, the first theme is the structures of the discourse, either task-specific [3] (VR-CoDES) or general [49] (behavioural codes), [52] (conversation dynamic), [4] (characterisation of utterances, sequential information), [47] (questions and answers), [27] (sequences of discourse elements). Related themes were task-based categories (interaction elements) [4] and the general structure of the dialogue (Speech acts) [56], [28]. The second theme focuses on topics what was discussed [57, 58, 35, 8, 10, 6]. The third theme relates to words: embeddings (use and context of a word) [39, 46, 10], types (e.g. part of speech) [28], frequency [47], and polarities (positive and negative words, e.g. related to gain and loss) [34, 17]. A final theme was the expression of affect (sentiment analysis) [47].

**Figure 2:**
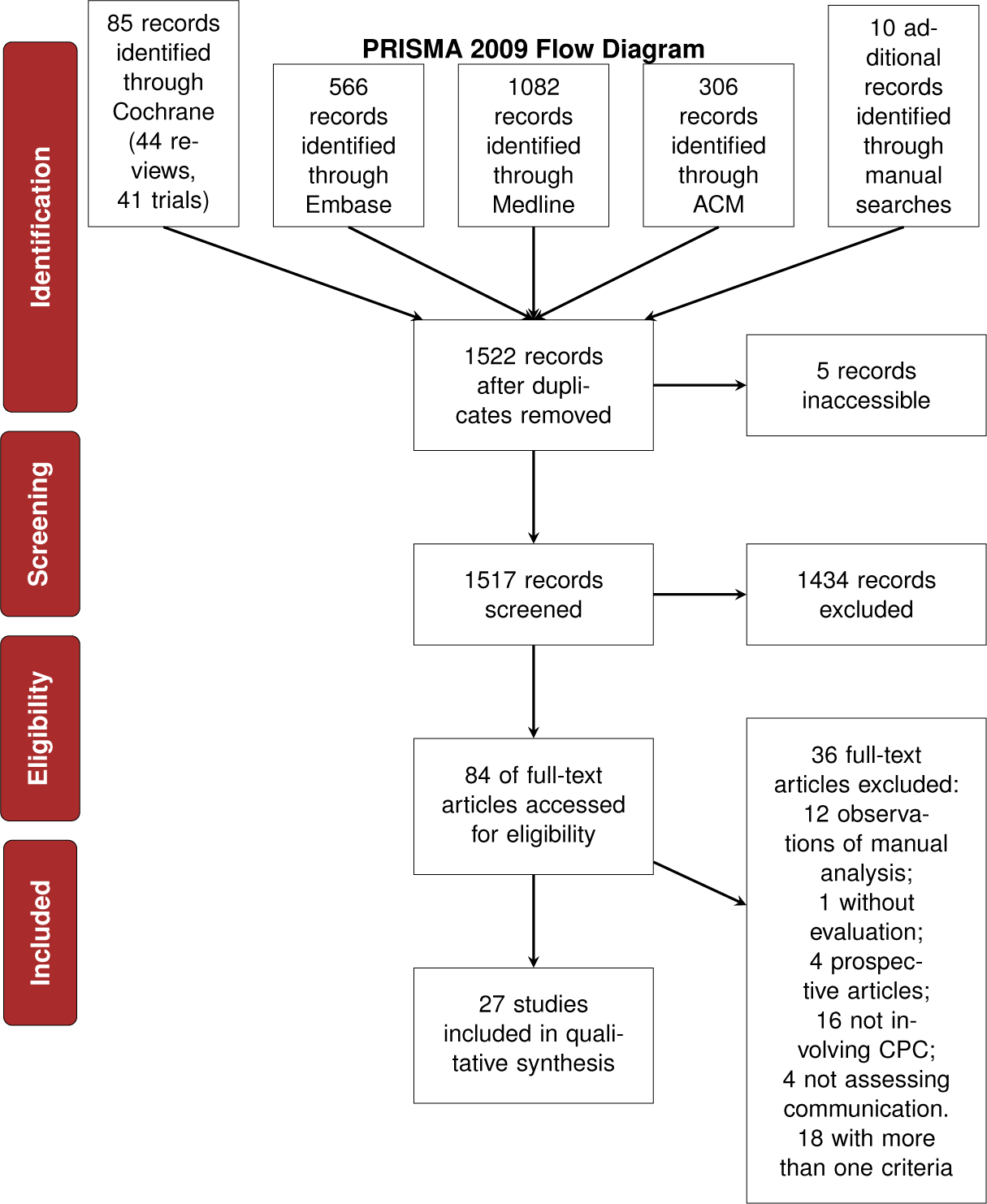
Detailed result screening procedure.

The other main group relates to the **non-verbal** components of the interaction: the part of the communication conveyed by other channels than the speech. Most use the visual modality: the face of participants [40, 39], gestures and movements [27, 18, 39], gaze [38, 39], and posture [7]. The other studies observed activities performed during the consultation: clinician’s activities [20] and computer / screen interactions [38].

The last group of investigation relates to the **paralinguistic** components: the part of the communication conveyed by the speech but not its content. Acoustic features (verbal dominance) [52] [46], pauses [26], and silence [12, 30, 26].

The type of interactions are shown in Table A.8. The *type of interaction* relates to the active participants in the interaction. Constrained analysis is specified when applying (e.g. only dyadic interactions are analysed). The value can be dyadic (2 persons) or triadic (3 persons). The *medical interaction* describes the context in which the clinician patient communication occurred, e.g. GP consultation, outpatient visits, etc.

Information of the eligible articles is summarised in two tables. First, studies following the “participants, interventions, comparisons and outcomes” (PICOS) framework [42] are shown in tables 1 and 2. Additionally, we included three columns: a brief description of the aim of the study, an outline of the methodology, and a summary of results.

**Table 1:**
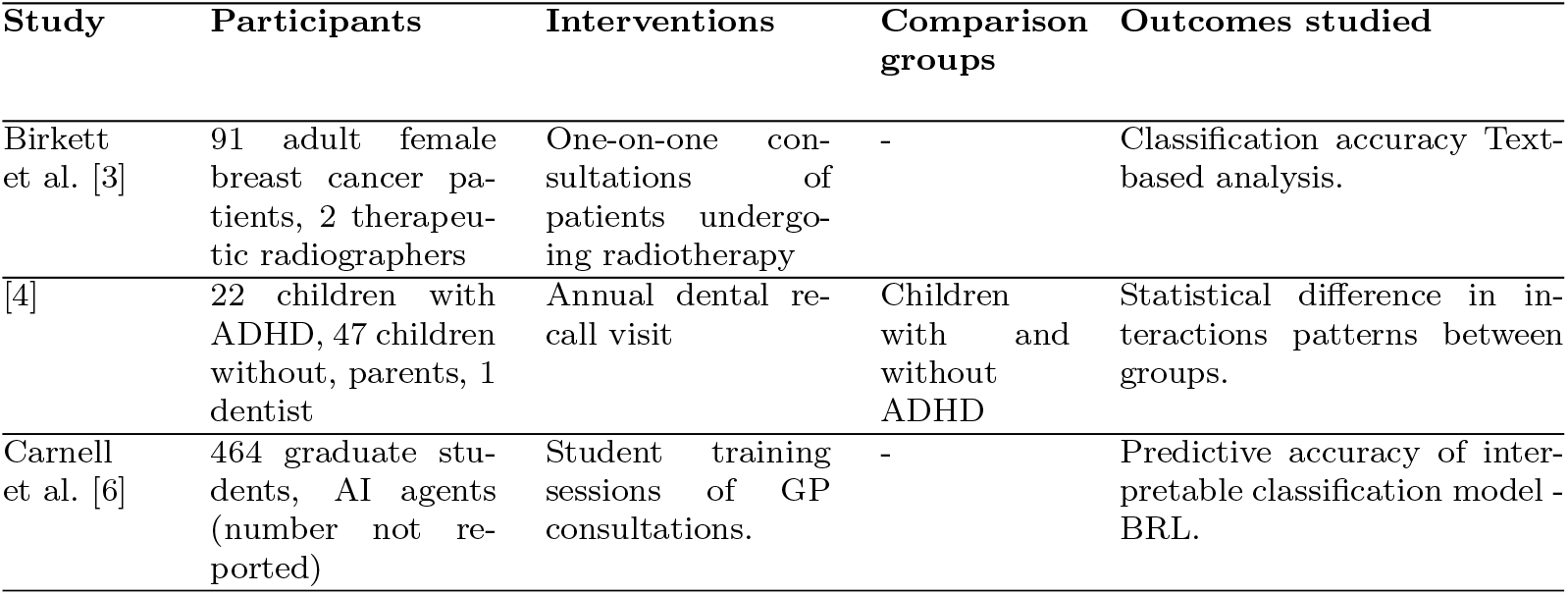
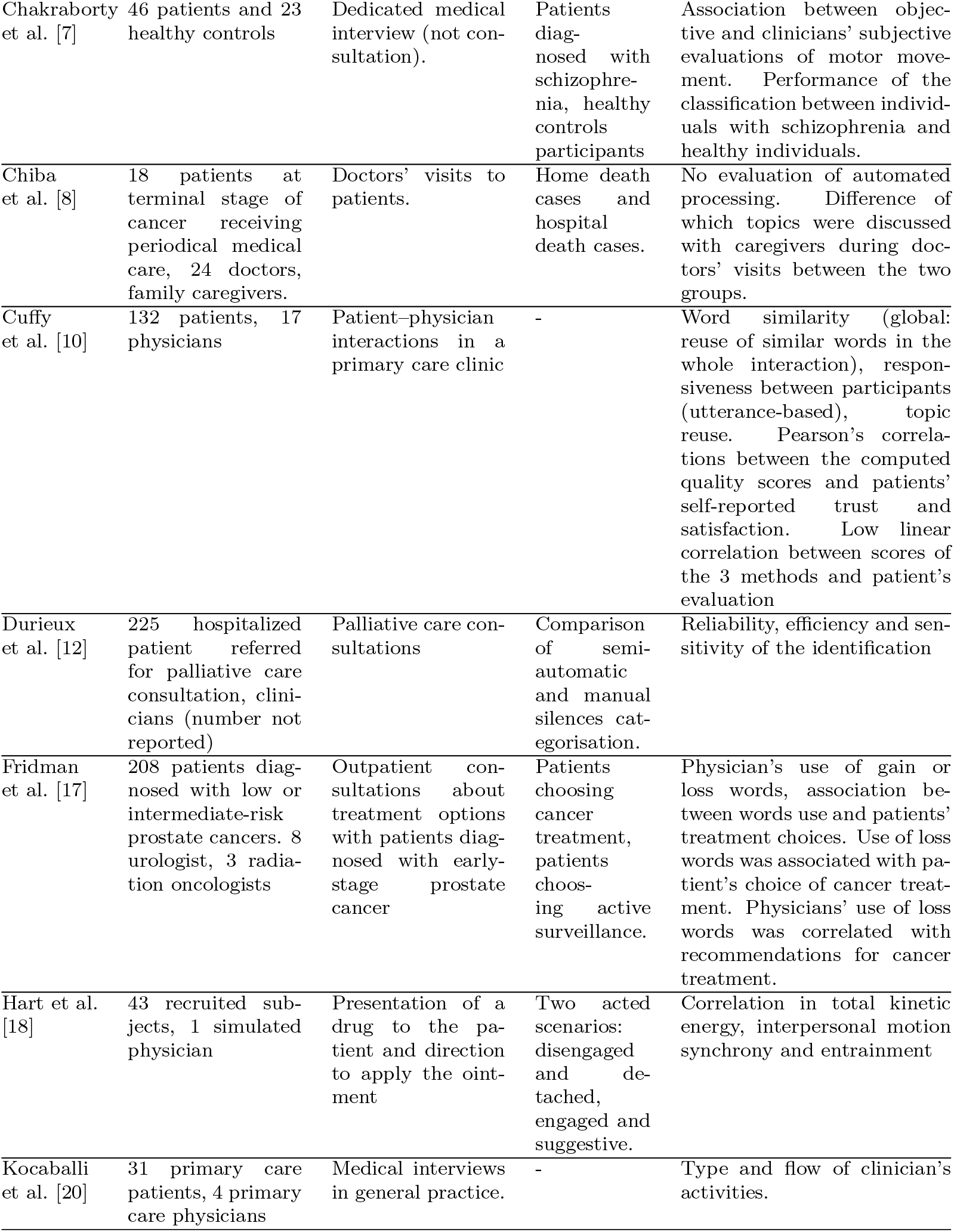
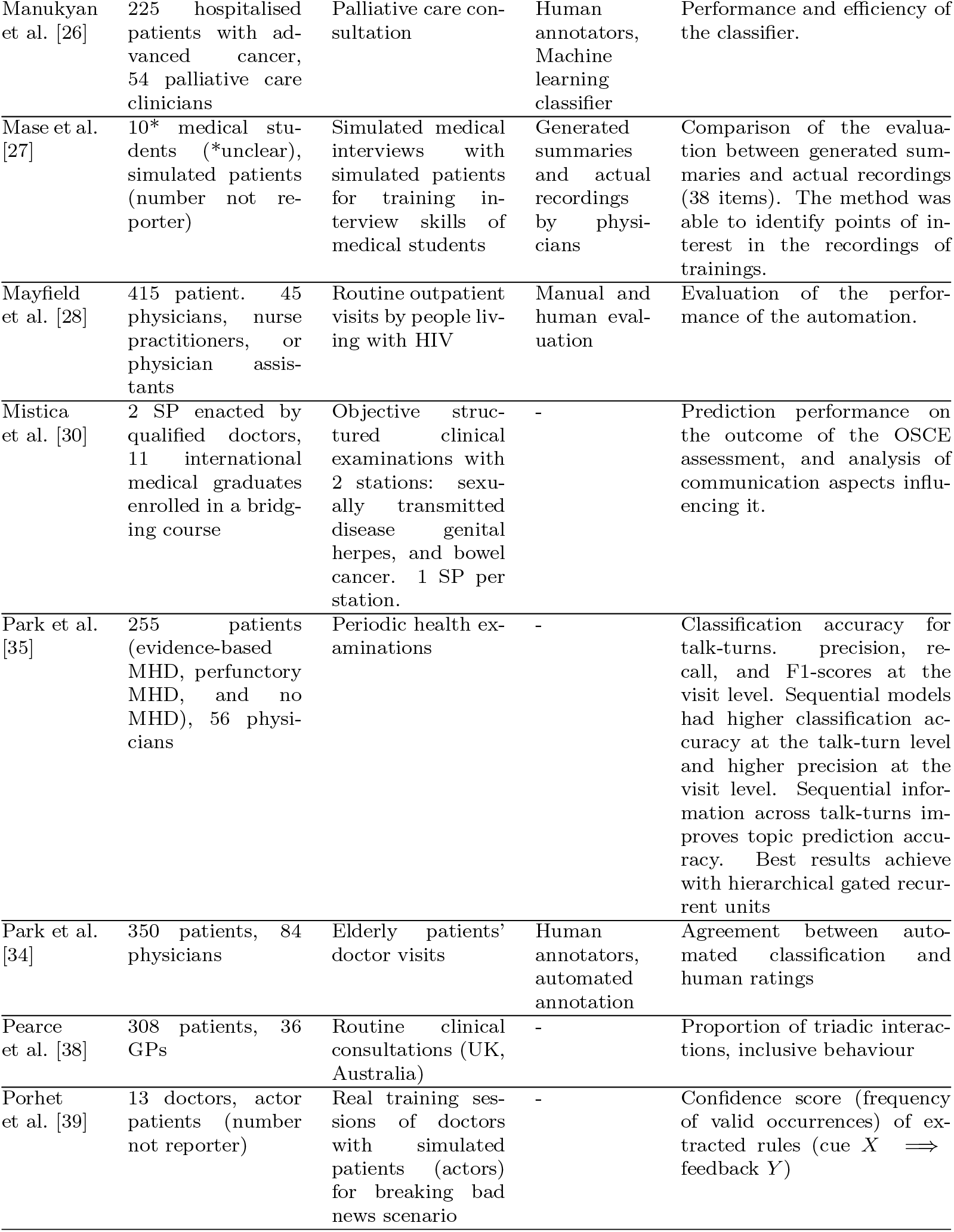
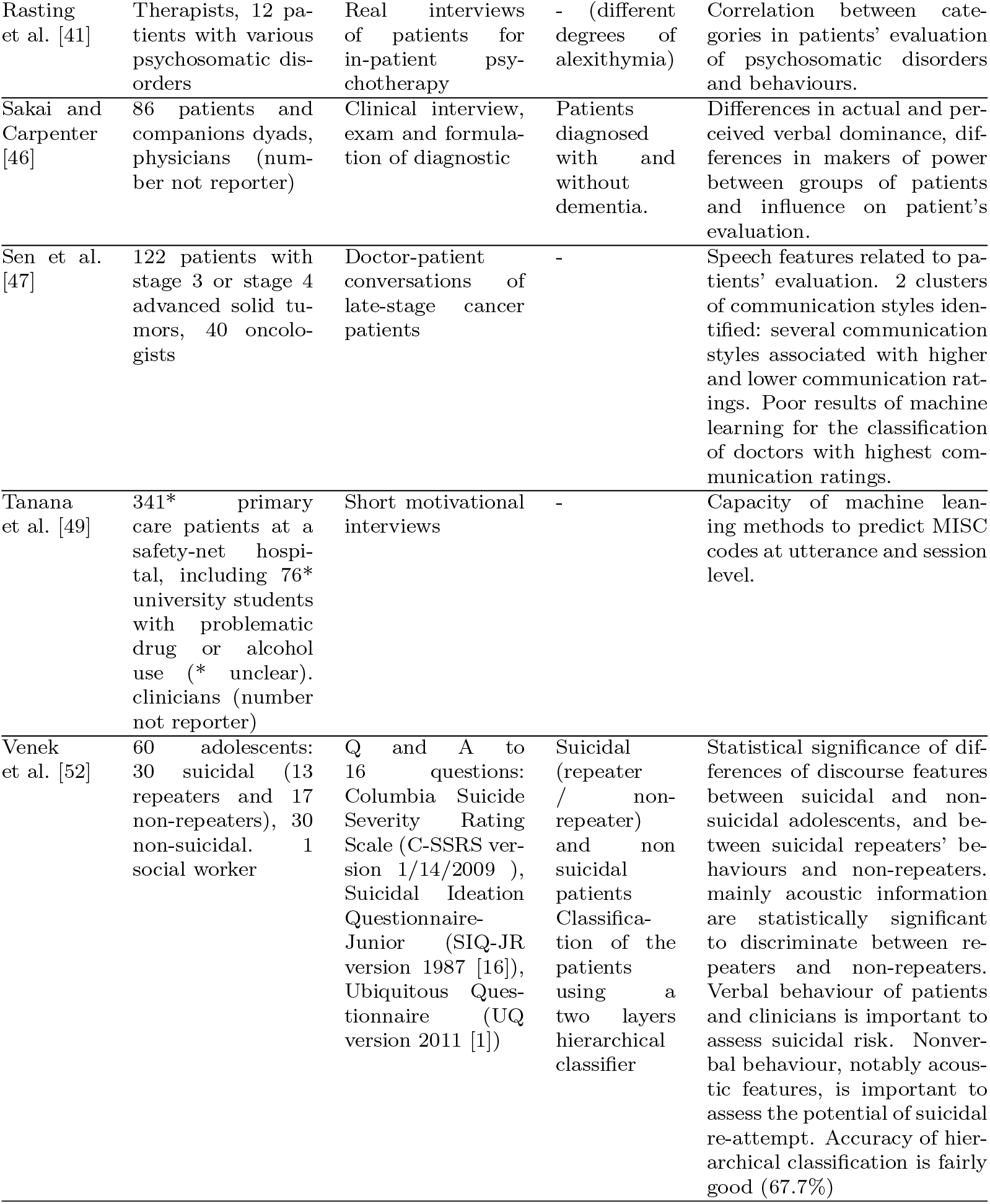
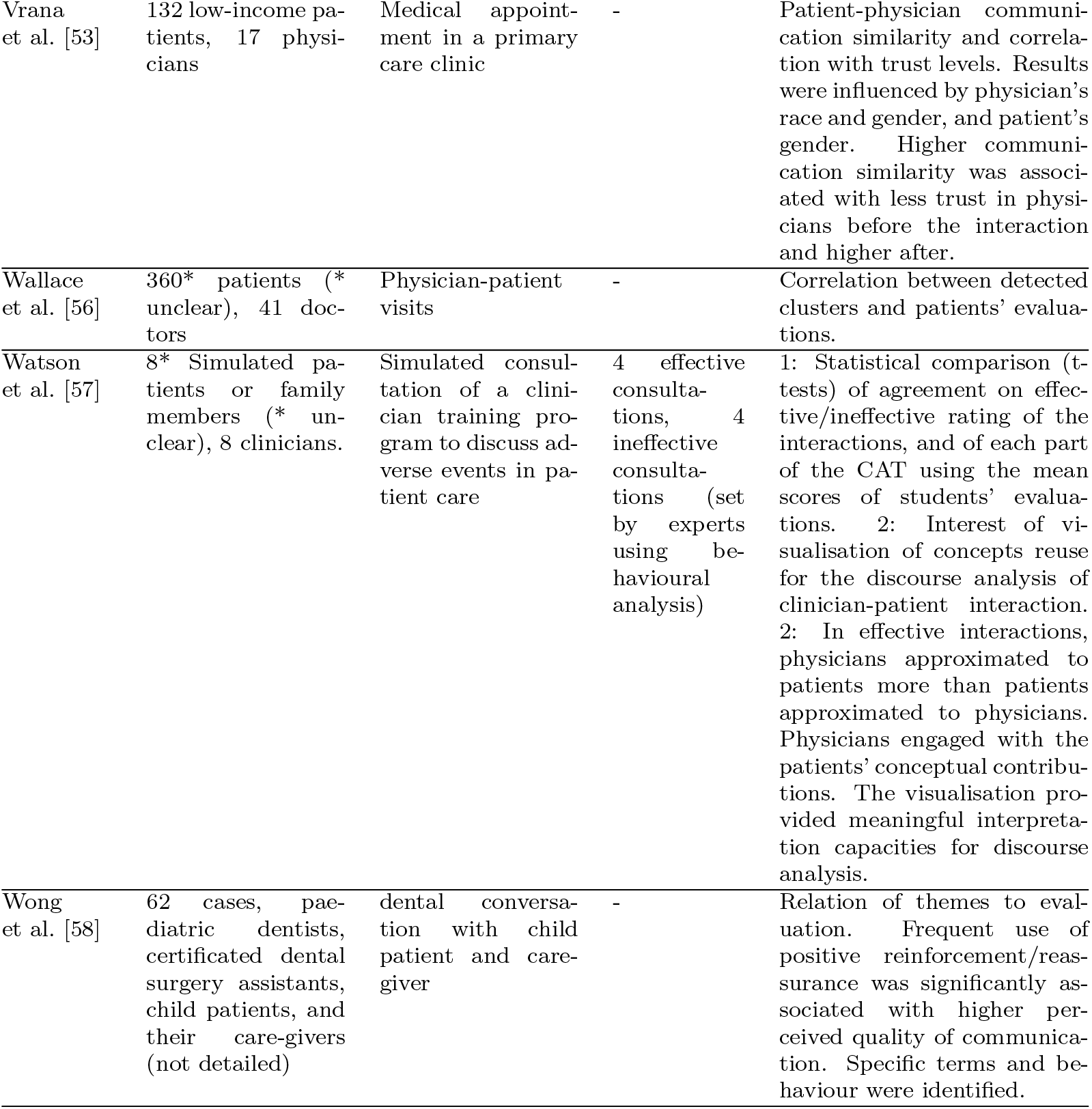
PICOS table. ADHD: Attention Deficit Hyperactivity Disorder, ANOVA: Analysis of variance, BRL: Bayesian Rule Lists, CAT: Communication Accommodation Theory, C-SSRS: Columbia Suicide Severity Rating Scale, kNN: k-Nearest Neighbours, LSA: Latent Semantic Analysis, MHD: Mental Health Discussion , MISC: Motivational Interviewing Skill Code, ML: Machine Learning, OSCE: Objective Structured Clinical Examination , PCA: Principal Component Analysis, RNN: Recurrent Neural Network, SIQ-JR: Suicidal Ideation Questionnaire-Junior, SP: Standardised Patients, SVM: Support Vector Machine, TFIDF: Term frequency-inverse document frequency, UQ: Ubiquitous Questionnaire

**Table 2:**
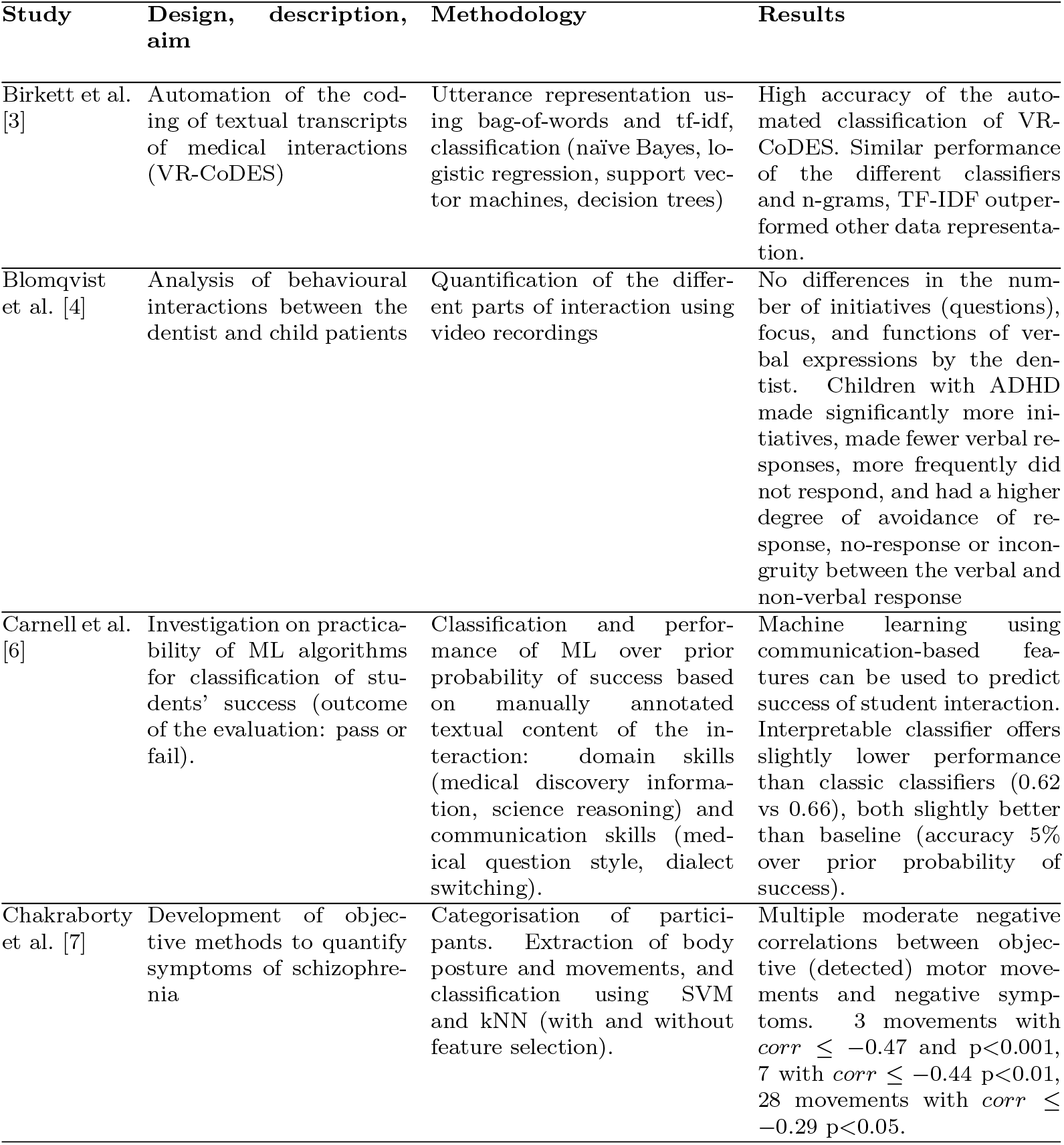
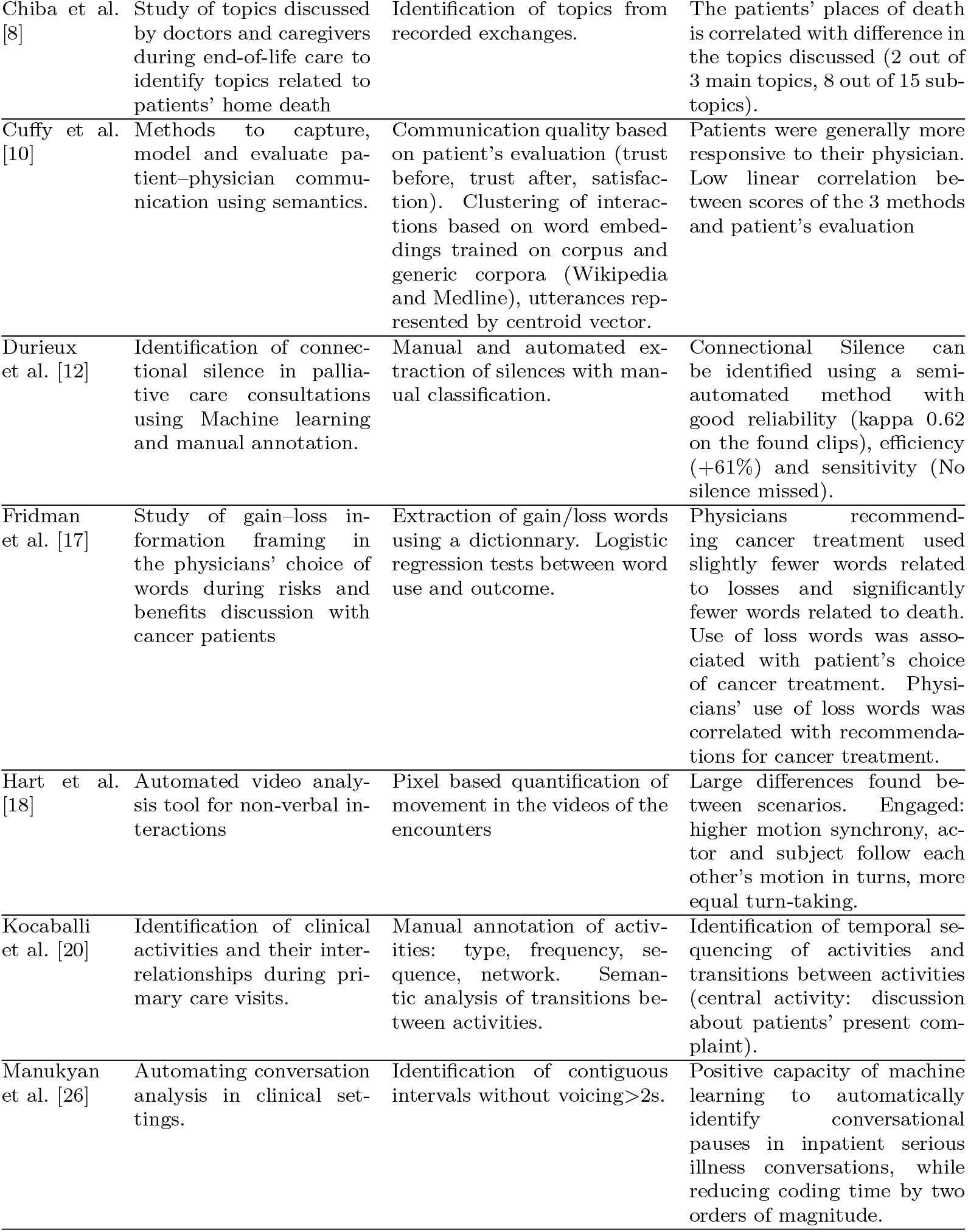
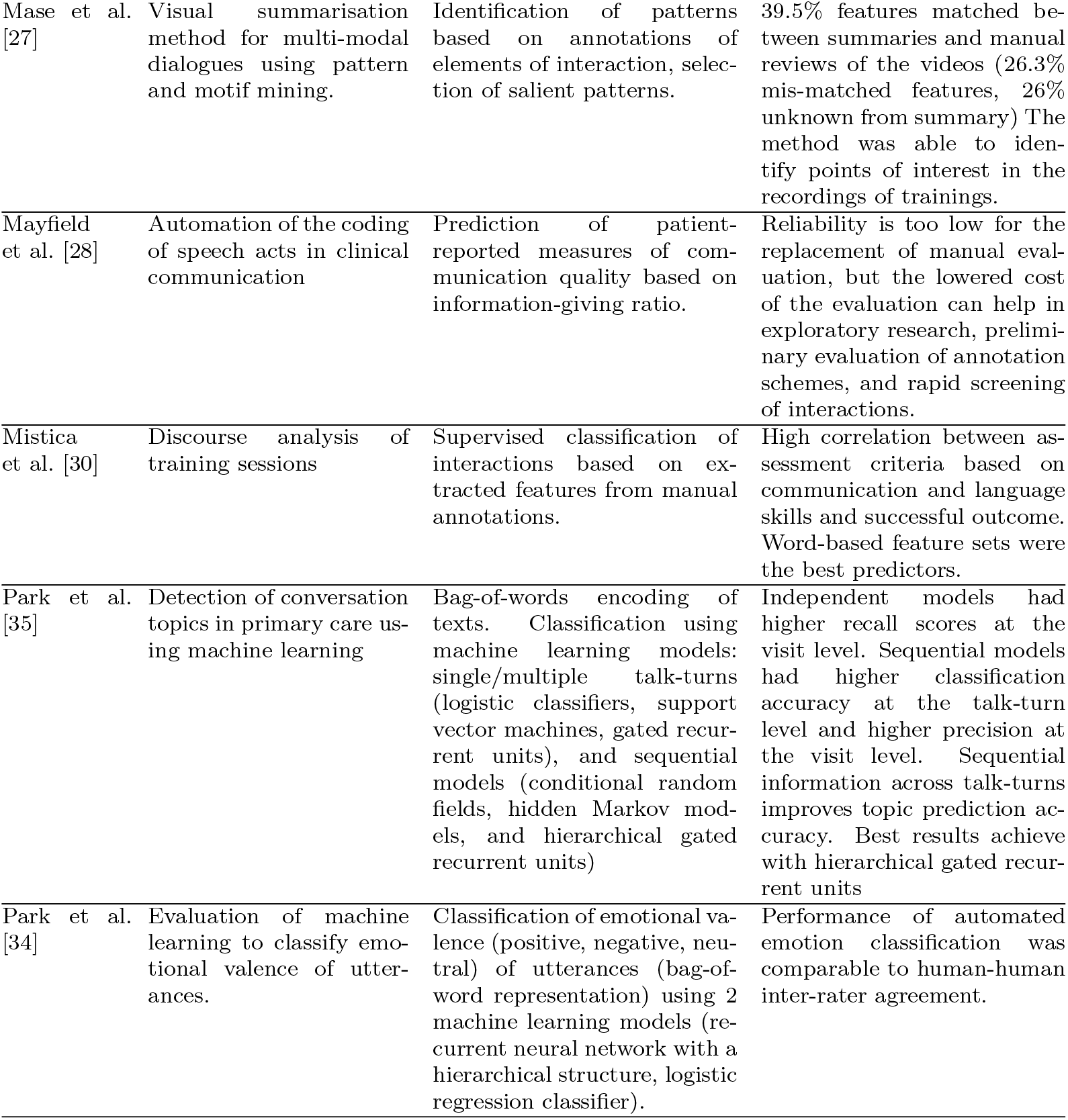
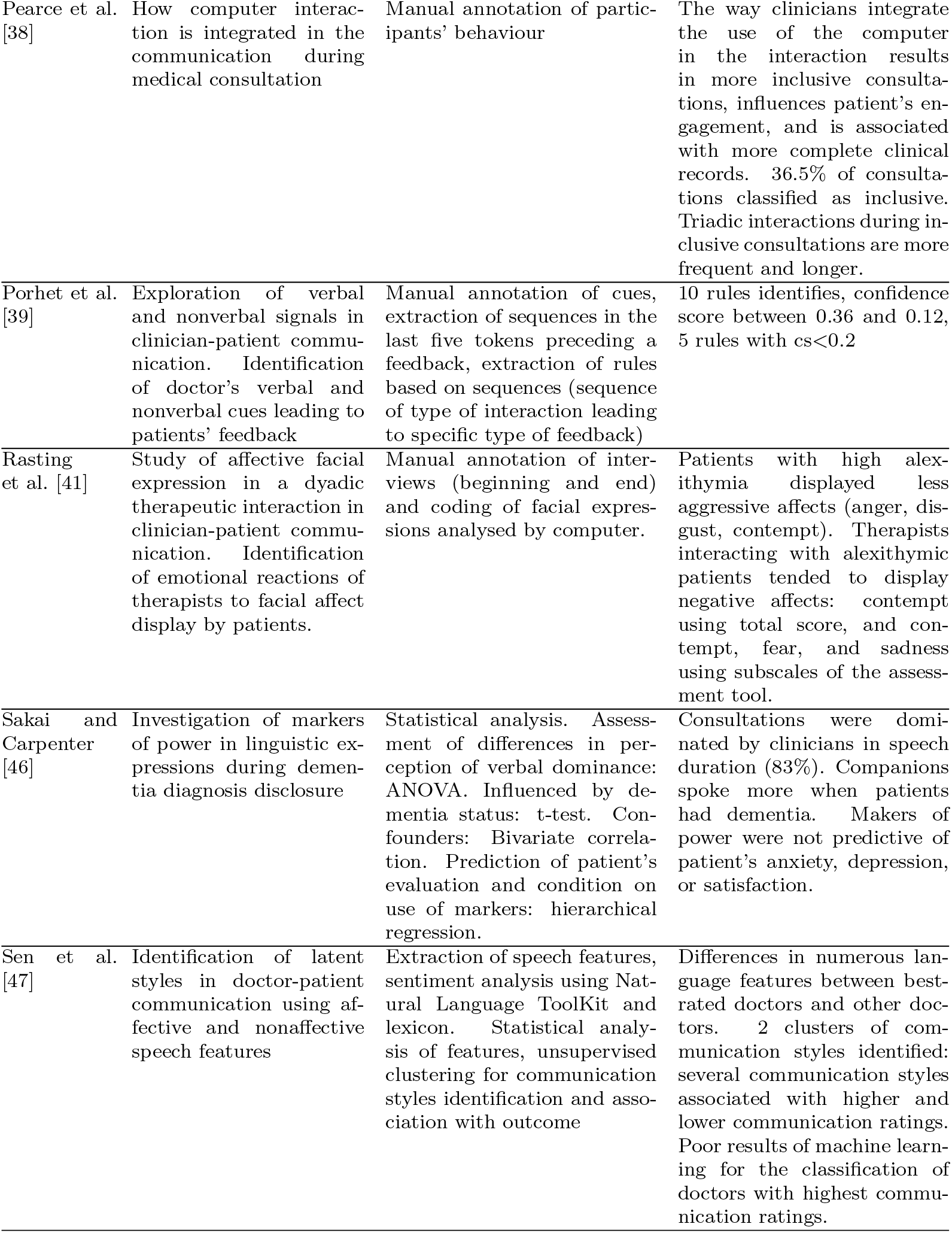
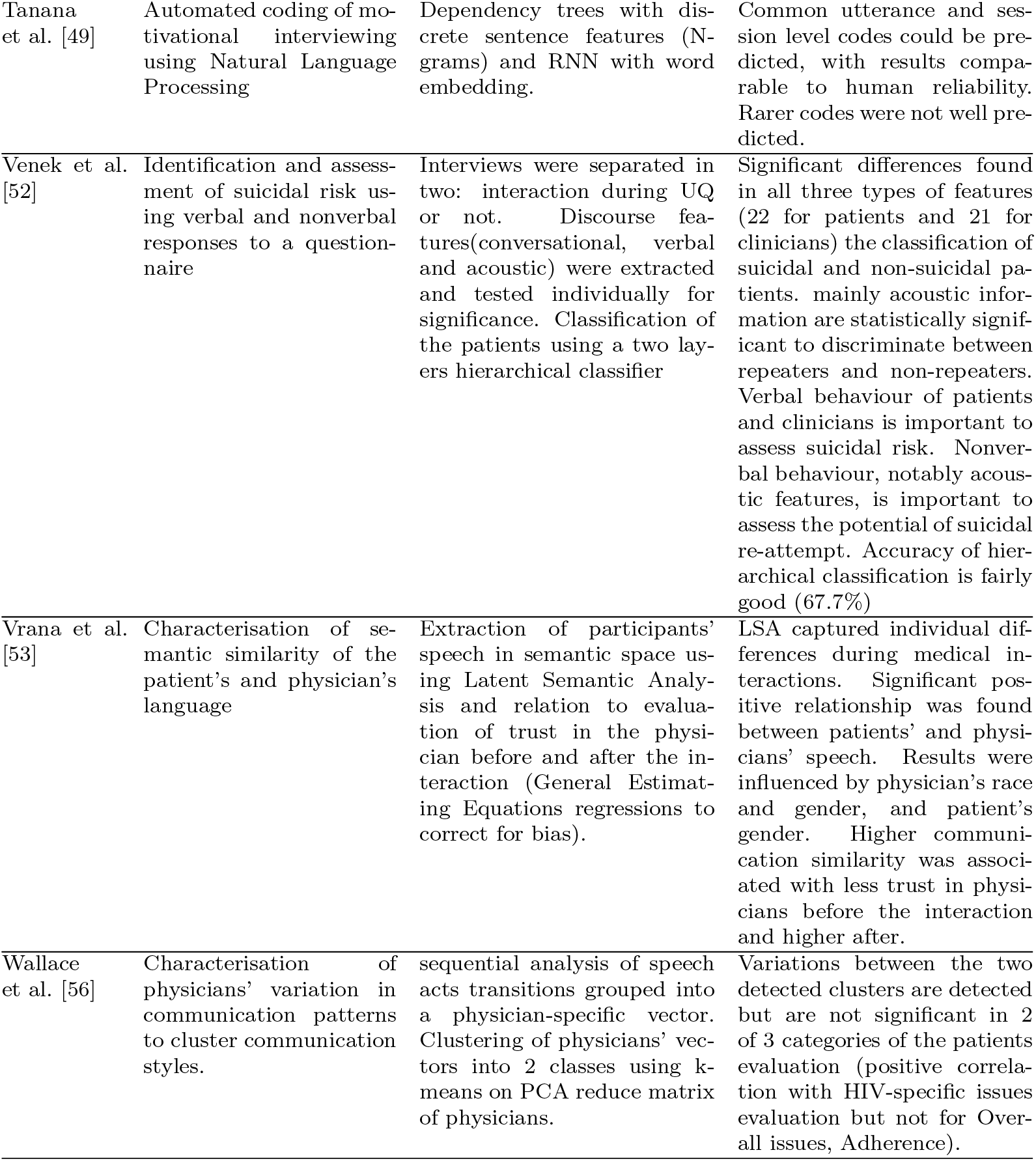
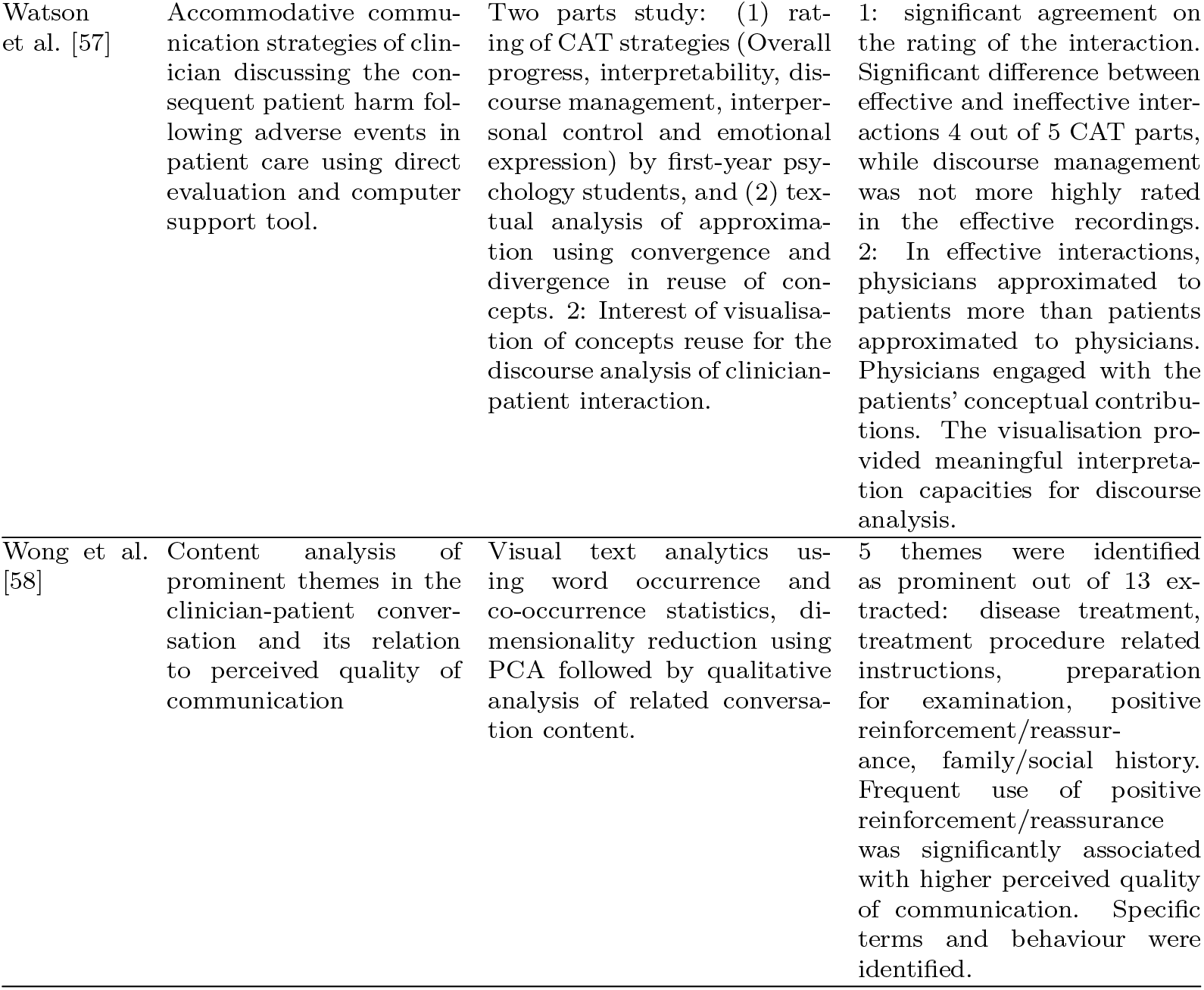
Design, methodoloy and results summary. ADHD: Attention Deficit Hyperactivity Disorder, ANOVA: Analysis of variance, BRL: Bayesian Rule Lists, CAT: Communication Accommodation Theory, C-SSRS: Columbia Suicide Severity Rating Scale, kNN: k-Nearest Neighbours, LSA: Latent Semantic Analysis, MHD: Mental Health Discussion , MISC: Motivational Interviewing Skill Code, ML: Machine Learning, OSCE: Objective Structured Clinical Examination , PCA: Principal Component Analysis, RNN: Recurrent Neural Network, SIQ-JR: Suicidal Ideation Questionnaire-Junior, SP: Standardised Patients, SVM: Support Vector Machine, TFIDF: Term frequency-inverse document frequency, UQ: Ubiquitous Questionnaire

| Study | Design, description, aim | Methodology | Results |
| --- | --- | --- | --- |
| Birkett et al. [3] | Automation of the coding of textual transcripts of medical interactions (VR-CoDES) | Utterance representation using bag-of-words and tf-idf, classification (naïve Bayes, logistic regression, support vector machines, decision trees) | High accuracy of the automated classification of VR-CoDES. Similar performance of the different classifiers and n-grams, TF-IDF outperformed other data representation. |
| Blomqvist et al. [4] | Analysis of behavioural interactions between the dentist and child patients | Quantification of the different parts of interaction using video recordings | No differences in the number of initiatives (questions), focus, and functions of verbal expressions by the dentist. Children with ADHD made significantly more initiatives, made fewer verbal responses, more frequently did not respond, and had a higher degree of avoidance of response, no-response or incongruity between the verbal and non-verbal response |
| Carnell et al. [6] | Investigation on practicality of ML algorithms for classification of students' success (outcome of the evaluation: pass or fail). | Classification and performance of ML over prior probability of success based on manually annotated textual content of the interaction: domain skills (medical discovery information, science reasoning) and communication skills (medical question style, dialect switching). | Machine learning using communication-based features can be used to predict success of student interaction. Interpretable classifier offers slightly lower performance than classic classifiers (0.62 vs 0.66), both slightly better than baseline (accuracy 5% over prior probability of success). |
| Chakraborty et al. [7] | Development of objective methods to quantify symptoms of schizophrenia | Categorisation of participants. Extraction of body posture and movements, and classification using SVM and kNN (with and without feature selection). | Multiple moderate negative correlations between objective (detected) motor movements and negative symptoms. 3 movements with $corr \leq -0.47$ and $p < 0.001$, 7 with $corr \leq -0.44$ $p < 0.01$, 28 movements with $corr \leq -0.29$ $p < 0.05$. |
| Chiba et al. [8] | Study of topics discussed by doctors and caregivers during end-of-life care to identify topics related to patients' home death | Identification of topics from recorded exchanges. | The patients' places of death is correlated with difference in the topics discussed (2 out of 3 main topics, 8 out of 15 sub-topics). |
| Cuffy et al. [10] | Methods to capture, model and evaluate patient-physician communication using semantics. | Communication quality based on patient's evaluation (trust before, trust after, satisfaction). Clustering of interactions based on word embeddings trained on corpus and generic corpora (Wikipedia and Medline), utterances represented by centroid vector. | Patients were generally more responsive to their physician. Low linear correlation between scores of the 3 methods and patient's evaluation |
| Durieux et al. [12] | Identification of connectional silence in palliative care consultations using Machine learning and manual annotation. | Manual and automated extraction of silences with manual classification. | Connectional Silence can be identified using a semi-automated method with good reliability (kappa 0.62 on the found clips), efficiency (+61%) and sensitivity (No silence missed). |
| Fridman et al. [17] | Study of gain-loss information framing in the physicians' choice of words during risks and benefits discussion with cancer patients | Extraction of gain/loss words using a dictionary. Logistic regression tests between word use and outcome. | Physicians recommending cancer treatment used slightly fewer words related to losses and significantly fewer words related to death. Use of loss words was associated with patient's choice of cancer treatment. Physicians' use of loss words was correlated with recommendations for cancer treatment. |
| Hart et al. [18] | Automated video analysis tool for non-verbal interactions | Pixel based quantification of movement in the videos of the encounters | Large differences found between scenarios. Engaged: higher motion synchrony, actor and subject follow each other's motion in turns, more equal turn-taking. |
| Kocaballi et al. [20] | Identification of clinical activities and their inter-relationships during primary care visits. | Manual annotation of activities: type, frequency, sequence, network. Semantic analysis of transitions between activities. | Identification of temporal sequencing of activities and transitions between activities (central activity: discussion about patients' present complaint). |
| Manukyan et al. [26] | Automating conversation analysis in clinical settings. | Identification of contiguous intervals without voicing > 2s. | Positive capacity of machine learning to automatically identify conversational pauses in inpatient serious illness conversations, while reducing coding time by two orders of magnitude. |
| Mase et al. [27] | Visual summarisation method for multi-modal dialogues using pattern and motif mining. | Identification of patterns based on annotations of elements of interaction, selection of salient patterns. | 39.5% features matched between summaries and manual reviews of the videos (26.3% mis-matched features, 26% unknown from summary) The method was able to identify points of interest in the recordings of trainings. |
| Mayfield et al. [28] | Automation of the coding of speech acts in clinical communication | Prediction of patient-reported measures of communication quality based on information-giving ratio. | Reliability is too low for the replacement of manual evaluation, but the lowered cost of the evaluation can help in exploratory research, preliminary evaluation of annotation schemes, and rapid screening of interactions. |
| Mistica et al. [30] | Discourse analysis of training sessions | Supervised classification of interactions based on extracted features from manual annotations. | High correlation between assessment criteria based on communication and language skills and successful outcome. Word-based feature sets were the best predictors. |
| Park et al. [35] | Detection of conversation topics in primary care using machine learning | Bag-of-words encoding of texts. Classification using machine learning models: single/multiple talk-turns (logistic classifiers, support vector machines, gated recurrent units), and sequential models (conditional random fields, hidden Markov models, and hierarchical gated recurrent units) | Independent models had higher recall scores at the visit level. Sequential models had higher classification accuracy at the talk-turn level and higher precision at the visit level. Sequential information across talk-turns improves topic prediction accuracy. Best results achieve with hierarchical gated recurrent units |
| Park et al. [34] | Evaluation of machine learning to classify emotional valence of utterances. | Classification of emotional valence (positive, negative, neutral) of utterances (bag-of-word representation) using 2 machine learning models (recurrent neural network with a hierarchical structure, logistic regression classifier). | Performance of automated emotion classification was comparable to human-human inter-rater agreement. |
| Pearce et al. [38] | How computer interaction is integrated in the communication during medical consultation | Manual annotation of participants' behaviour | The way clinicians integrate the use of the computer in the interaction results in more inclusive consultations, influences patient's engagement, and is associated with more complete clinical records. 36.5% of consultations classified as inclusive. Triadic interactions during inclusive consultations are more frequent and longer. |
| Porhet et al. [39] | Exploration of verbal and nonverbal signals in clinician-patient communication. Identification of doctor's verbal and nonverbal cues leading to patients' feedback | Manual annotation of cues, extraction of sequences in the last five tokens preceding a feedback, extraction of rules based on sequences (sequence of type of interaction leading to specific type of feedback) | 10 rules identifies, confidence score between 0.36 and 0.12, 5 rules with $cs < 0.2$ |
| Rasting et al. [41] | Study of affective facial expression in a dyadic therapeutic interaction in clinician-patient communication. Identification of emotional reactions of therapists to facial affect display by patients. | Manual annotation of interviews (beginning and end) and coding of facial expressions analysed by computer. | Patients with high alexithymia displayed less aggressive affects (anger, disgust, contempt). Therapists interacting with alexithymic patients tended to display negative affects: contempt using total score, and contempt, fear, and sadness using subscales of the assessment tool. |
| Sakai and Carpenter [46] | Investigation of markers of power in linguistic expressions during dementia diagnosis disclosure | Statistical analysis. Assessment of differences in perception of verbal dominance: ANOVA. Influenced by dementia status: t-test. Confounders: Bivariate correlation. Prediction of patient's evaluation and condition on use of markers: hierarchical regression. | Consultations were dominated by clinicians in speech duration (83%). Companions spoke more when patients had dementia. Makers of power were not predictive of patient's anxiety, depression, or satisfaction. |
| Sen et al. [47] | Identification of latent styles in doctor-patient communication using affective and nonaffective speech features | Extraction of speech features, sentiment analysis using Natural Language ToolKit and lexicon. Statistical analysis of features, unsupervised clustering for communication styles identification and association with outcome | Differences in numerous language features between best-rated doctors and other doctors. 2 clusters of communication styles identified: several communication styles associated with higher and lower communication ratings. Poor results of machine learning for the classification of doctors with highest communication ratings. |
| Tanana et al. [49] | Automated coding of motivational interviewing using Natural Language Processing | Dependency trees with discrete sentence features (N-grams) and RNN with word embedding. | Common utterance and session level codes could be predicted, with results comparable to human reliability. Rarer codes were not well predicted. |
| Venek et al. [52] | Identification and assessment of suicidal risk using verbal and nonverbal responses to a questionnaire | Interviews were separated in two: interaction during UQ or not. Discourse features(conversational, verbal and acoustic) were extracted and tested individually for significance. Classification of the patients using a two layers hierarchical classifier | Significant differences found in all three types of features (22 for patients and 21 for clinicians) the classification of suicidal and non-suicidal patients. mainly acoustic information are statistically significant to discriminate between repeaters and non-repeaters. Verbal behaviour of patients and clinicians is important to assess suicidal risk. Nonverbal behaviour, notably acoustic features, is important to assess the potential of suicidal re-attempt. Accuracy of hierarchical classification is fairly good (67.7%) |
| Vrana et al. [53] | Characterisation of semantic similarity of the patient's and physician's language | Extraction of participants' speech in semantic space using Latent Semantic Analysis and relation to evaluation of trust in the physician before and after the interaction (General Estimating Equations regressions to correct for bias). | LSA captured individual differences during medical interactions. Significant positive relationship was found between patients' and physicians' speech. Results were influenced by physician's race and gender, and patient's gender. Higher communication similarity was associated with less trust in physicians before the interaction and higher after. |
| Wallace et al. [56] | Characterisation of physicians' variation in communication patterns to cluster communication styles. | sequential analysis of speech acts transitions grouped into a physician-specific vector. Clustering of physicians' vectors into 2 classes using k-means on PCA reduce matrix of physicians. | Variations between the two detected clusters are detected but are not significant in 2 of 3 categories of the patients evaluation (positive correlation with HIV-specific issues evaluation but not for Overall issues, Adherence). |
| Watson et al. [57] | Accommodative communication strategies of clinician discussing the consequent patient harm following adverse events in patient care using direct evaluation and computer support tool. | Two parts study: (1) rating of CAT strategies (Overall progress, interpretability, discourse management, interpersonal control and emotional expression) by first-year psychology students, and (2) textual analysis of approximation using convergence and divergence in reuse of concepts. 2: Interest of visualisation of concepts reuse for the discourse analysis of clinician-patient interaction. | 1: significant agreement on the rating of the interaction. Significant difference between effective and ineffective interactions 4 out of 5 CAT parts, while discourse management was not more highly rated in the effective recordings. 2: In effective interactions, physicians approximated to patients more than patients approximated to physicians. Physicians engaged with the patients' conceptual contributions. The visualisation provided meaningful interpretation capacities for discourse analysis. |
| Wong et al. [58] | Content analysis of prominent themes in the clinician-patient conversation and its relation to perceived quality of communication | Visual text analytics using word occurrence and co-occurrence statistics, dimensionality reduction using PCA followed by qualitative analysis of related conversation content. | 5 themes were identified as prominent out of 13 extracted: disease treatment, treatment procedure related instructions, preparation for examination, positive reinforcement/reassurance, family/social history. Frequent use of positive reinforcement/reassurance was significantly associated with higher perceived quality of communication. Specific terms and behaviour were identified. |

A summary of tasks related to clinician-patient communication assessment performed in each study is provided in table A.5. The *frameworks* column contains the medical and/or annotation that were used or referenced, the *type of material* is the type of data on which the study was conducted (e.g. audio, video). The *task performed* lists the processing applied to the data, either manual and automated (e.g. emotion recognition). The *performance* variable summarises the main quantified results, and the *dataset* variable describes the collected data. The information is then developed using six tables to extract detailed information relevant to this review (available in appendix, see section Appendix A).

The population of each study is described in two tables: table A.6 for the patients, and table A.7 for the clinicians. Both tables include the same demographic information in addition to the population included in the study: age, sex, ethnicity, location and socio-economical information. Patient-specific information relates to the personal, socioeconomic attributes, and medical condition of the cohort. Clinician-specific information regards speciality and experience. The analysis conducted in each study is then detailed in table

A.9, which contains the following columns:

- *Preprocessing* list the procedures undertaken on raw data (text, audio, video) as preparation steps for subsequent extraction of features analysis. Text processing usually include transcription, in which case the method is reported (by professionals or by researchers). Since no instance of the use of ASR was found, all reported transcriptions were manually produced. It must be stressed however that instances of uses of ASR to help generate transcripts were found: Alloatti et al. [1] for instance used manually corrected ASR output on 30 physiotherapy sessions. Other text preprocessing methods include cleaning of transcripts and removal of unwanted events, such as stop-words or disfluency. preprocessing of audio and video can include segmentation, extraction of parts (beginning, end), signal processing (background noise removal, normalisation, colour balance etc.). Finally, any manual processing is also listed.
- *Feature extraction* reports on automated processing. The generation of the features was documented either from raw data (acoustic or video analysis) or manually generated data (through text analysis: tokenisation, part-of-speech tagging, etc.)
- *Task and method* reports on the task that was performed (e.g. classification of a sentence) and on the methodology that was used. This includes supervised learning or unsupervised learning, a detail of any analysis used used: machine learning algorithms, clustering, feature set reduction, classification, etc. An accompanying table of abbreviations is provided in section A.3.
- *Evaluation* reports how the results were assessed in the study. This is broken down in four items, if present in the study — B: baseline, PM: performance metric, CV: cross-validation technique used, T: test set held out and its size.
- *Results* are numerical results of the reported performance metrics.

An assessment of the research potential and applications of each of the study is presented in table A.10. It is structured around the following columns:

- *Research implications* regroups three general characteristics. Novelty (yes/no): whether the study implemented a new method or applied an existing one — no is assigned where the study uses an existing tool or method. Replicability (low/partial/full): whether the reported procedure is described in sufficient details and data is available — low is assigned where both data is n4ot available and method description is incomplete; partial where either is the case, and full where both data and detailed methods are available. Generalisability (low/medium/high): whether the analysis is specific to the task — low is assigned where the method can only be applied to similar settings; medium where the analysis can be applied to other settings (i.e. type of medical encounters) with adaptations (e.g. changing a dictionary of terms); high where the analysis can be applied directly to other settings.
- *Risk of bias* Real life (RL, yes/partial/no): whether the interaction featured real interactions (e.g. between patients and doctors) or simulated interactions (e.g. training sessions with an actor). Feature balance (FB, yes/no): whether reported individual features were balanced across classes. Suitable metrics (SM, yes/no): whether metrics other than overall accuracy are reported when data are class-imbalanced. Contextualised results (CR, yes/no): whether a baseline is provided to put the results into perspective. Overfitting (yes/no): whether crossvalidation and/or hold-out set were used. Sample size (S): three ranges are reported: ≤ 50, ≤ 100, and ≥ 100).
- *Strengths/Limitations* five characteristics are reported with yes/partial/no assessment, each yes indicating a strength, each no indicating a limitation. Spontaneous speech: whether speech was naturally generated or prompted in response to open-answer questions. Conversational speech: whether the study is based dialogue. Automation: whether the automation (other than the machine learning tasks) was complete (excluding preprocessing) or only some aspects of the procedure used in the study. Transcription-free: whether the method required transcription of the dialogue. Content-independence: whether the method is content-based or not.

Finally the dataset table (table A.11) summarises details of the datasets used in the reviewed studies. It contains the following columns:

- *Data set/Subset size* Quantification of the number of documents details by groups of participants, including number of minutes recorded and number of words when available.
- *Data type* Data recorded and used in the study. Two types of data are reported. Data streams (audio, video) and derived data (e.g. transcripts — with information about the transcription when available). Other type of data (patients’ information, questionnaires, etc.). The type of interaction during the dialogue is characterised as either structured, semi-structured, or conversational.
- *Data annotation* Type of annotations with details about the annotation set.
- *Data balance* reports whether the dataset is balanced in terms of age (a), gender (g), and socio-professional class (s). Yes reports balance for both between and within class balance when applicable.
- *Data availability* whether the dataset has been published or made available.
- *Language* is the spoken language used during the interactions. It can differ from the main language of the country where the collection took place.

## 3. Results and discussion

Before analysing their content, a look at the distribution of the dates of publications of the included articles (see figure 3) provides a sense of how recently the field has emerged. All studies were published after 2005, and more than half of the articles were published after 2018.

**Figure 3:**
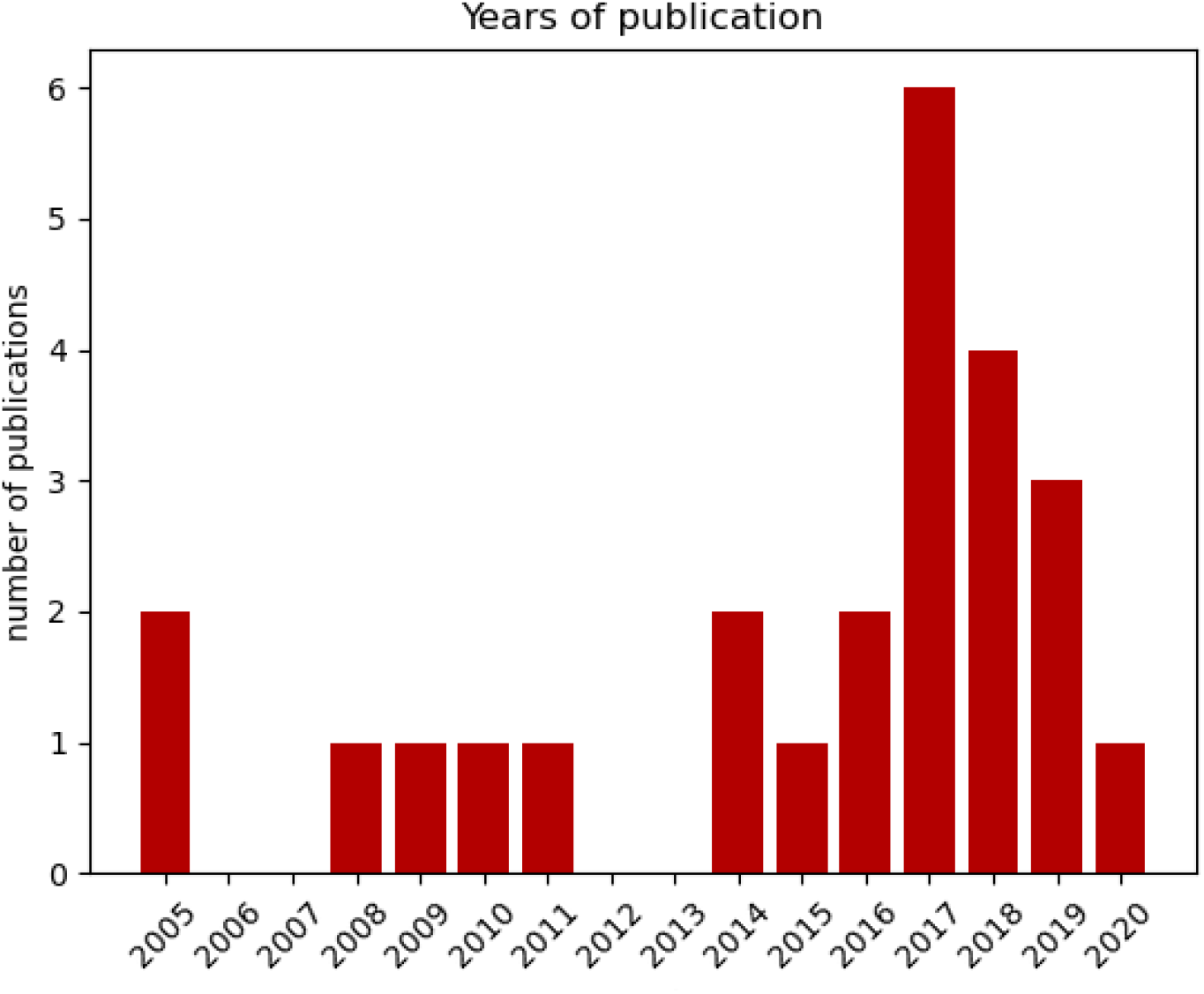
Distribution of the years of publication of included studies.

A total of 27 studies are included in the final selection. While they cover a wide range of aspects of clinician-patient communication, with only a limited number of studies having been dedicated to each aspect. A wide range of medical speciality are featured: General Practice, dentistry, radiography, language pathology, psychometry, oncology, urology, palliative care, psychotherapy, home medical care. In five occurrences, the interacting clinicians were medical students. A single study used an actor to perform the role of the doctor [18], in order to control the behaviour in preset scenarios (engaged or disengaged).

The retrieved studies feature several types of clinician-patient interactions. Twenty-two studies were conducted on real interactions and five were simulated, including one with a virtual avatar. The dialogues during medical interactions can be grouped in three different type. Twenty studies are based on conversational interactions, i.e. free form interactions during which the participants exchange freely without constraints over the content. Five studies used semi-structured interviews, i.e. an open discussion with a set of themes or questions to direct the interaction or elicit answers. Finally two studies used structured interactions, i.e. a planned, constrained discussion during which the same set of predefined questions is asked to each participant.

Regarding settings, 17 studies investigated medical consultations, either during GP consultations or routine patient visits (e.g. dental care), of which 13 were dyadic consultations (clinician and patient) and three were triadic interactions (a patient’s helper or a second clinician). One study features mainly dyadic interactions with a small fraction of triads. Overall, 7 studies report triadic interactions. A majority of the triads concerns an additional caregiver (e.g. parent). Only one feature an active second clinician although a few report non-interactive clinicians (passive, observing) or interacting before or after the studied interaction.

Five studies used clinical interviews (intake interviews, diagnoses or assessment of a particular condition). Two featured motivational interviewing (one on substance use, one on adherence dialogues). Two investigated disclosure interactions and the breaking bad news. Finally, one used a instruction session (on how to use a specific drug).

Of the 17 studies investigating medical consultations, seven used constrained topics and one investigated only a specific phase of the interaction. The cultural context of the studies (see table A.6) was fairly restricted. More than half of the studies (fifteen) were conducted in the USA, and all but five were conducted in western countries (USA, UK, Scotland, Australia, France, Germany). Of the others, none were conducted in developing countries: four were conducted in Asian countries (Japan, Singapore (PRC), Hong Kong) and one in Israel. The socio-cultural diversity was also quite lacking. Reported age and sex were generally balanced (featuring patients of all age, from children to elderly people). The distribution of ethnicity seemed balanced when reported, but the information is missing in more than half of the studies. While some patients of lower income or lower education were included in some studies, with two studies specifically on low-income cohorts, the information is also often lacking.

Most of the studies investigated cohorts of patients with cancer (seven studies) or patients for general consultations (five). Five studies were conducted with patients suffering from psychological issues such as suicidal thoughts or patients with Dementia, which could potentially influence their speech.

Regarding clinicians (see table A.7), most studies do not report information beyond sex distribution, and even this is missing for eighteen of the studies. Out of the studies reporting those, sex distribution was equal, which can simply signal that studies which paid attention to this metric paid attention to the sex distribution while recruiting the cohort. This is further illustrated regarding the ethnicity, where out of the four studies reporting it, only one was featuring a cohort of white only clinicians. Interactions featured a wide range of clinicians: nurses, oncologists, GPs, etc. Five studies were conducted with students clinicians, and two with resident doctors.

### 3.1. Investigated aspects

Most studies, 20 out of 27, investigated the semantics of the interaction. Ten studies used the global semantic space (such as topics in Carnell et al. [6] or participants’ semantic space in Vrana et al. [53]), i.e. the spoken content of the participants as a whole to characterise the communication.

Nine studies investigated topics or closely related concepts in conversations [57, 58, 35, 8, 10], either investigating consequences of differences in their presence or frequency, or evaluating internal structures, e.g. tracking reuse by participants. One additional study addressed the use and presence of more specific task-based categories: Blomqvist et al. [4] investigated interaction elements, characterising syntactic roles of utterances (statement/information, question, request).

Word-based studies are another type of unstructured characterisation, based on the quantification of used words: [39, 46, 10], the words used and their context (word embeddings), their type (part of speech) [28], or their frequency [47]. Investigations of emotions using verbal features were undertaken with two objectives: the classification of positive or negative speech using word polarities [34, 17], and the detection of sentiments from text [47]. While 15 studies investigated unstructured content (e.g. occurrences of topics), five studies investigated the discourse structure of the interaction, tracking the use, presence and absence of predefined sets of structuring elements: either task-specific structure in Birkett et al. [3]) based on VRCoDES [59], a system for coding the patient’s expressions of emotional distress, or using a more general linguistic approach (using behavioural codes in Tanana et al. [49], or a set of conversation dynamic features in Venek et al. [52]).

Other studies interpreted the interaction in a more global way by investigating the structure of the interaction, identifying links between its elements and their sequences: Sen et al. [47] tracked the questions and answers between participants, and Mase et al. [27] extracted patterns of interaction from sequences of discourse elements. Blomqvist et al. [4] combined the characterisation of utterances (syntax and type) with sequential information: source (spoke, focus) what was the aim of the utterance, response.

Some studies used concepts stemming directly from theoretical linguistics, such as speech acts [56, 28], although the precise definition of what constitutes a speech act varies across studies. Although both Mayfield et al. [28] and Wallace et al. [56] defined the speech act as a social act embodied in an utterance, and both restricted the possible acts to the categories listed by the Generalized Medical Interaction System (GMIAS) [22], Mayfield et al. [28] aggregated multiple categories into two acts: information-giving and information-requesting.

Further paralinguistic analysis uses acoustic features for the characterisation of speech, generally for its classification, e.g. between healthy and unhealthy patients, [52], but also for investigating non-verbal aspects of the interaction such as the types of pauses [26]. Another paralinguistic aspect of the interaction is the characterisation of the sequences of spoken interaction, or speakers turns: silences [12, 30] and verbal dominance [46] (calculated indirectly by quantifying the words of each participants).

Manukyan et al. [26] and Durieux et al. [12] are based on the same parent cohort study. Their experiments were conducted by the same team and complement each other: speech and silence detection, characterisation of the silences. Manukyan et al. [26] extracted and aggregated of acoustic features for the identification of conversational pauses. The random forest classifier achieved slightly lower accuracy than manual annotators (94.4% vs 99.1% over a ground truth defined as the consensus of three human coders) but it was much faster than the human coders (two orders of magnitude, requiring minutes instead of hours). Durieux et al. [12] used similar acoustic features with statistical aggregators to classify types of connectional silences (emotional, compassionate, invitational). While the automated identification misidentified 41.3% of the clips, its use to semi-automate the annotation task for human annotators was significantly more efficient, manual annotation requiring 61% more time.

In the evaluation of non-verbal element of the interaction, studies prominently investigated the visual modality: studies have used face [40], gestures and movements [27, 18], gaze [38], posture [7], and a combination of them (head movements, posture, gaze, eyebrow, hand gesture, smile) in [39]. Another element of the communication investigated was the ongoing activity of the participant while the interaction was taking place. This included the clinician’s activities in Kocaballi et al. [20], and computer / screen interactions in Pearce et al. [38], both of which are known to affect consultations.

### 3.2. Theoretical background

The theoretical background for the evaluation of the communication was diverse. Eleven studies used ad-hoc coding systems, either designed and tailored for the study, derived from previous works by the same authors (e.g. Pearce et al. [38]), or inspired by concepts defined by existing framework but heavily modified (Two studies, [35]: modified Multi-Dimensional Interaction Analysis, [26]: ad-hoc set of acoustic features including mel-frequency cepstral coefficients (MFCC). The frameworks used in the studies can be separated into four (+ one) types.

The papers of the first type comprise assessment criteria of a medical authority (e.g. the Australian Open Disclosure Standard in [57]) and nor-malised assessment tools such as patients’ feedback tools, such as scales used to quantify anxiety (20-item State-Trait Anxiety Inventory), depression (15item Geriatric Depression Scale), and satisfaction with the appointment (Dementia Care Satisfaction Questionnaire) used by Sakai and Carpenter [46]. Two other studies used these scales to assess interaction quality: [58] (Dental Patient Feedback on Consultation skills), and [47].

The second type of framework are medical scales, used to evaluate the medical condition of patients such as the NSA16 in Chakraborty et al. [7]. Four different medical scales were used in the reviewed studies. The list is provided in table A.4.

The third type is frameworks for aspect-specific elements of the communication. The largest subset concerns semantic analysis of the interaction and linguistic or word based dictionaries, e.g. MetaMap for medical terms. Watson et al. [57] used Discursis, a visualisation tool for the analysis of term reuse. Sakai and Carpenter [46], Fridman et al. [17], Carnell et al. [6] and Venek et al. [52] used the LIWC, a word-based framework to quantify the frequency of terms and word categories (e.g. to quantify the use of possessives pronouns). Watson et al. [57] used a generic conversation and dialogue analysis tools, CAT, providing higher level structuring of the dialogue in terms of interpretability, discourse management, interpersonal control and emotional expression.

While a number of studies used acoustic and prosodic features, all studies have used their own set of features [26, 31], usually selected from a combination of sets used in other studies making it very difficult to compare their findings. It must be noted however that part of the feature selected in Manukyan et al. [26] is MFCC, a common set of acoustic features. The study of other non-verbal and paralinguistic aspects of the communication can be similarly depicted, i.e. extraction and study of ad-hoc sets of features, however one study [40] used the Emotion Facial Action Coding System (EmFACS) to code expressions of affects (happiness, social smiles, sadness, fear, anger, disgust and contempt) as well as social smiles and combinations of different affects.

The fourth type of framework used are the medical frameworks designed to study patient-clinician communication: VRCoDES [3], GMIAS [28, 56], the Comprehensive Analysis of the Structure of Encounters System CASES [28], and the Motivational Interviewing Skill Code (MISC) [49].

The Roter interaction analysis (RIAS) framework [43] is also referenced by Carnell et al. [6], although only its distinction between biomedical utterances and psychosocial utterances is used. Finally, six studies (e.g. [58]) did not use medical or conversational frameworks, instead reporting exploratory findings, for instance using data analysis (unsupervised machine learning methods such as principal component analysis) to identify prominent themes and observe the influence of their use on patients’ caregivers’ perceived quality of communication.

Similar to the variety of aspects investigated, the large set of frameworks used for reference or in the assessment reported in A.9 makes it difficult to compare the results of the studies and integrate them into a meta interpretation.

### 3.3. Paralinguistic and non-verbal communication

While the semantic aspect of the interaction has been frequently investigated, partly automated in the frame of this review but also more globally in observational studies of the clinician-patient interaction, non-semantic analysis of communication during consultations has been less studied. From the studies retrieved in this review, a number of aspects can be identified as promising.

*Visual cues* constitute the most frequent modality investigated. *Facial features* of the patient during communication has been used to detect facial expressions of different affects (happiness, social smiles, sadness, fear, anger, disgust, contempt) [40] in relation to signs of illness. Beyond the scope of this review, facial features were also used for the detection of illness. Barzilay et al. [2] classified patients’ affect using Face Action Recognition, noting the potential of the method as a clinician-supporting tool to detect schizophrenia. Joshi et al. [19] extracted generic facial spatio-temporal descriptors — Local Binary Patterns on Three Orthogonal Planes (LBP-TOP) and SpaceTime Interest Points (STIP) — as part of a multimodal classification model of depression (speech and video features), demonstrating the capacity of automated analysis to classify patients, but using extreme cases of the DSM-IV scale.

Focusing on *gaze and eye contact*, Pearce et al. [38] limited its use to detect computer activity while Porhet et al. [39] investigated gaze as elements of patterns of interaction (cues leading to cues in reaction) in verbal and nonverbal communication during consultations. Gaze was present in detected rules alongside other visual cues (nods, hand movements), however with low confidence scores for the strength of the observed patterns. Visual elements of bodily actions in time, *gestures and movements* have also been investigated.

The *posture* was investigated by Chakraborty et al. [7] to quantify symptoms of schizophrenia, finding a negative correlation between motor movements and negative symptoms.

Using a small number of interactions (*n* = 10) Mase et al. [27] analysed gestures as part of more abstracted interactional patterns. They did not analyse the gestures in themselves however, and their use was only as elements of sequential patterns for the interpretation of the interaction as a whole. At the smaller scale of motions realised during the interaction, Hart et al. [18] looked at interpersonal motions synchrony and mutual-followership between two communication styles in acted scenarios (disengaged, engaged). While their corpus is larger, investigation of real interactions would be required to validate these findings.

Finally, a few studies used a combination of visual cues. Porhet et al. [39] extracted head movements, posture, gaze, eyebrow, hand gesture, and smile to identify of cues leading to patients’ feedback in the form of rules (*X* = *Y* , e.g. doctor_head nod_ = patient_head nod_). They assessed the confidence of an extracted rule by computing the proportion of cases verifying the rule. While patterns of interactions were identified, low confidence (the confidence scores of the top 11 rules are between 0.36 and 0.12) and the acted nature of the data limits generalisability.

Finally, speech related investigations are mostly focused on *silences and pauses*. Identified as a significant component of the medical consultation, notably by Byrne and Heath [5], the therapeutic use of silences described in theoretical models can be detected using a systematic approach, while evidence of more complex usage and functions of pauses and silences is reported. Durieux et al. [12] investigated connectional silences: pauses between clinician’s and patient’s turns identified as potential markers of shared understanding and presence. They demonstrated the capacity of machine learning to detect connectional silences (recall 0.58, precision 1 compared with human coders) and support the annotation by human coders (human annotation without automation took 61% more time) but did not proceed to their analysis as a part of the communication beside a quantification over 32 samples. Conversational pauses are an element of the dynamics of the interaction (as a marker of engagement, power distribution, turn-taking, listening, connection, politeness, etc.). Manukyan et al. [26] investigated the performance of automated methods for the identification of conversational pauses, on its own (they report an accuracy of 94.4%) and as a supporting tool for manual coders (the annotation of one hour of audio took between 113 and 156 minutes for human coders, whereas the automated classification took 1.46 minute on a standard laptop). All studies used simple definitions of pauses, usually based on the length of silences (*t_duration_ >* 3*s*), and simple definition of pauses, i.e. not characterising types of pauses.

### 3.4. Methodologies employed

A first overview of the assessment of the studies (see table A.10) outlines shared limitations. Concerning research implications, reviewed studies used generally novel methodologies (23 out of 27), going beyond the simple application of existing tools. Replicability was low (10 studies) or partial (17), notably due to the expected unavailability of datasets. Generalisability was globally high (seventeen studies) with only five studies using a methodology tailored for a specific setting and five studies requiring sensible work to adapt it to other contexts. Concerning the evaluation of the risks of bias, the major limitation came from feature imbalance (25 studies) associated with a lack of suitable metrics in twelve studies. Fifteen did not provide contextualised results and six did not account for overfitting. Seven did not use real life settings (e.g. features simulated interactions), and ten had rather small sample size (seven used less than 50 documents, three less than 100). Regarding other limitations, automation was only partial in twenty-three studies, and the large majority (nineteen) required the transcription of the encounters while eighteen relied on the spoken content of the interaction (the difference is explained by one study that investigated phases of the interaction [4]).

Most studies (12) used supervised learning with common classifiers (e.g. decision trees, SVM, and neural networks) to predict a type of interaction at the utterance level (e.g. coarse coding of VRCoDES in Birkett et al. [3]) or at the session level (e.g. prediction of student success in Carnell et al. [6]). Tanana et al. [49] predicted of MISC behavioural codes at the utterance and session level, with good results at session level but low performance on utterances. Venek et al. [52] used conversation dynamic features, verbal information (topic identification) and acoustic features to classify non-suicidal and suicidal patients, and a second classification of repeaters and non-repeaters. The use of clinicians’ features in addition to patients’ features lead to a slight accuracy improvement (90% vs 85%) in the first step but marginally reduced the performance of the second step (-1.2%). Chakraborty et al. [7] had a similar task, correlating body movement and speech with prediction of negative symptoms of schizophrenia. This approach was used to assess successful interactions [47, 30, 6], to detect connecting silences [12] and in content-based analysis to classify topics [35], emotional valences [34], speech acts [28], and gain words [17].

Observational studies, identifying patterns from extracted features constitute another group of investigations. These studies focus on specific elements of the communication, such as semantic similarity between the patient and the physician, to find correlation between observed variations and expected dependant and independent variables. Word-based studies are common, including studies of dominance [46] anf temporal ordering of activities in [20]. Wong et al. [58] investigated word-related statistics (e.g. occurrence and co-occurrence) in relation with the perceived quality of the consultation by patients. Vrana et al. [54] searched semantic (dis-)similarities across patients and doctors of different ethnic backgrounds, observing significantly lower communication similarity from white physicians, controlling for confounders (gender of both participants). Other features were used. Rasting et al. [40] used facial display of affect to correlate patients expression with therapists emotional reactions. Porhet et al. [39] investigated sequences of multimodal behaviour elements that elicit feedback from patients. Pearce et al. [38] observed computer use behaviour. Mase et al. [27] identified points of interest in the recordings of trainings based on patterns of interactions (sequences of multimodal behaviour).

Another group of studies performed clustering to detect types of interactions (unsupervised learning, e.g. grouping clinicians by style of communication), or to distinguish between known groups (supervised learning, e.g. interaction featuring good and bad communication). For instance, Wallace et al. [56] clustered physicians based on turn-taking patterns and speech act transitions through semantics, detecting two clusters corresponding to the difference in patients’ evaluation for three categories of questions investigated. Cuffy et al. [10] captured semantic aspects of communication, notably the relatedness between discourse content (however limited by the small scale of the study and the disparity between computed scores and self-reported questionnaires). Using the opposite approach (i.e. using fixed groups), Watson et al. [57] extracted word-related statistics on topics to compare speech during effective and ineffective interactions, as evaluated by experts using behavioural analysis, and found significant difference between effective and ineffective interactions in four out of five aspect of the communication. Manukyan et al. [26] evaluated the performance of automating the detection of conversational pauses with good results (accuracy=94.4%). Chiba et al. [8] investigated differences in topics found in conversations between doctors interacting with caregivers of patients who died at home or at hospital. Blomqvist et al. [4] found differences between patients with and without attention deficit and hyperactivity disorder ADHD (higher degree of non-coordination for patients with ADHD).

Finally, some studies used a combination of approaches, for instance Hart et al. [18] first conducted an exploration of motion synchrony between patients and nurses, before classifying interactions using engaged and disengaged scenarios (accuracy=0.72%)

## 4. Conclusion

Many of the studies identified reviewed used structured or semi-structured interviews, featuring more restricted interactions than in medical consultations. While this helps retrieving investigated cues (behaviours, emotions, gestures) more consistently, it also limits the weight of the findings of these studies as regards the less restricted range of interaction that take place during medical consultations.

A number of aspects of the patient-clinician communication, verbal and non-verbal, have been investigated using systematic approaches to facilitate objective evaluations. However, while much of the focus has been set on the semantic of the interaction, investigations using paralinguistic and nonverbal components are much less common. In fact, the analysis of non-verbal behaviour has focused more on visual aspects (face, posture, movements).

The analysis of speech is fairly common in studies seeking to discriminate impaired speech of a person (e.g. patients affected by a physical or neurological conditions). However, the characterisation of speech during the patient-clinician communication is mostly limited to the quantification of silences and pauses using simple definitions.

While some touched upon some of its elements, very few studies have investigated the structure of turns in the interaction. Turn-taking behaviours combined with the analysis of speech patterns remains an area that was not investigated, supporting and legitimating the focus of the work. The rather unexplored domain of the paralinguistic and non-verbal elements associated with the automation of the assessment of the communication happening in consultations constitutes its background [44].

Overall, the result of this review shows that the automated analysis of consultation is feasible. Numerous elements of the communication happening during medical encounters can be retrieved and analysed automatically.

The literature focuses largely on semantics, while little work exists on paralinguistic analysis. Methodologies employed in consultation analysis vary. Whereas semantic analysis often use existing frameworks as a basis, studies of non-verbal and paralinguistic communication shared little methodological common ground. Much remains to be done to standardise elements, features, and metrics for the analysis of medical consultations. While majority of studies we reviewed used automation in classification tasks, exploration and identification of patterns of interaction is also a focus of research. The results of this review shows that features of multimodal behaviour in consultations can be extracted and identified. The characterisation of these features complement existing knowledge on elements of patient-clinician communication that are new and complementary, notably relating to linguistic, paralinguistic and non-verbal behaviour.

## Data Availability

All data produced in the present work are contained in the manuscript

## Acknowledgements

This research received funding from the Health Research Board, Ireland, towards the INCA project (Interaction Analytics for Automatic Assessment of Communication in Healthcare).

## Appendix A. Supplemental tables

**Table A.3:** List of abbreviation for methods and terms used in studies reported in the review.

| Abbreviation | Full name |
| --- | --- |
| ANOVA | analyses of variance |
| BoW | bag of words |
| BTS | Bartlett’s test of sphericity |
| CFR | conditional random fields |
| CNN | Counterpropagation Neural Networks |
| DT | Decision Tree |
| GRU | gated recurrent units |
| GT | golden truth |
| HMM | hidden Markov models |
| HGRU | hierarchical gated recurrent units |
| JSD | Jensen-Shannon Divergence |
| KLD | Kullback-Leibler Divergence |
| KMO | Kaiser-Meyer-Olkin |
| kNN | k-nearest neighbors |
| LIWC | Linguistic Inquiry and Word Count |
| LR | logistic regression |
| LSM | Language style matching |
| NB | Naïve Bayes |
| RNN | recurrent neural network |
| SVM | Support Vector Machine |
| t-tests | Student’s t-tests |
| tf-idf | term frequency-inverse document frequency |

**Table A.4:** List of questionnaires (top) and medical scales (bottom) used in studies reported in the review.

| Abbreviation | Full name |
| --- | --- |
| DCSQ | Dementia Care Satisfaction Questionnaire |
| DPFC | 16-item Dental Patient Feedback on Consultation skills |
| GDS | Geriatric Depression Scale |
| MISC | Motivational Interviewing Skill Code |
| NSA | Negative Symptom Assessment |
| STAI | State-Trait Anxiety Inventory |
| TAS | Toronto Alexithymia Scale (TAS-26) |

**Table A.5:**
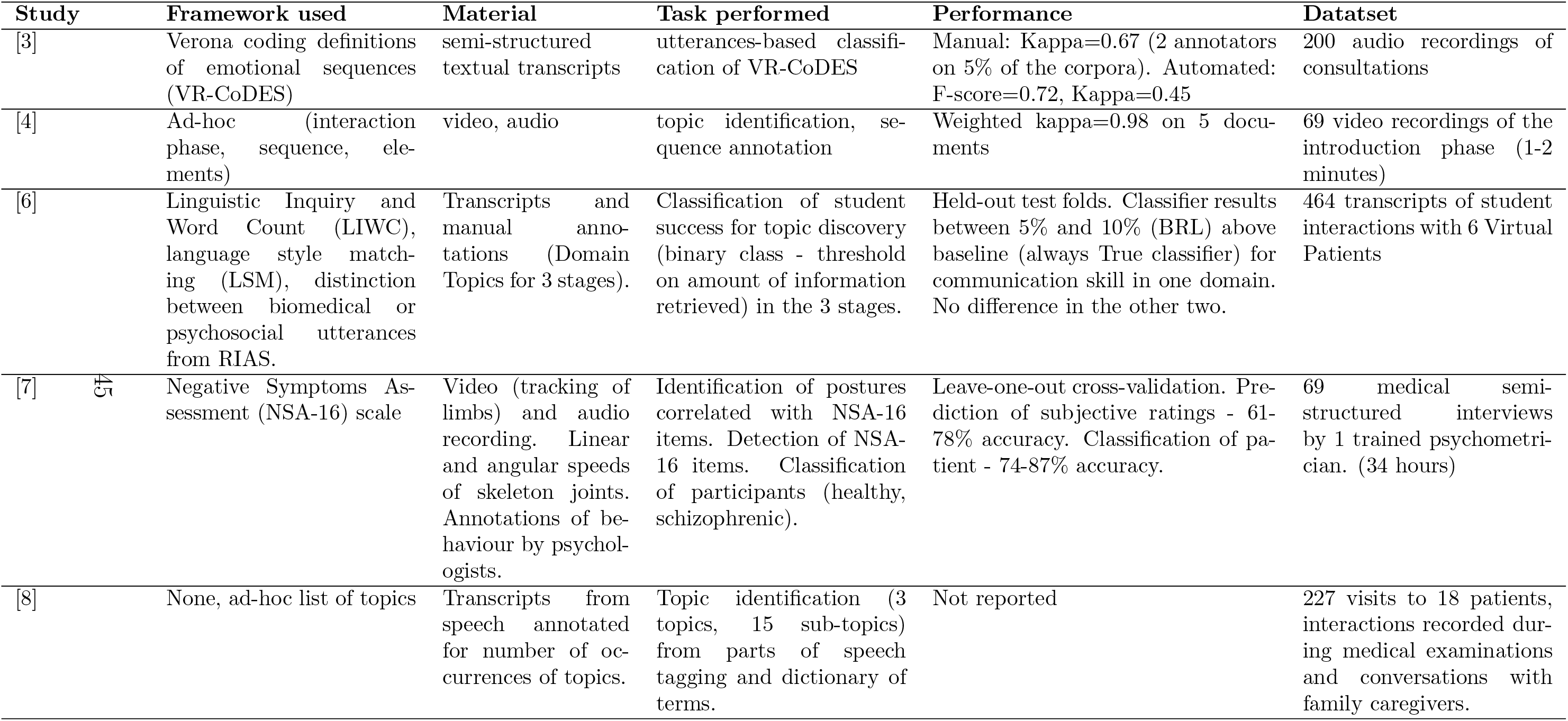
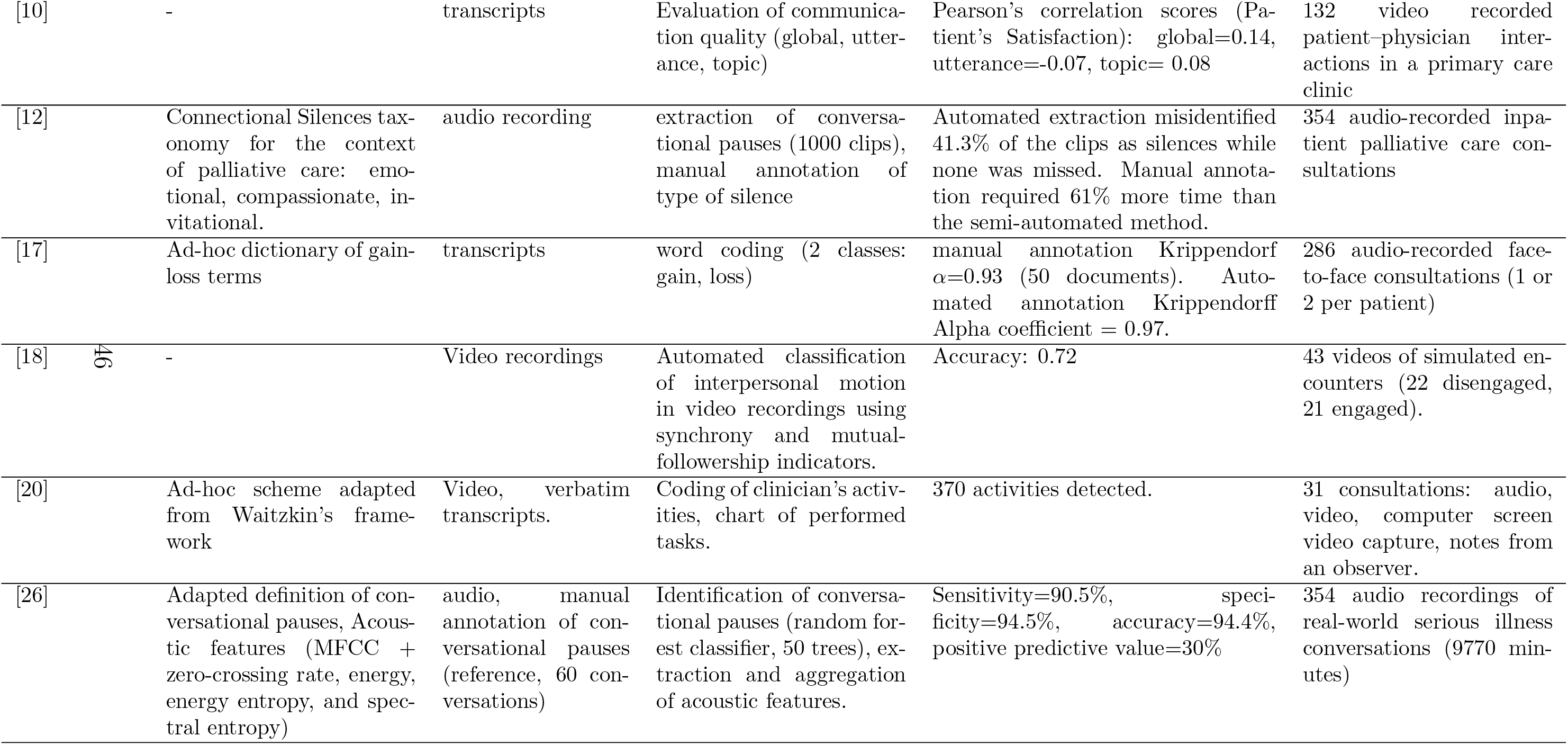
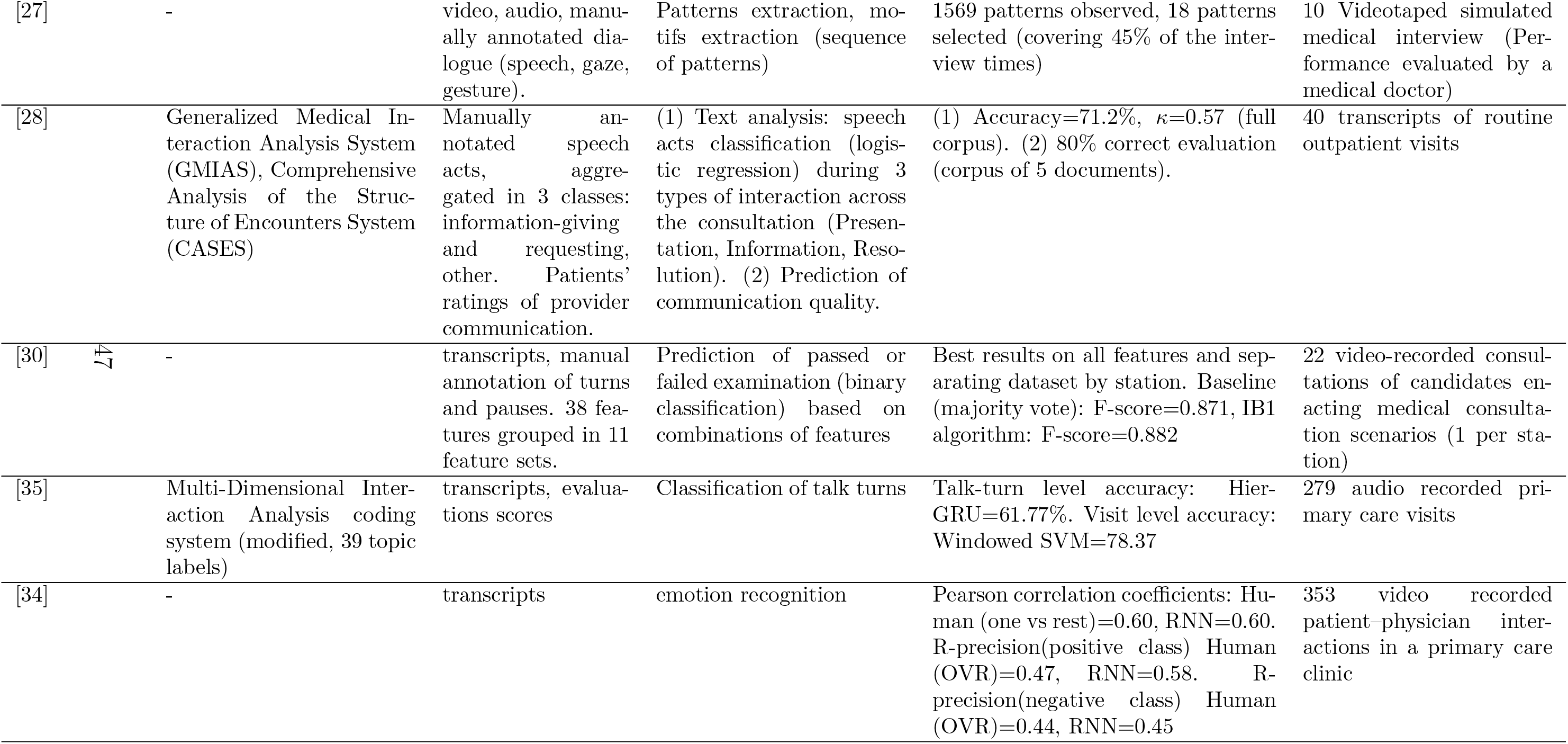
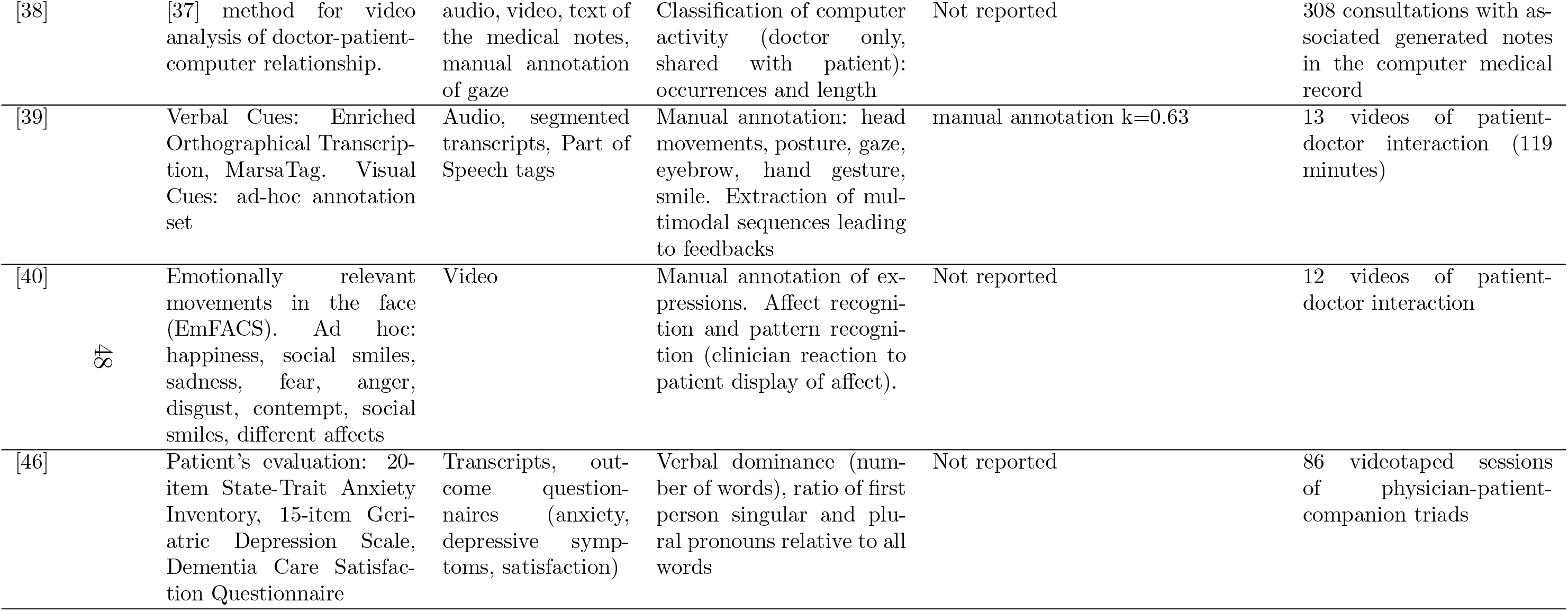
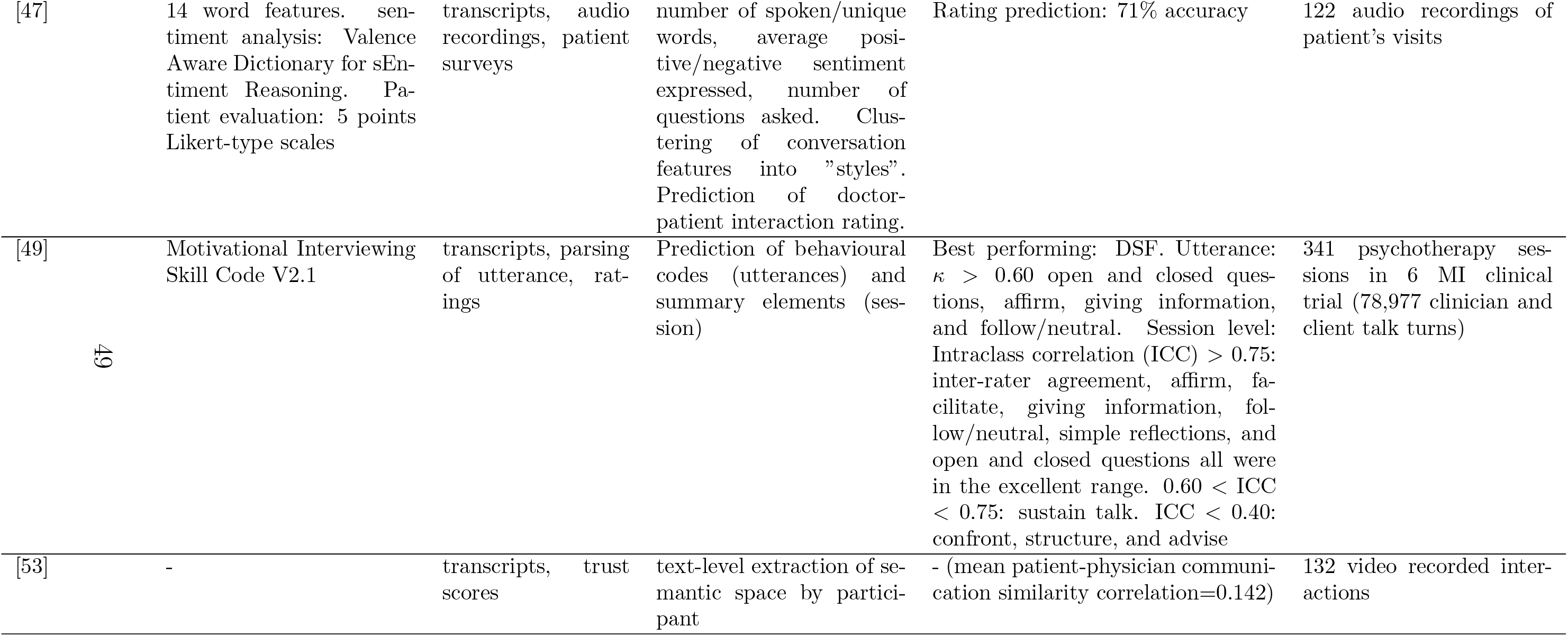
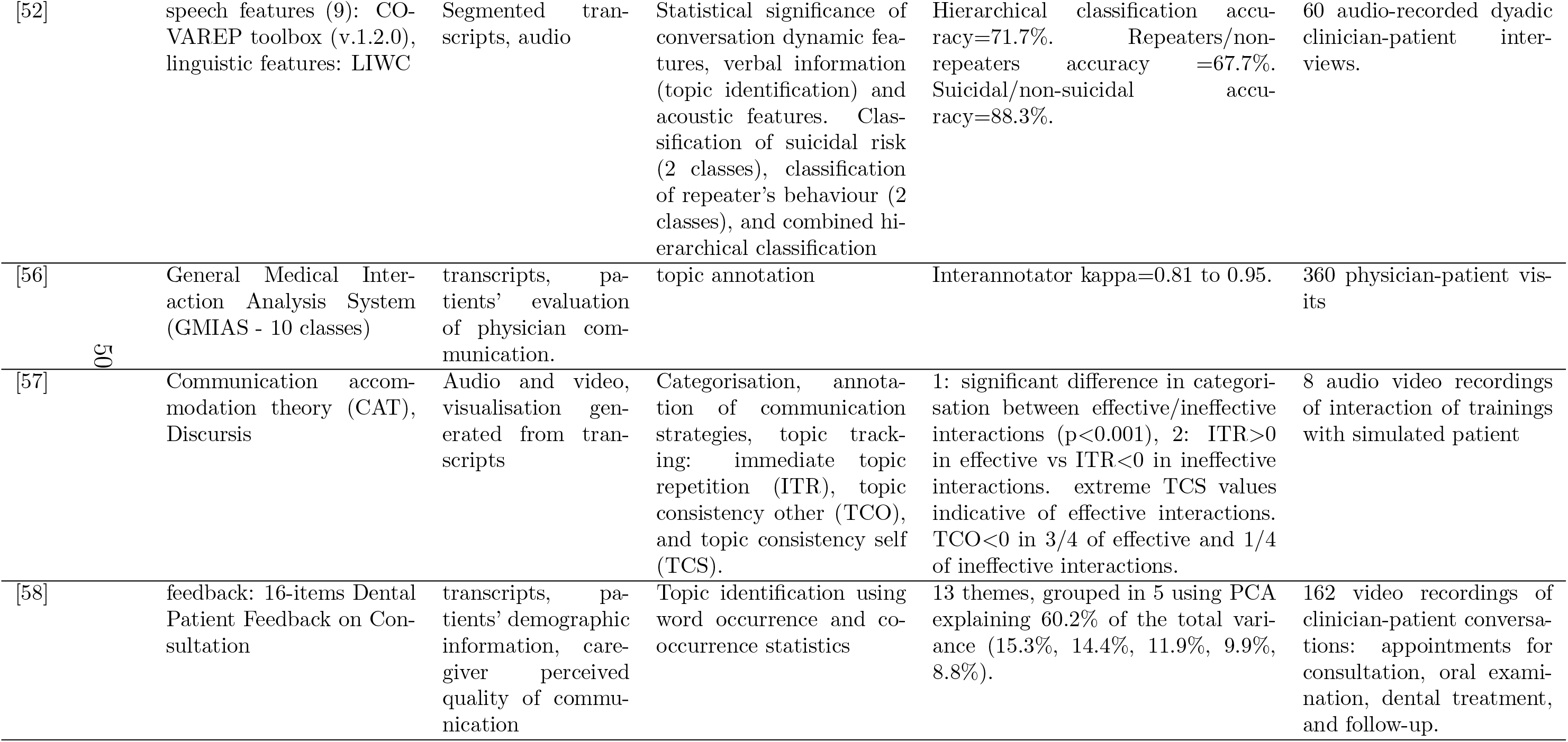
Tasks table.

**Table A.6:**
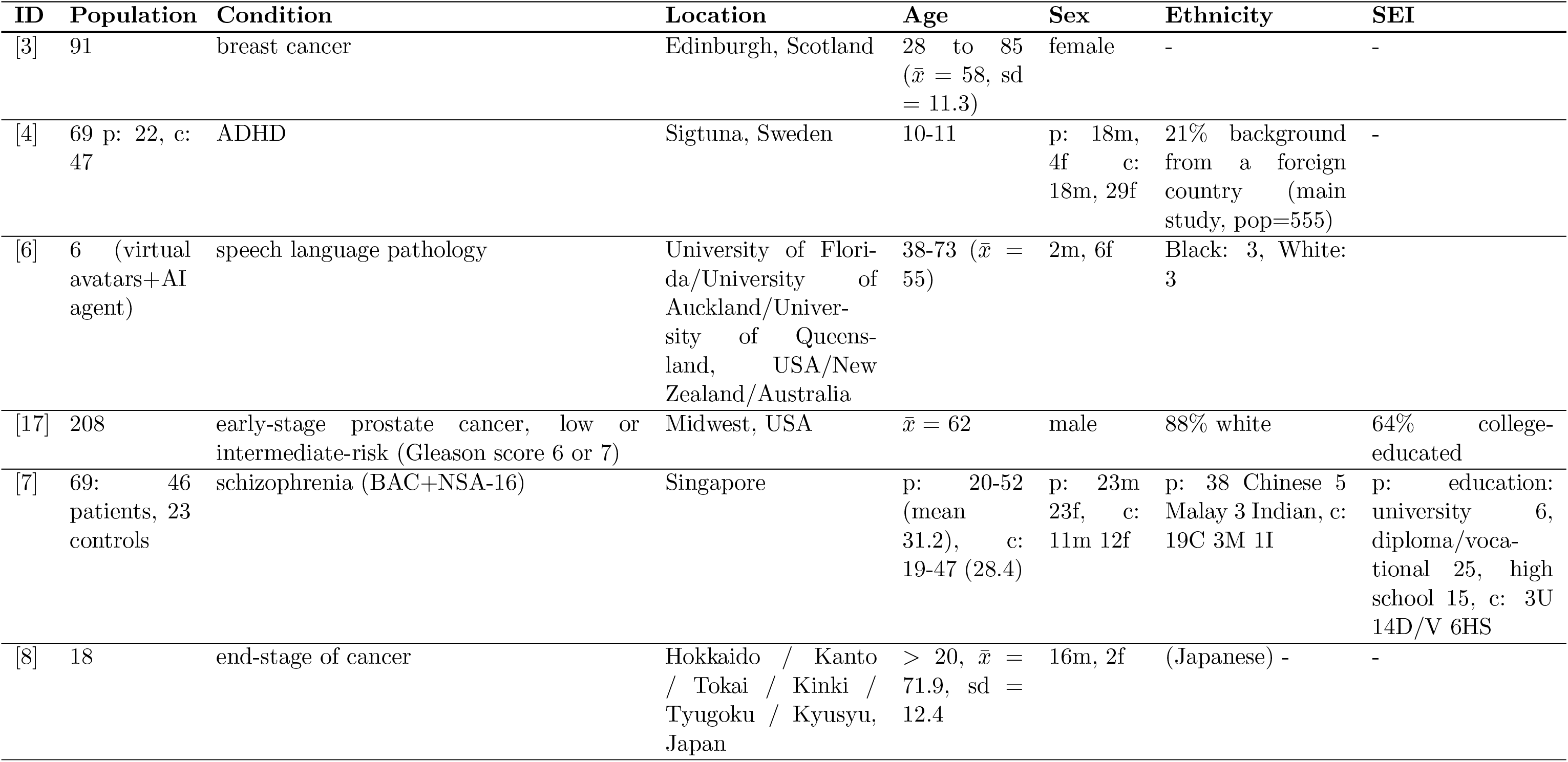
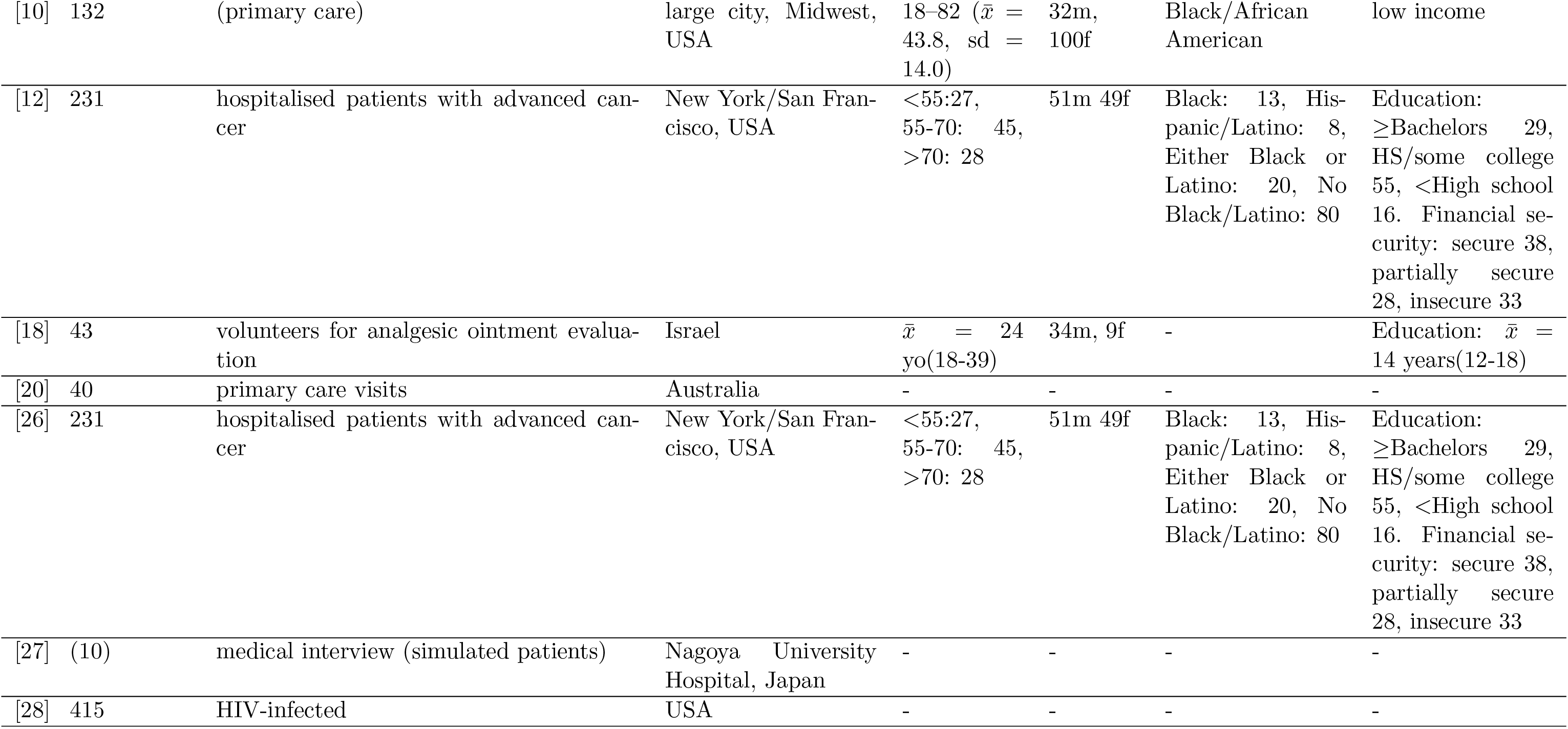
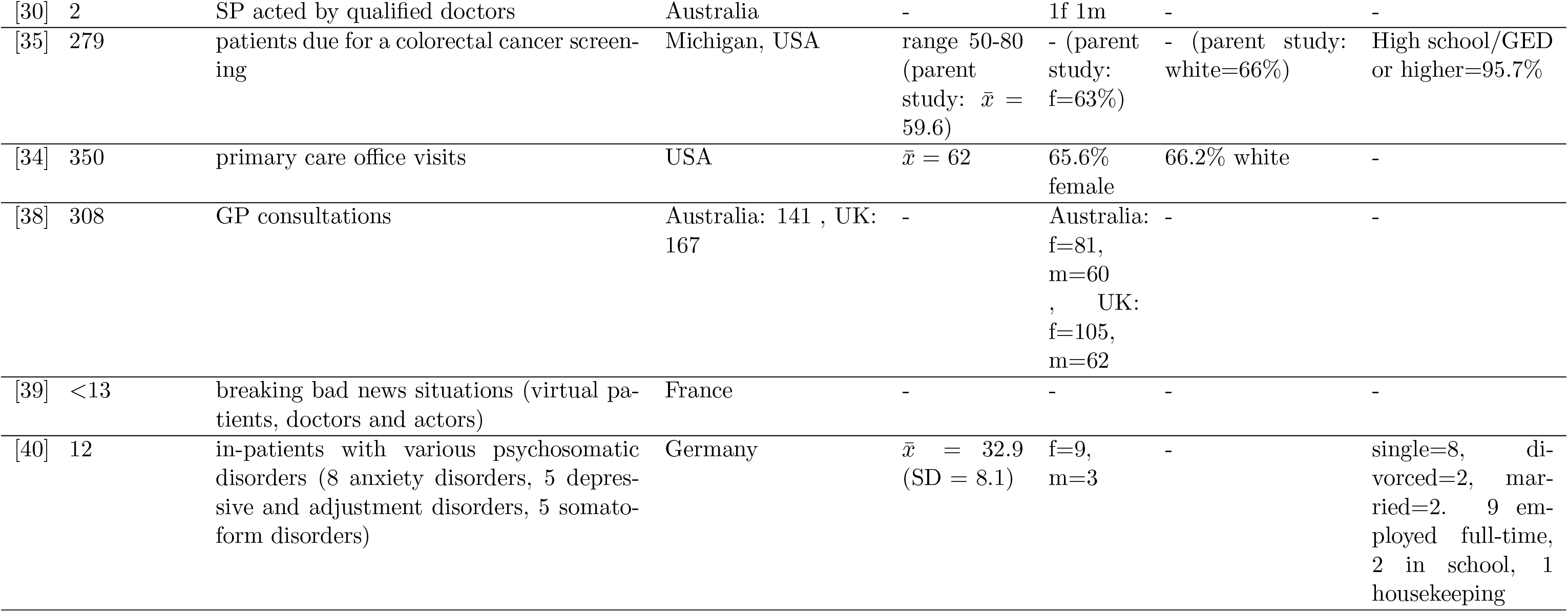
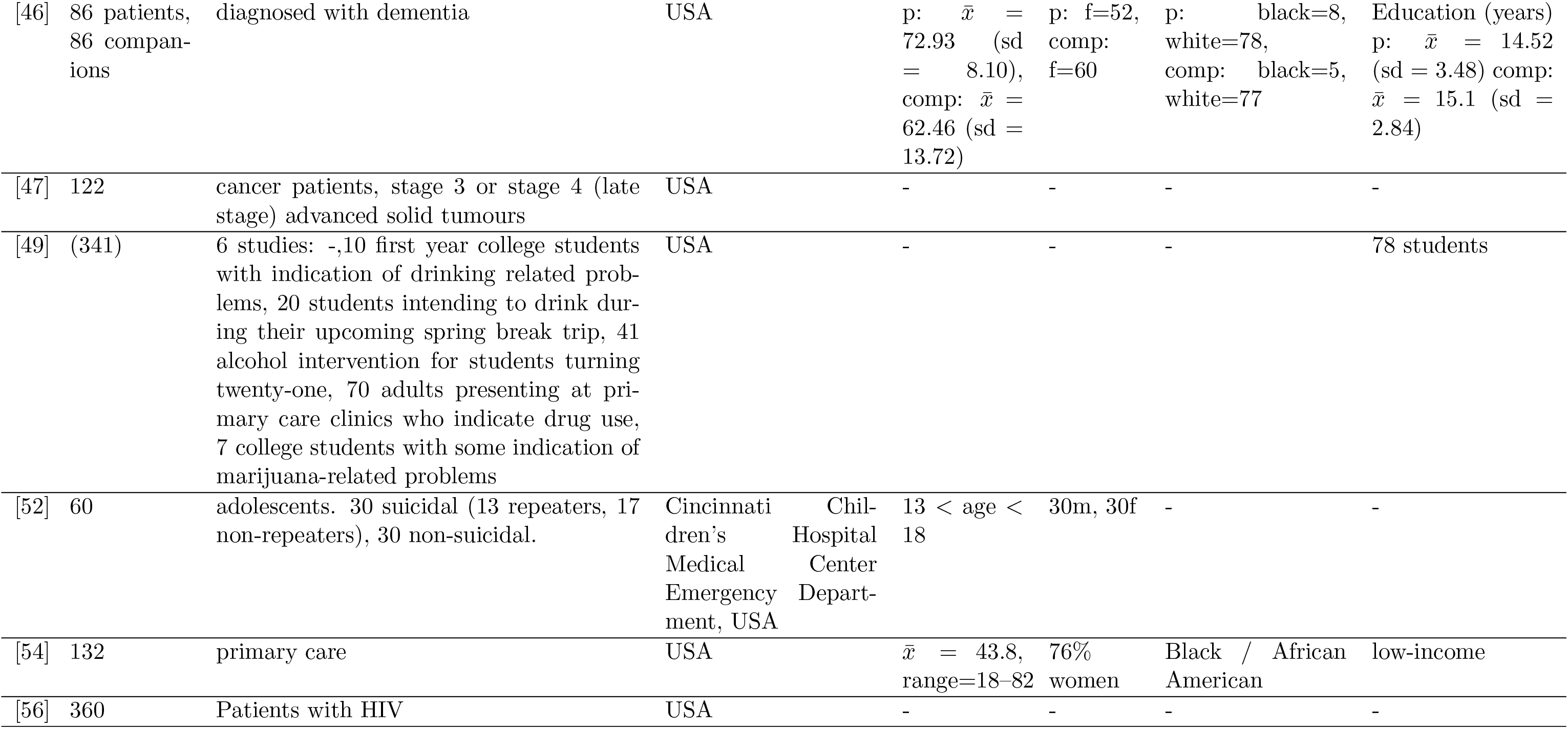
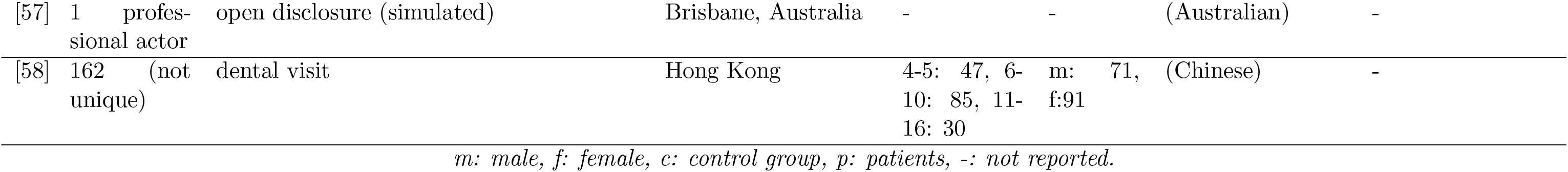
Patients information. (SEI = socioeconomic information; *x̄* = mean)

| ID | Population | Condition | Location | Age | Sex | Ethnicity | SEI |
| --- | --- | --- | --- | --- | --- | --- | --- |
| [3] | 91 | breast cancer | Edinburgh, Scotland | 28 to 85<br>($\bar{x} = 58$, sd = 11.3) | female | - | - |
| [4] | 69 p: 22, c: 47 | ADHD | Sigtuna, Sweden | 10-11 | p: 18m, 4f<br>c: 18m, 29f | 21% background from a foreign country (main study, pop=555) | - |
| [6] | 6 (virtual avatars+AI agent) | speech language pathology | University of Florida/University of Auckland/University of Queensland, USA/New Zealand/Australia | 38-73 ($\bar{x} = 55$) | 2m, 6f | Black: 3, White: 3 | |
| [17] | 208 | early-stage prostate cancer, low or intermediate-risk (Gleason score 6 or 7) | Midwest, USA | $\bar{x} = 62$ | male | 88% white | 64% college-educated |
| [7] | 69: 46 patients, 23 controls | schizophrenia (BAC+NSA-16) | Singapore | p: 20-52 (mean 31.2), c: 19-47 (28.4) | p: 23m 23f, c: 11m 12f | p: 38 Chinese 5 Malay 3 Indian, c: 19C 3M 1I | p: education: university 6, diploma/vocational 25, high school 15, c: 3U 14D/V 6HS |
| [8] | 18 | end-stage of cancer | Hokkaido / Kanto / Tokai / Kinki / Tyugoku / Kyusyu, Japan | $> 20$, $\bar{x} = 71.9$, sd = 12.4 | 16m, 2f | (Japanese) - | - |
| [10] | 132 | (primary care) | large city, Midwest, USA | 18–82 ($\bar{x}$ = 43.8, sd = 14.0) | 32m, 100f | Black/African American | low income |
| [12] | 231 | hospitalised patients with advanced cancer | New York/San Francisco, USA | <55:27, 55-70: 45, >70: 28 | 51m 49f | Black: 13, Hispanic/Latino: 8, Either Black or Latino: 20, No Black/Latino: 80 | Education: ≥Bachelors 29, HS/some college 55, <High school 16. Financial security: secure 38, partially secure 28, insecure 33 |
| [18] | 43 | volunteers for analgesic ointment evaluation | Israel | $\bar{x}$ = 24 yo(18-39) | 34m, 9f | - | Education: $\bar{x}$ = 14 years(12-18) |
| [20] | 40 | primary care visits | Australia | - | - | - | - |
| [26] | 231 | hospitalised patients with advanced cancer | New York/San Francisco, USA | <55:27, 55-70: 45, >70: 28 | 51m 49f | Black: 13, Hispanic/Latino: 8, Either Black or Latino: 20, No Black/Latino: 80 | Education: ≥Bachelors 29, HS/some college 55, <High school 16. Financial security: secure 38, partially secure 28, insecure 33 |
| [27] | (10) | medical interview (simulated patients) | Nagoya University Hospital, Japan | - | - | - | - |
| [28] | 415 | HIV-infected | USA | - | - | - | - |
| [30] | 2 | SP acted by qualified doctors | Australia | - | 1f 1m | - | - |
| [35] | 279 | patients due for a colorectal cancer screening | Michigan, USA | range 50-80<br>(parent study: $\bar{x}$ = 59.6) | - (parent study: f=63%) | - (parent study: white=66%) | High school/GED or higher=95.7% |
| [34] | 350 | primary care office visits | USA | $\bar{x}$ = 62 | 65.6% female | 66.2% white | - |
| [38] | 308 | GP consultations | Australia: 141 , UK: 167 | - | Australia: f=81, m=60 , UK: f=105, m=62 | - | - |
| [39] | <13 | breaking bad news situations (virtual patients, doctors and actors) | France | - | - | - | - |
| [40] | 12 | in-patients with various psychosomatic disorders (8 anxiety disorders, 5 depressive and adjustment disorders, 5 somatoform disorders) | Germany | $\bar{x}$ = 32.9 (SD = 8.1) | f=9, m=3 | - | single=8, divorced=2, married=2. 9 employed full-time, 2 in school, 1 housekeeping |
| [46] | 86 patients, 86 companions | diagnosed with dementia | USA | p: $\bar{x} = 72.93$ (sd = 8.10), comp: $\bar{x} = 62.46$ (sd = 13.72) | p: f=52, comp: f=60 | p: black=8, white=78, comp: black=5, white=77 | Education (years) p: $\bar{x} = 14.52$ (sd = 3.48) comp: $\bar{x} = 15.1$ (sd = 2.84) |
| [47] | 122 | cancer patients, stage 3 or stage 4 (late stage) advanced solid tumours | USA | - | - | - | - |
| [49] | (341) | 6 studies: -,10 first year college students with indication of drinking related problems, 20 students intending to drink during their upcoming spring break trip, 41 alcohol intervention for students turning twenty-one, 70 adults presenting at primary care clinics who indicate drug use, 7 college students with some indication of marijuana-related problems | USA | - | - | - | 78 students |
| [52] | 60 | adolescents. 30 suicidal (13 repeaters, 17 non-repeaters), 30 non-suicidal. | Cincinnati Children's Hospital Medical Center Emergency Department, USA | 13 < age < 18 | 30m, 30f | - | - |
| [54] | 132 | primary care | USA | $\bar{x} = 43.8$, range=18–82 | 76% women | Black / African American | low-income |
| [56] | 360 | Patients with HIV | USA | - | - | - | - |
| [57] | 1 profes-<br>sional actor | open disclosure (simulated) | Brisbane, Australia | - | - | (Australian) | - |
| [58] | 162 (not<br>unique) | dental visit | Hong Kong | 4-5: 47, 6-10: 85, 11-16: 30 | m: 71,<br>f:91 | (Chinese) | - |

*m: male, f: female, c: control group, p: patients, -: not reported.*

**Table A.7:**
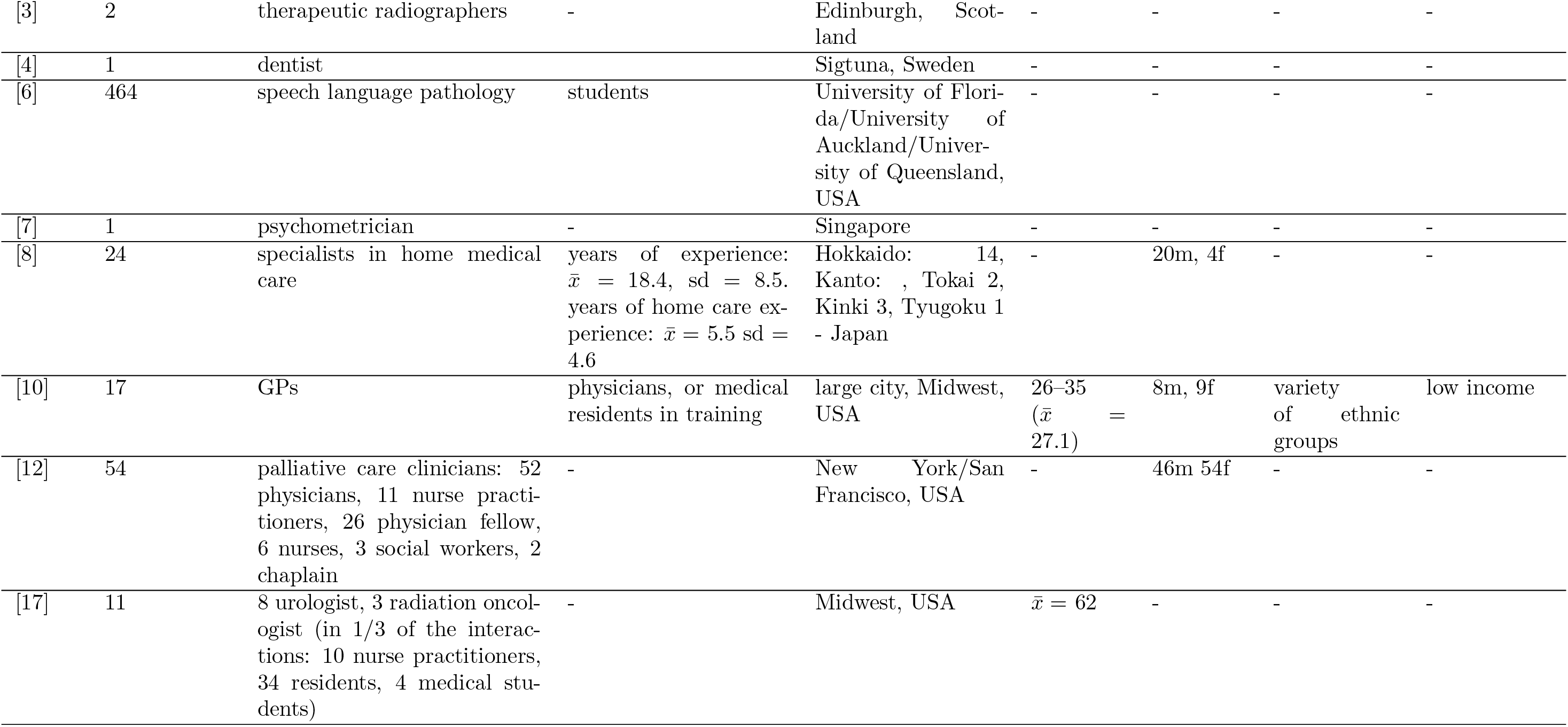
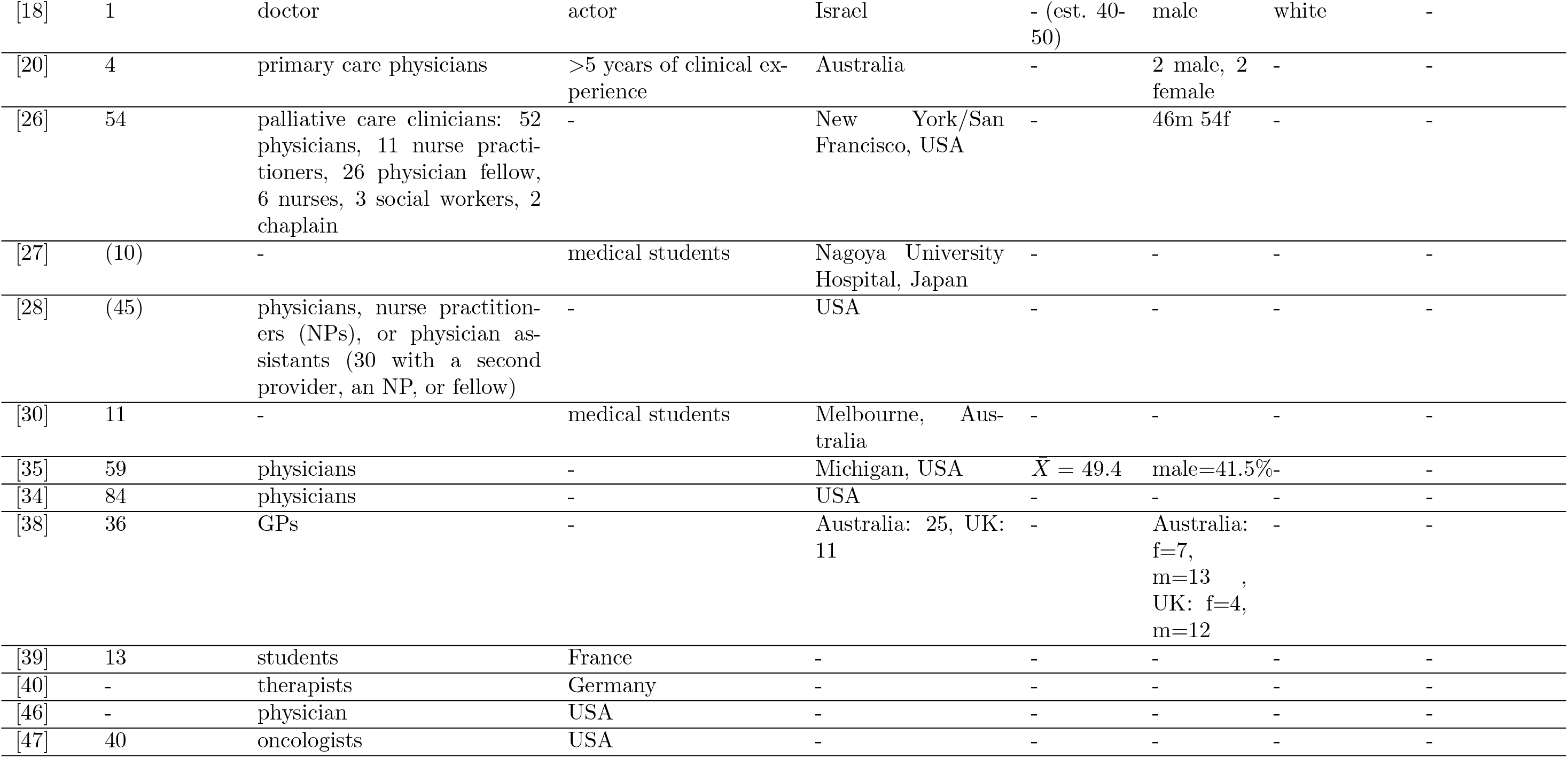
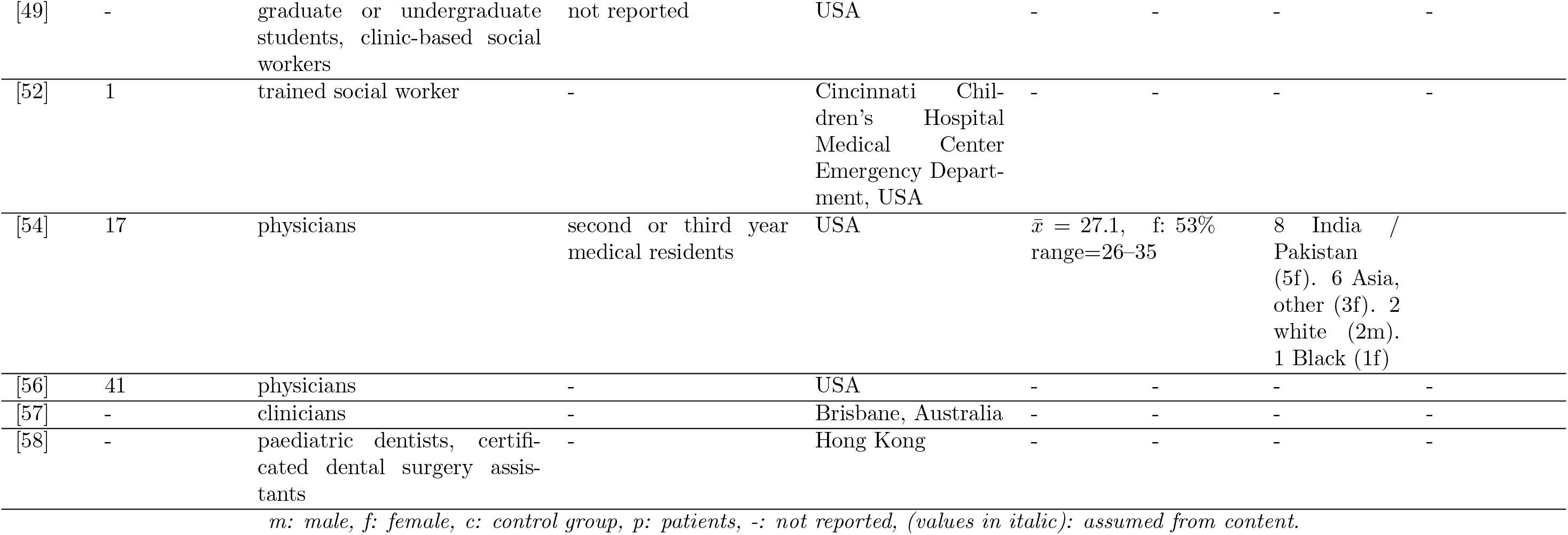
Clinicians information. (SEI = socioeconomic information)

**Table A.8:** Interactions.

| ID | Type of interaction | Medical interaction |
| --- | --- | --- |
| [3] | dyadic | breast cancer consultations |
| [4] | triadic, analysis: dyadic only | introduction phase of dentist visits |
| [6] | dyadic | GP consultations |
| [7] | dyadic | semi-structured interviews |
| [8] | triadic | medical examinations and conversations with family caregivers |
| [10] | dyadic | patient–physician interactions in a primary care clinic |
| [12] | dyadic | palliative care consultations |
| [17] | dyadic | medical interview (discuss treatment options). 1/3 visits included a discussion with a nurse practitioner or resident before the interview |
| [18] | dyadic | presentation and instructions of a drug for an evaluation study (simulated). Engaged scenario: opened questions. Disengaged scenario closed questions |
| [20] | dyadic | GP consultation |
| [26] | dyadic | palliative care consultations |
| [27] | dyadic | Simulated medical interview |
| [28] | dyadic (36 with a second clinician) | routine outpatient visits of HIV patients |
| [30] | dyadic | OSCE examinations: sexually transmitted disease – genital herpes and bowel cancer (scripted, uncued, free from) |
| [35] | dyadic, small fraction triadic (nurse, family member) | primary care visits (preventive health discussions.) |
| [34] | dyadic | elderly patients primary care office visits |
| [38] | dyadic | GP consultations |
| [39] | dyadic | breaking bad news, real training sessions |
| [40] | dyadic | intake interviews for in-patient psychotherapy |
| [46] | triadic | dementia diagnosis disclosure sessions with the patient and companion |
| [47] | triadic (family caregiver) | physician-patient visits |
| [49] | dyadic | Motivational Interviewing (substance use disorders) |
|  | *OSCE: Objective structured clinical examination* |  |

| **ID** | **Type of inter-**<br>**action** | **Medical interaction** |
| --- | --- | --- |
| [52] | dyadic | interviews, 16 questions (Columbia Suicide Severity Rating Scale (C-SSRS version 1/14/2009), Suicidal Ideation Questionnaire-Junior (SIQ-JR version 1987), Ubiquitous Questionnaire (UQ version 2011). |
| [54] | dyadic | primary care |
| [56] | dyadic | adherence dialogue in HIV care |
| [57] | dyadic | discussions about adverse events in patient care, open disclosure interactions scenarios taken from actual adverse events |
| [58] | triadic | examination: 60, treatment: 71, consultation: 31 |

*OSCE: Objective structured clinical examination*

**Table A.9:**
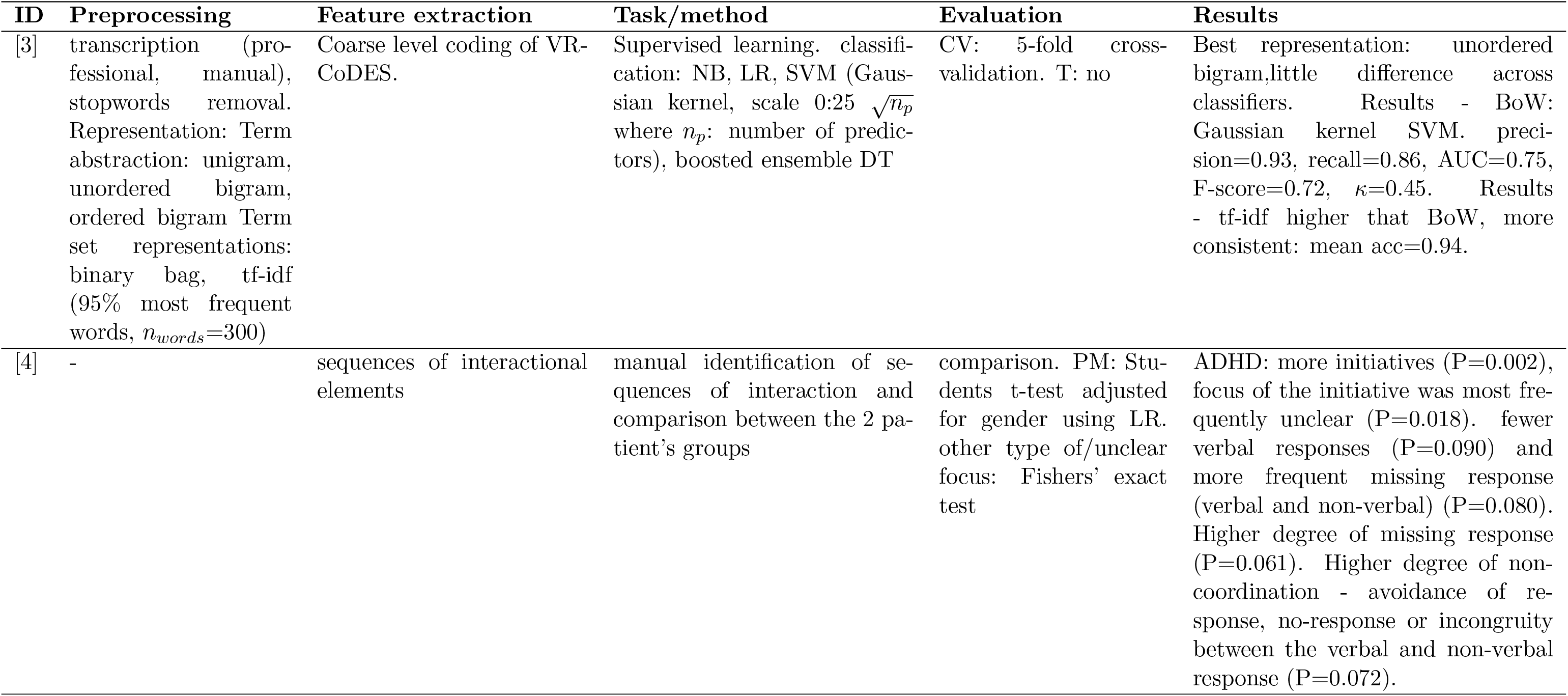
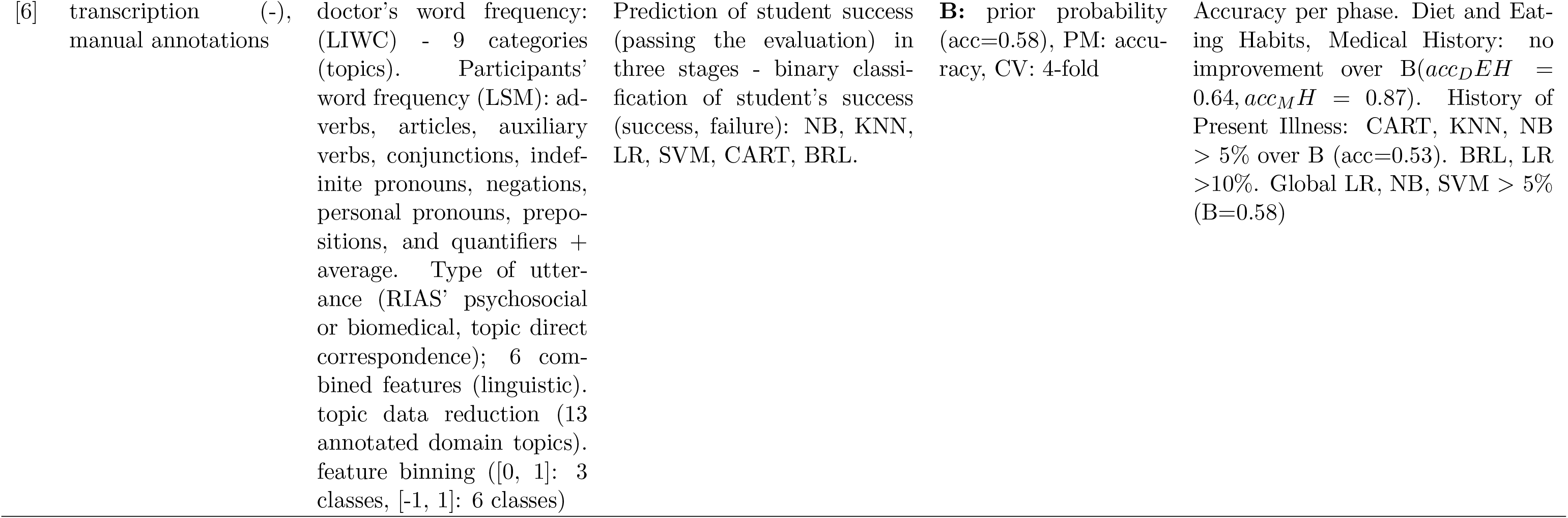
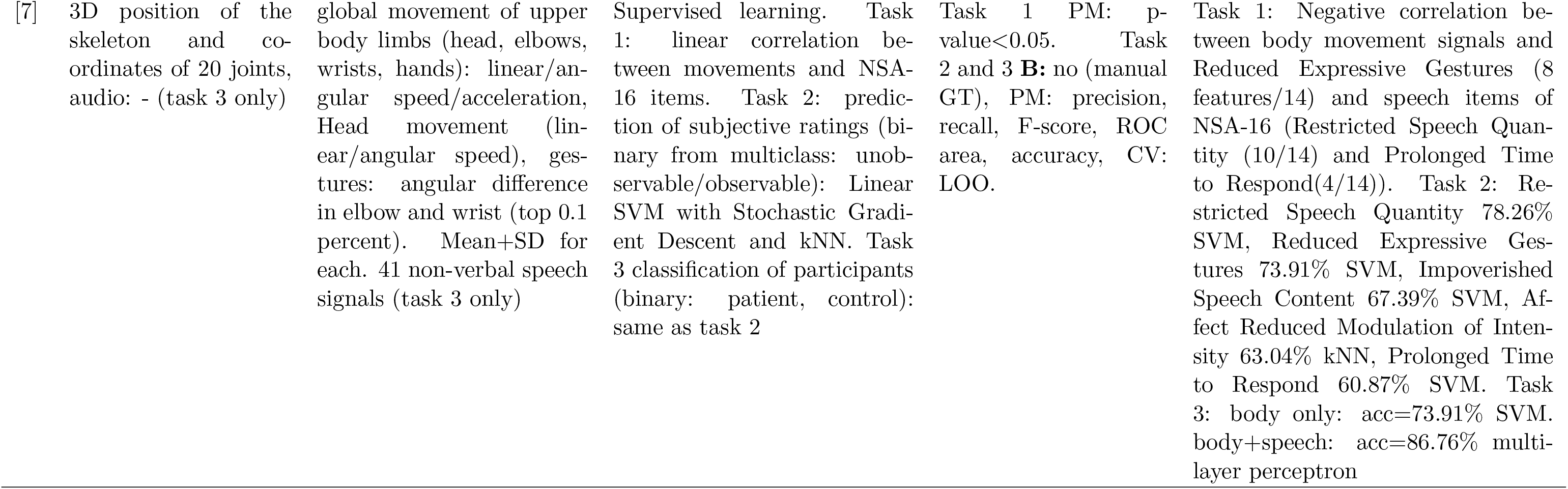
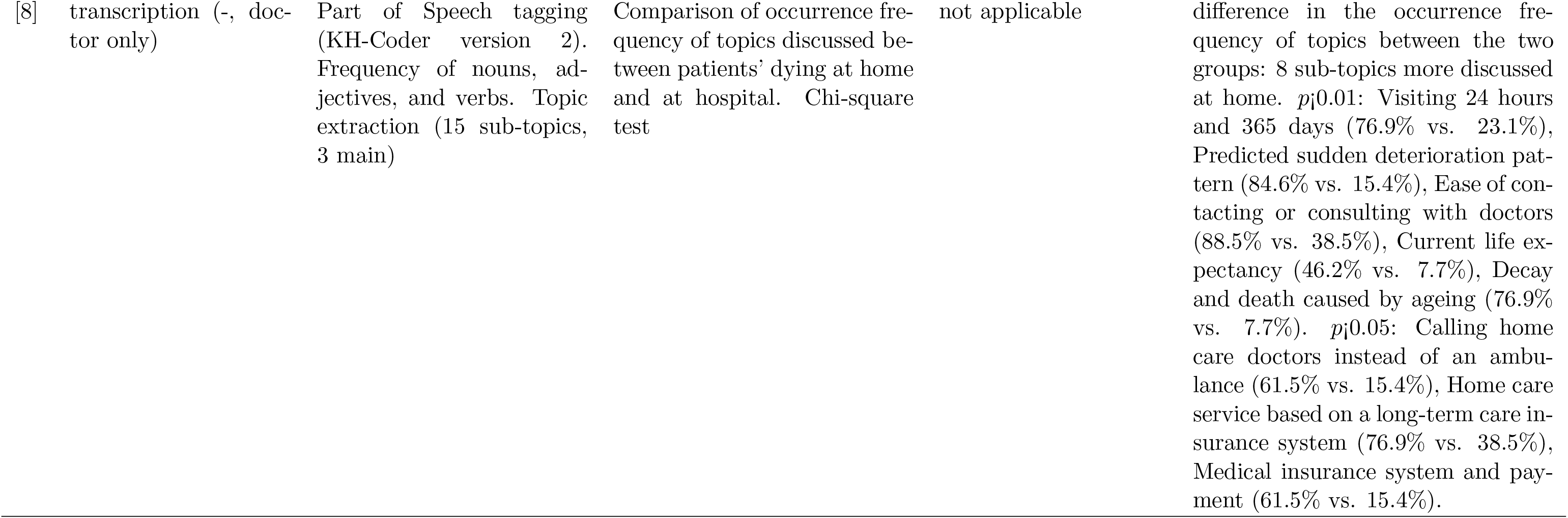
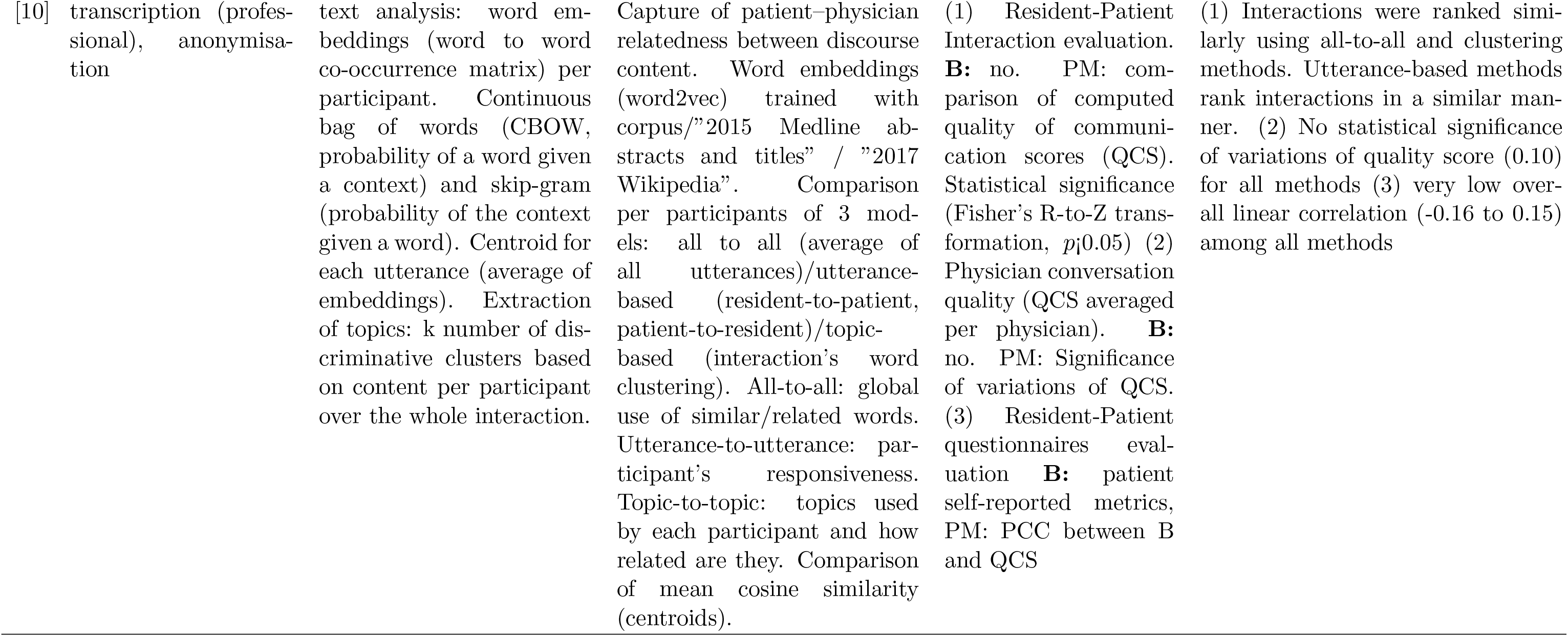
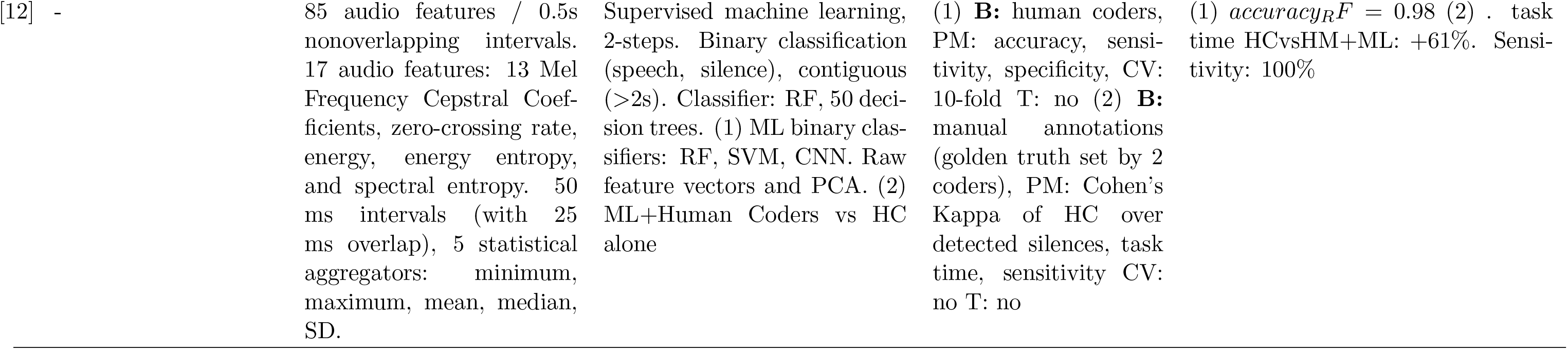
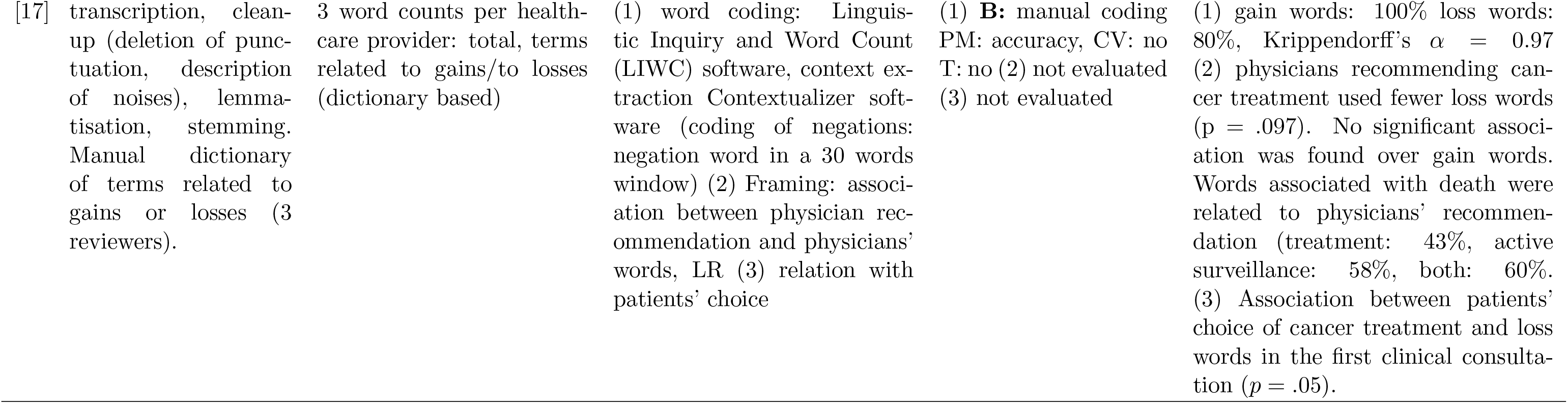
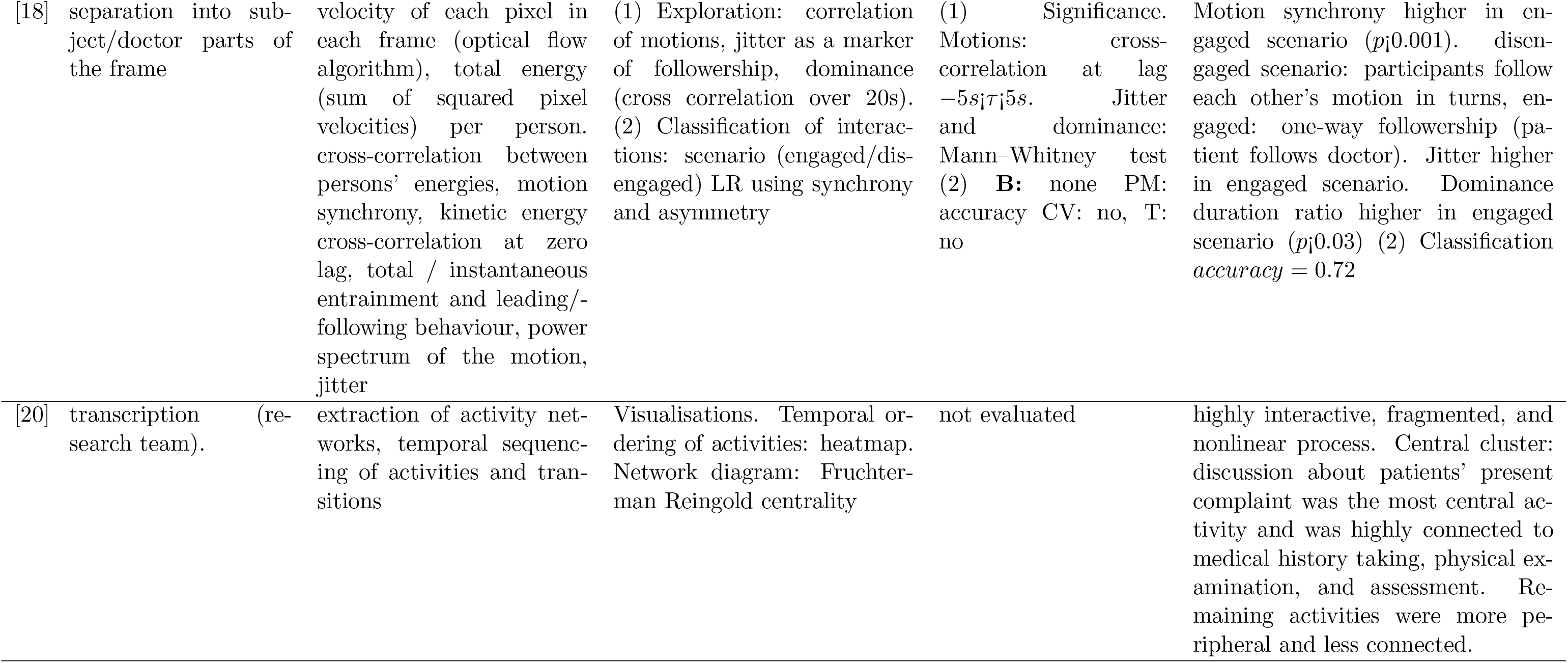
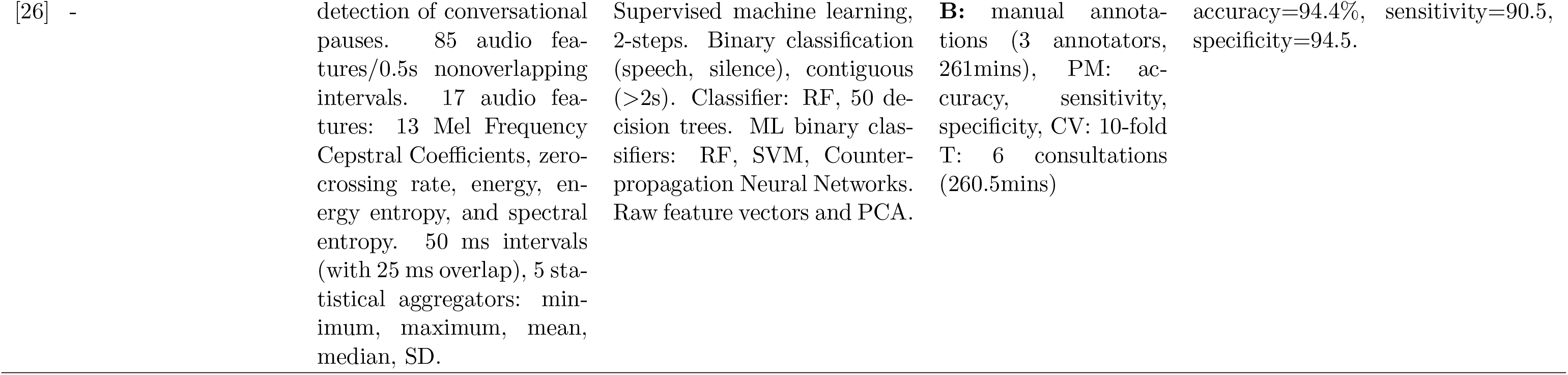
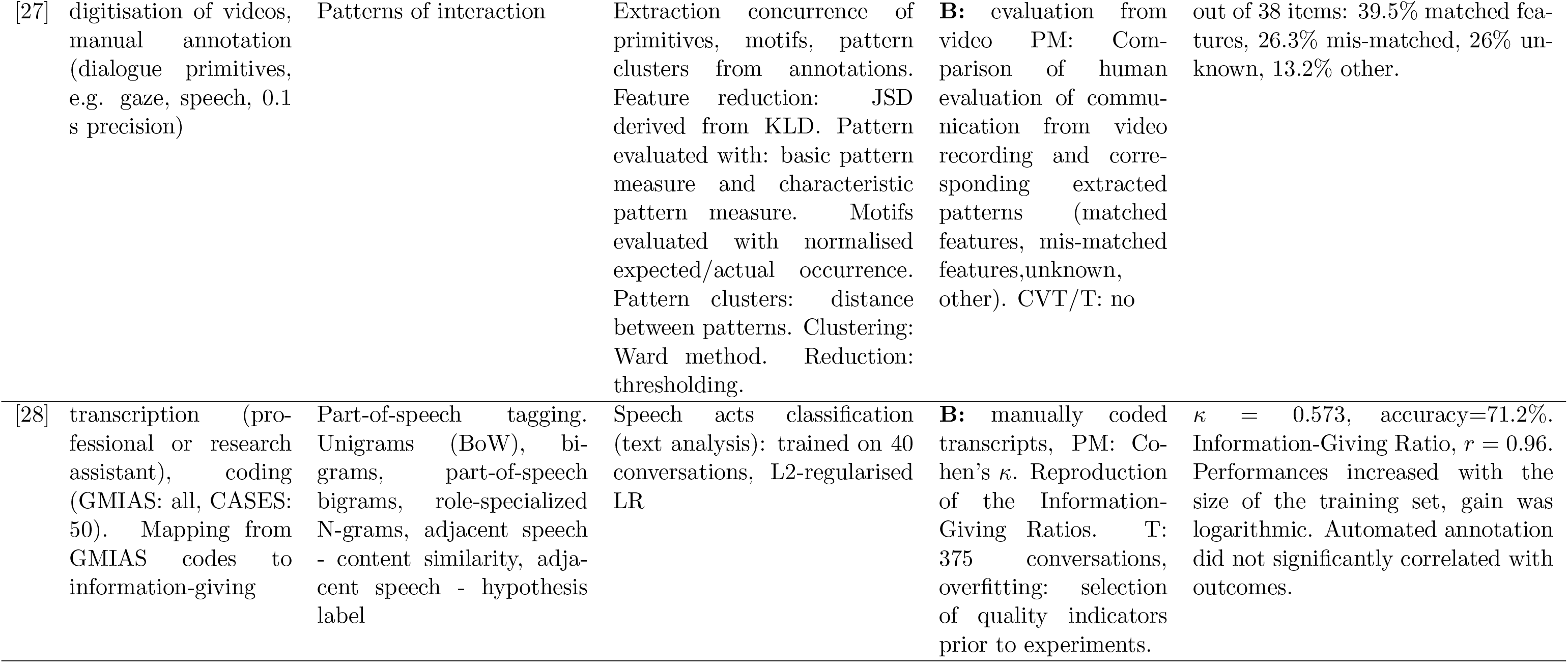
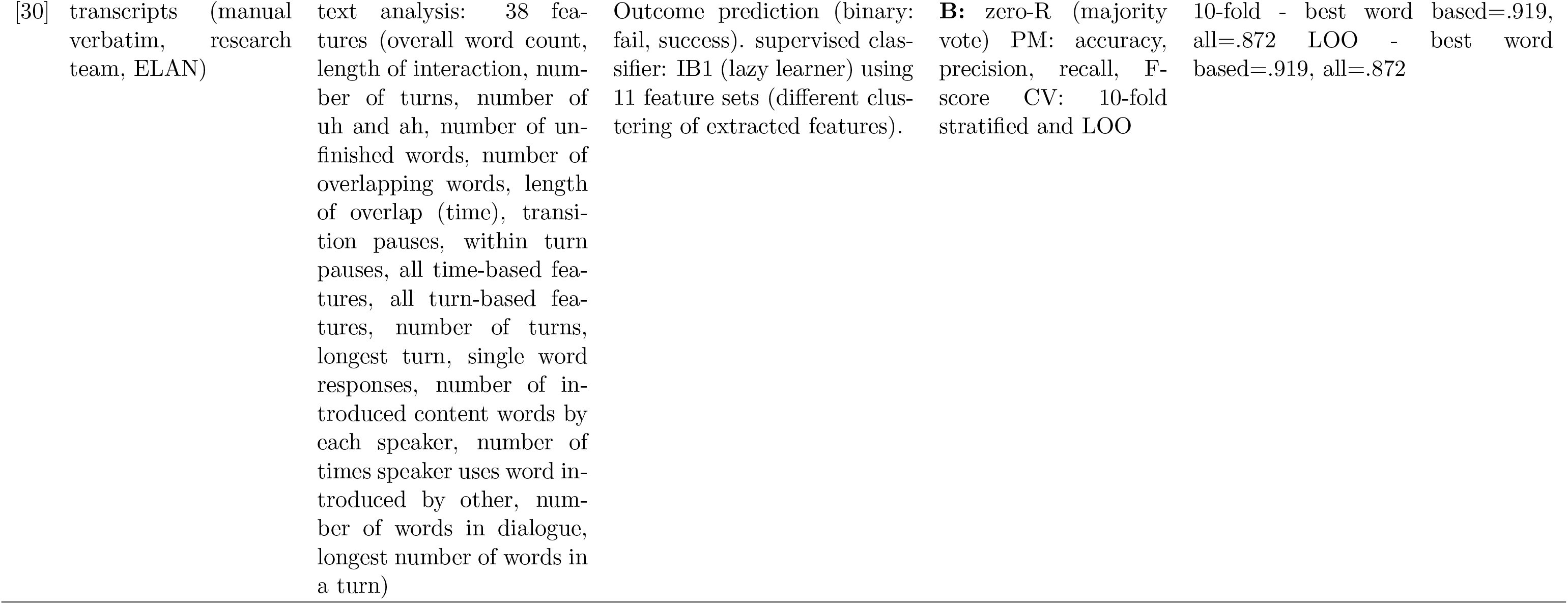
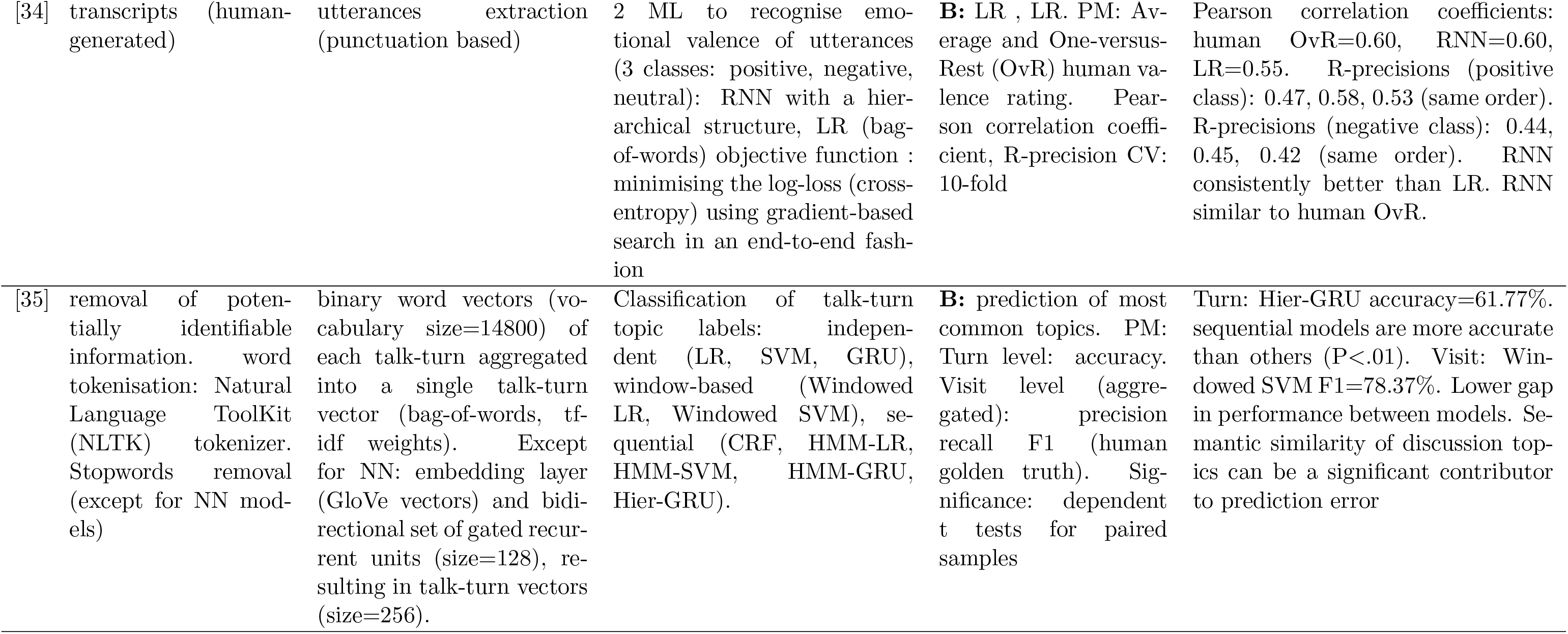
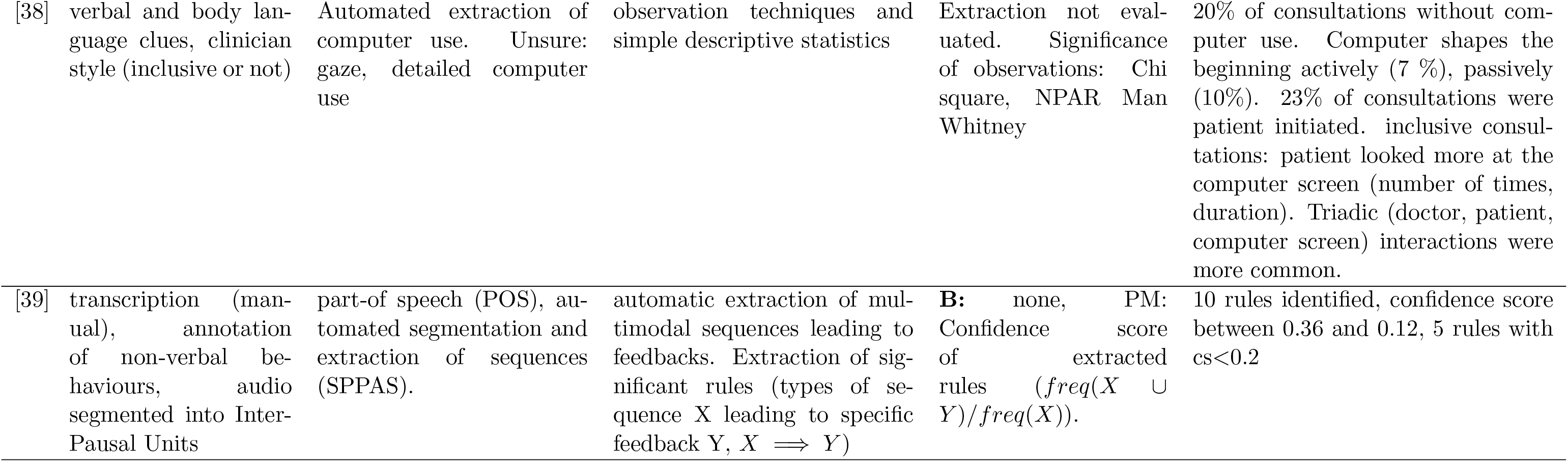
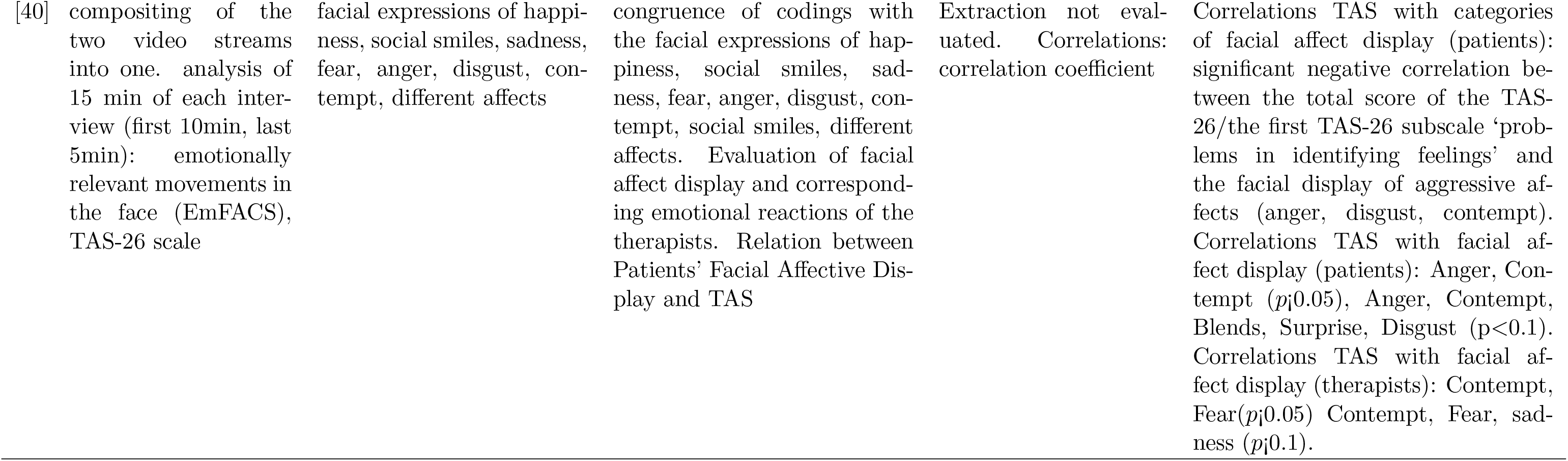
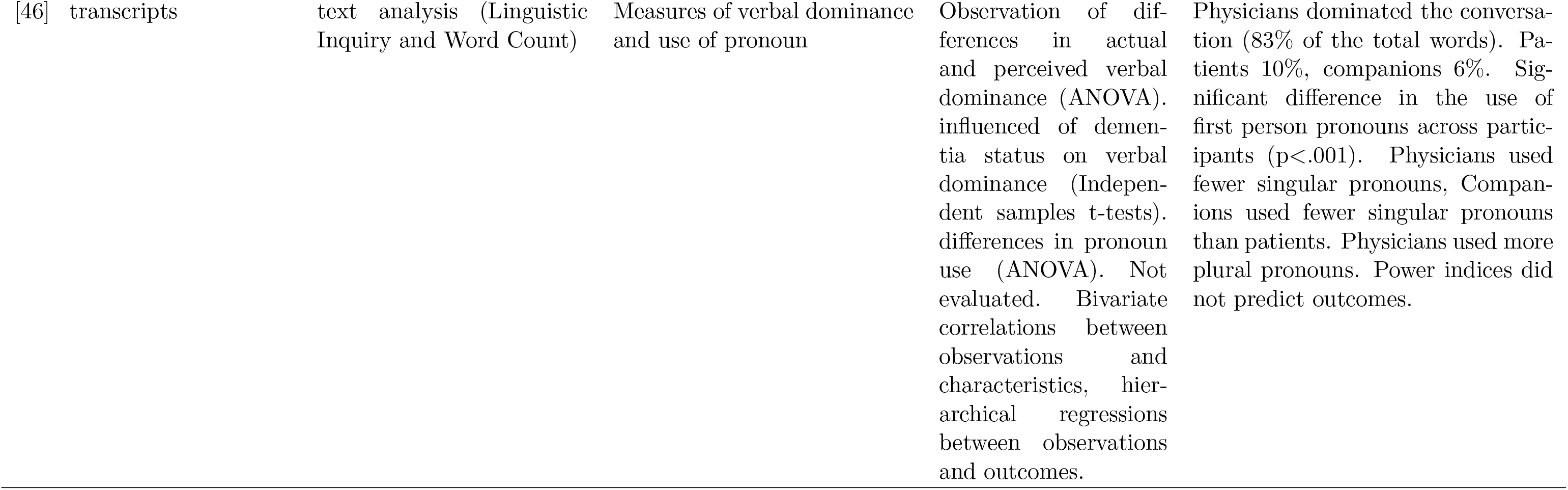
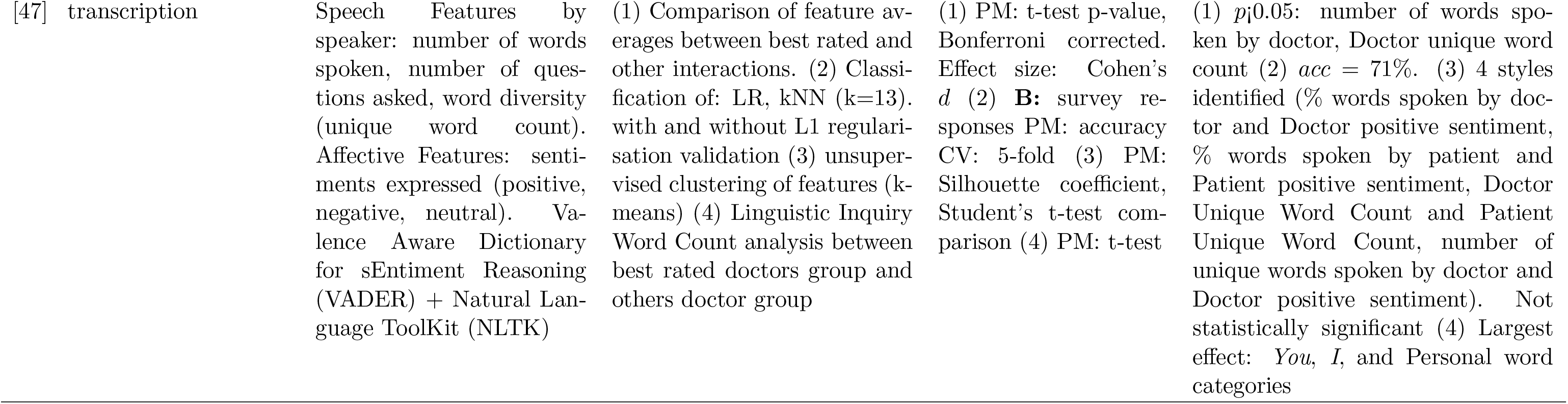
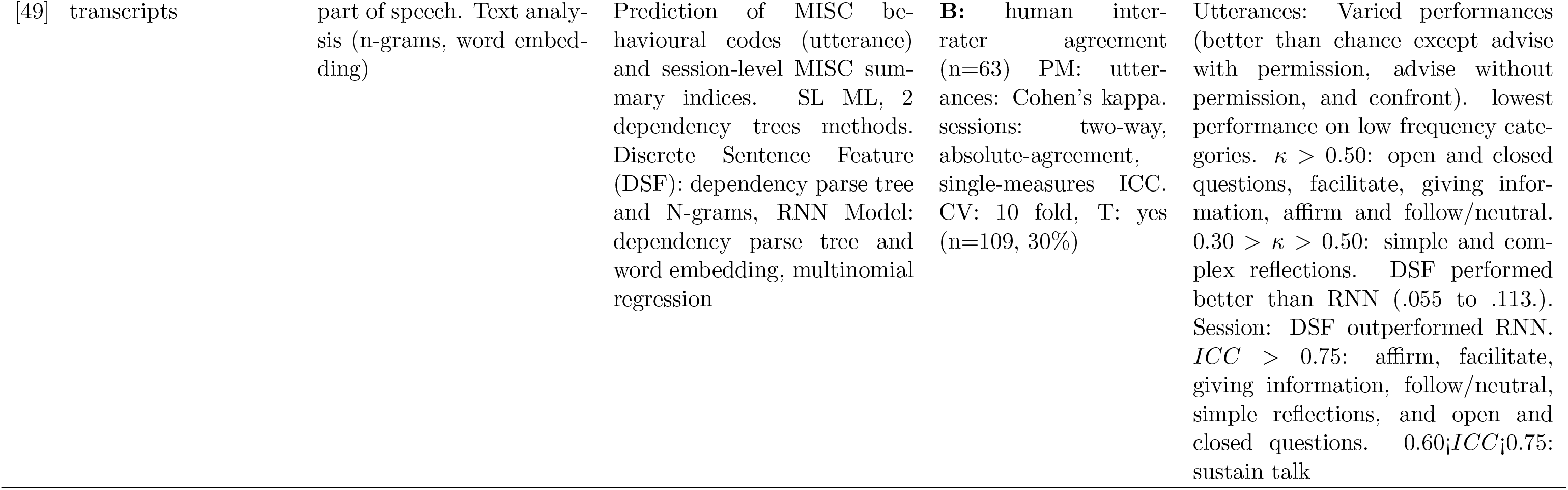
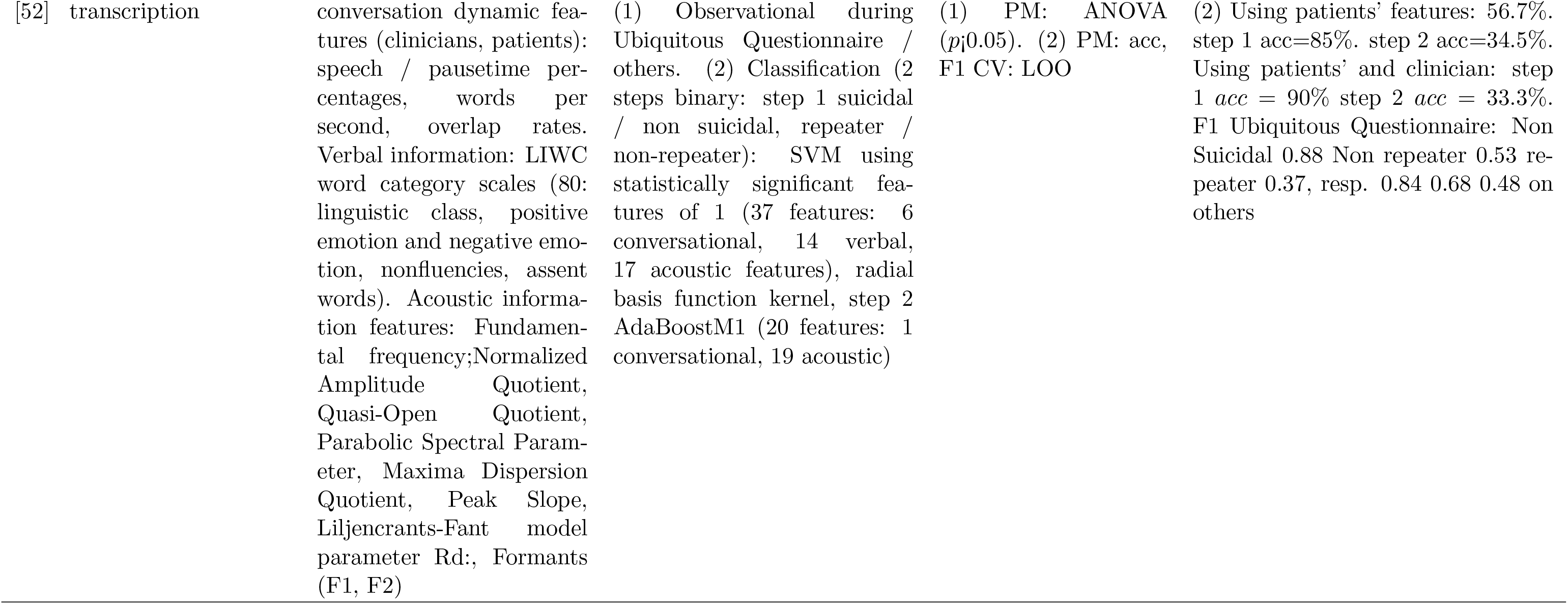
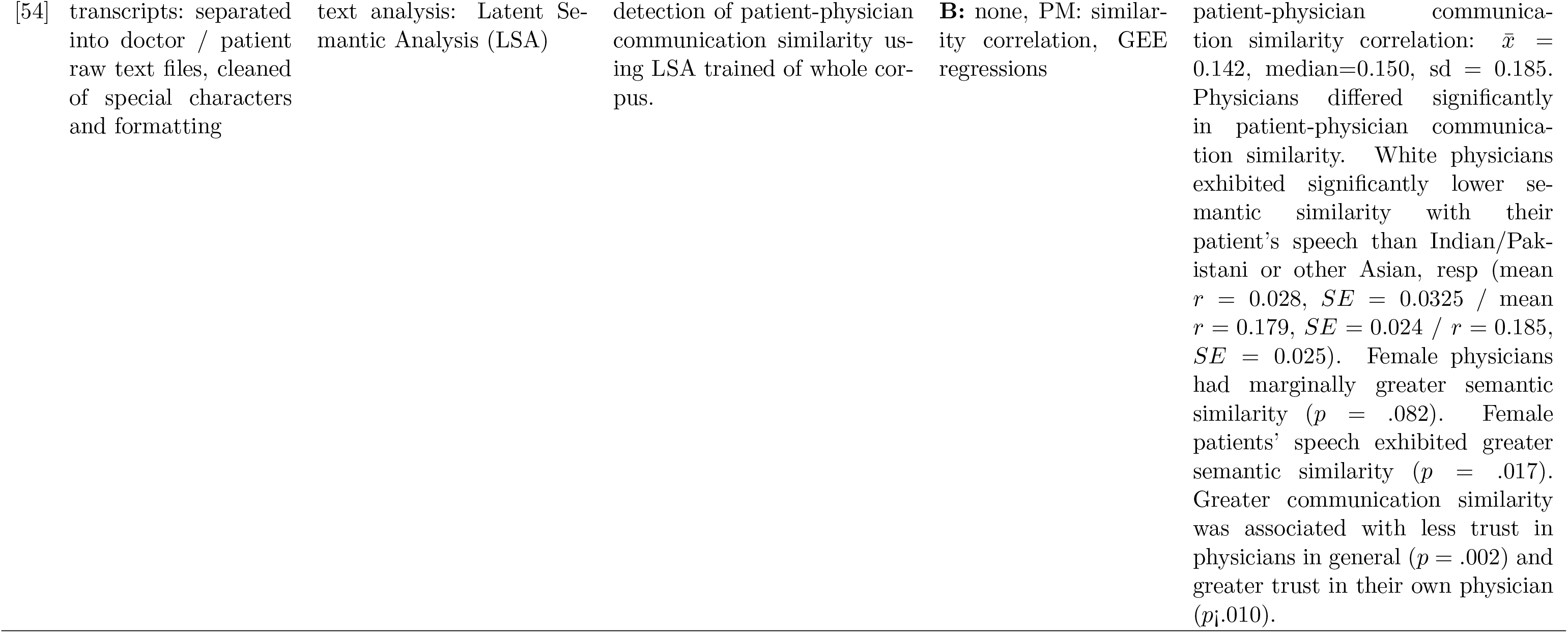
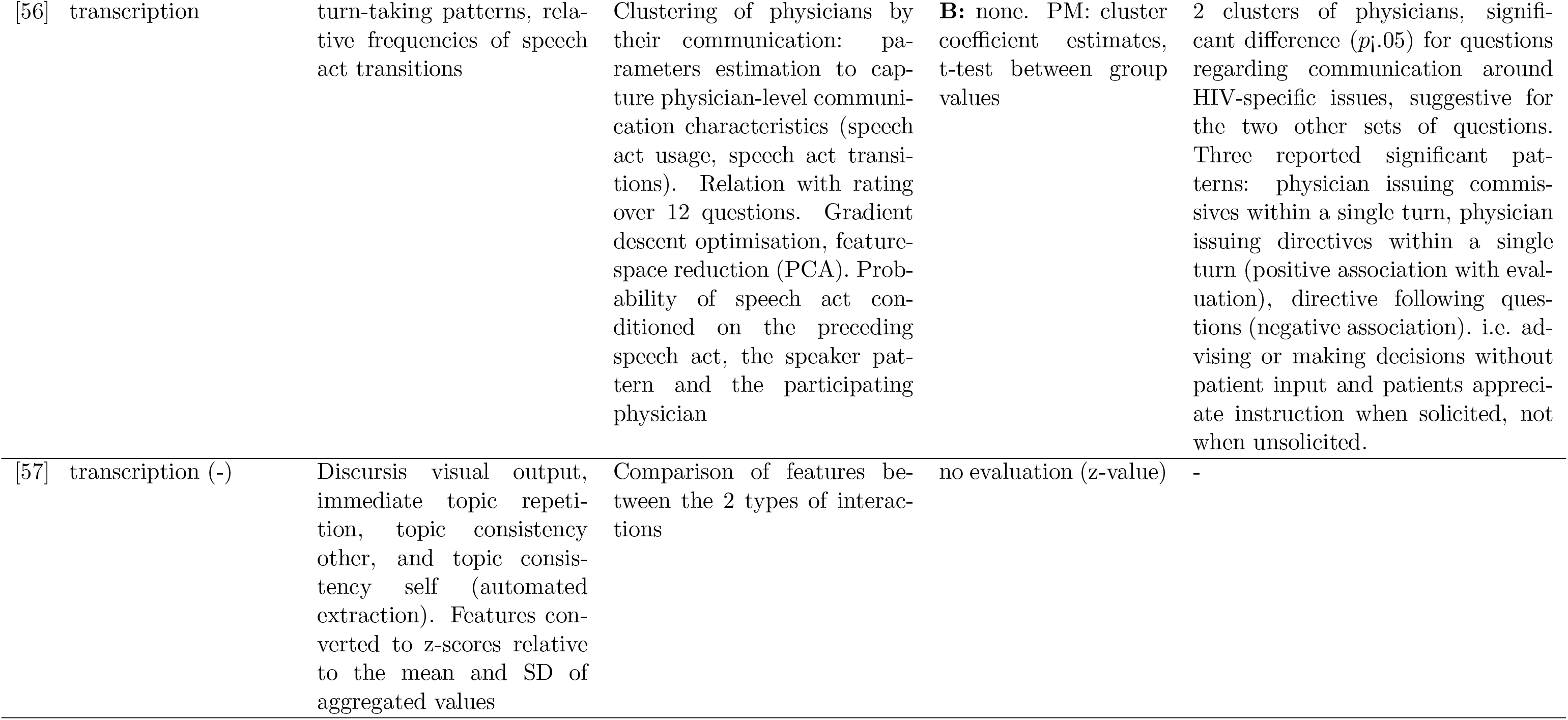
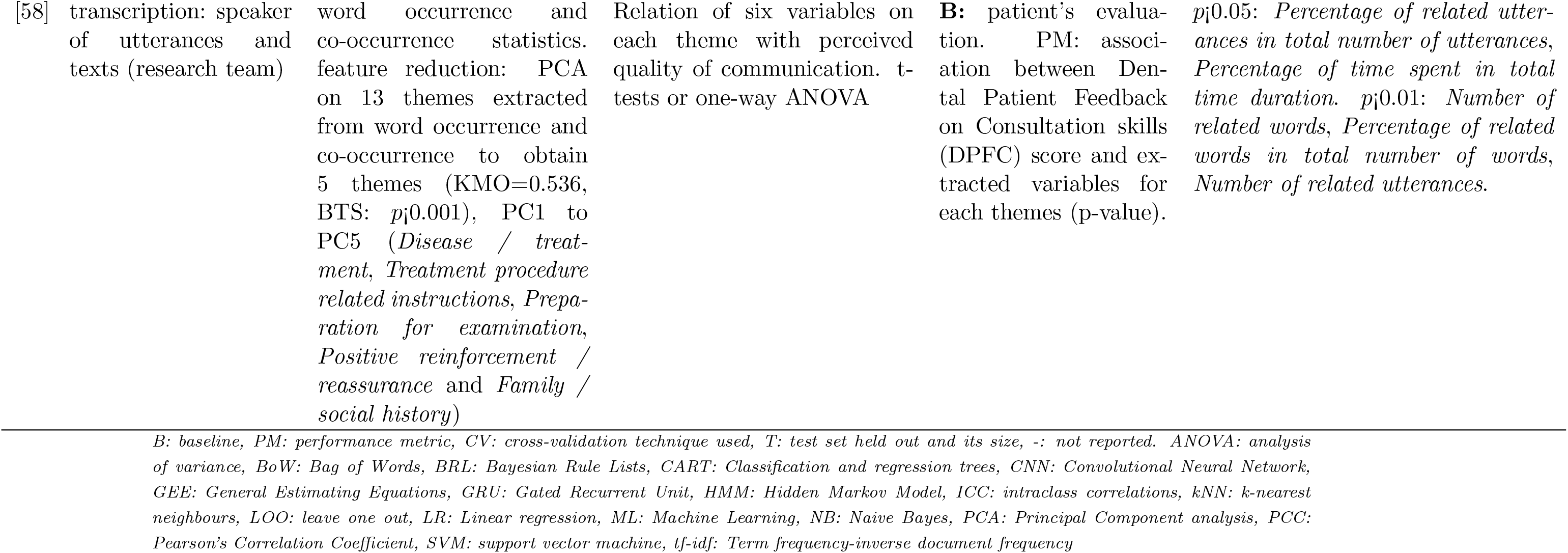
Data analysis.

**Table A.10:**
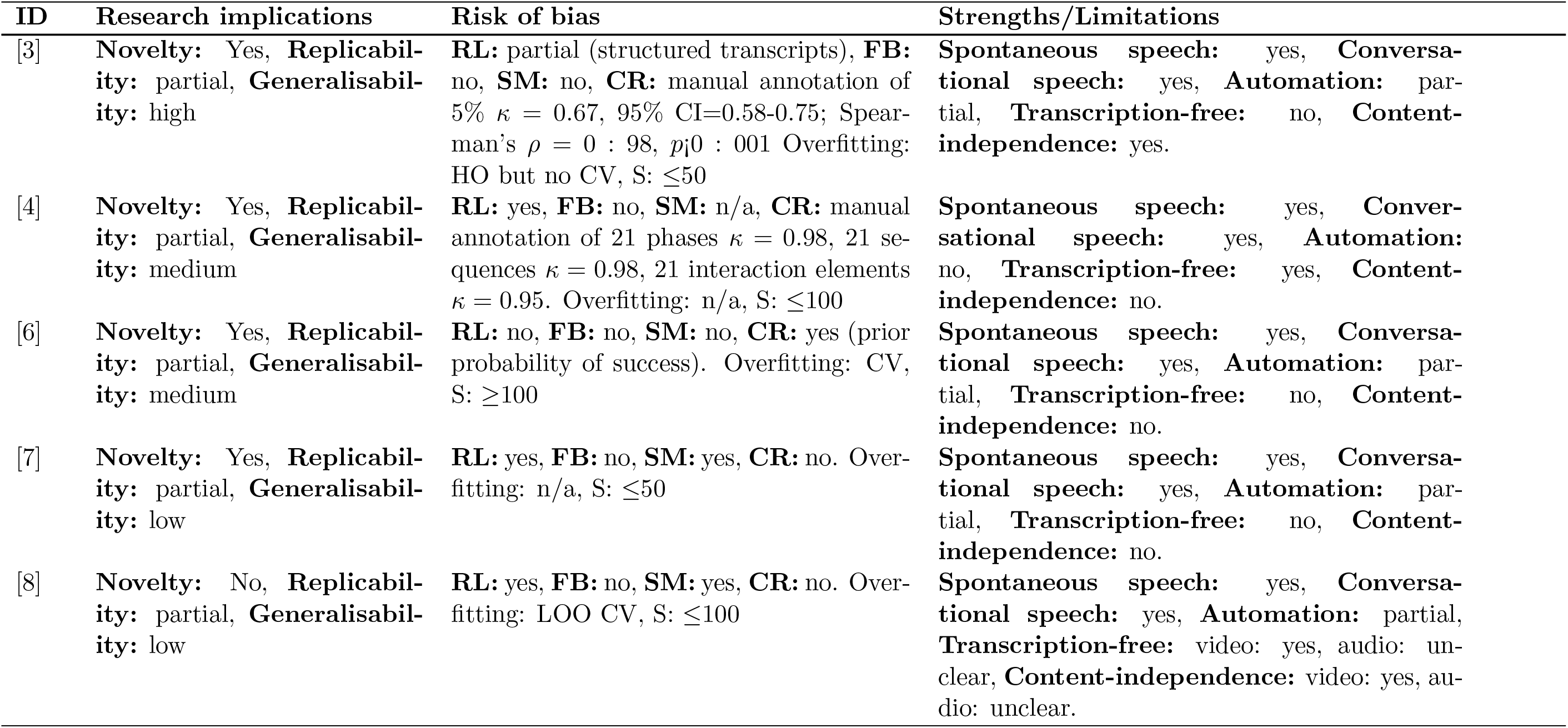
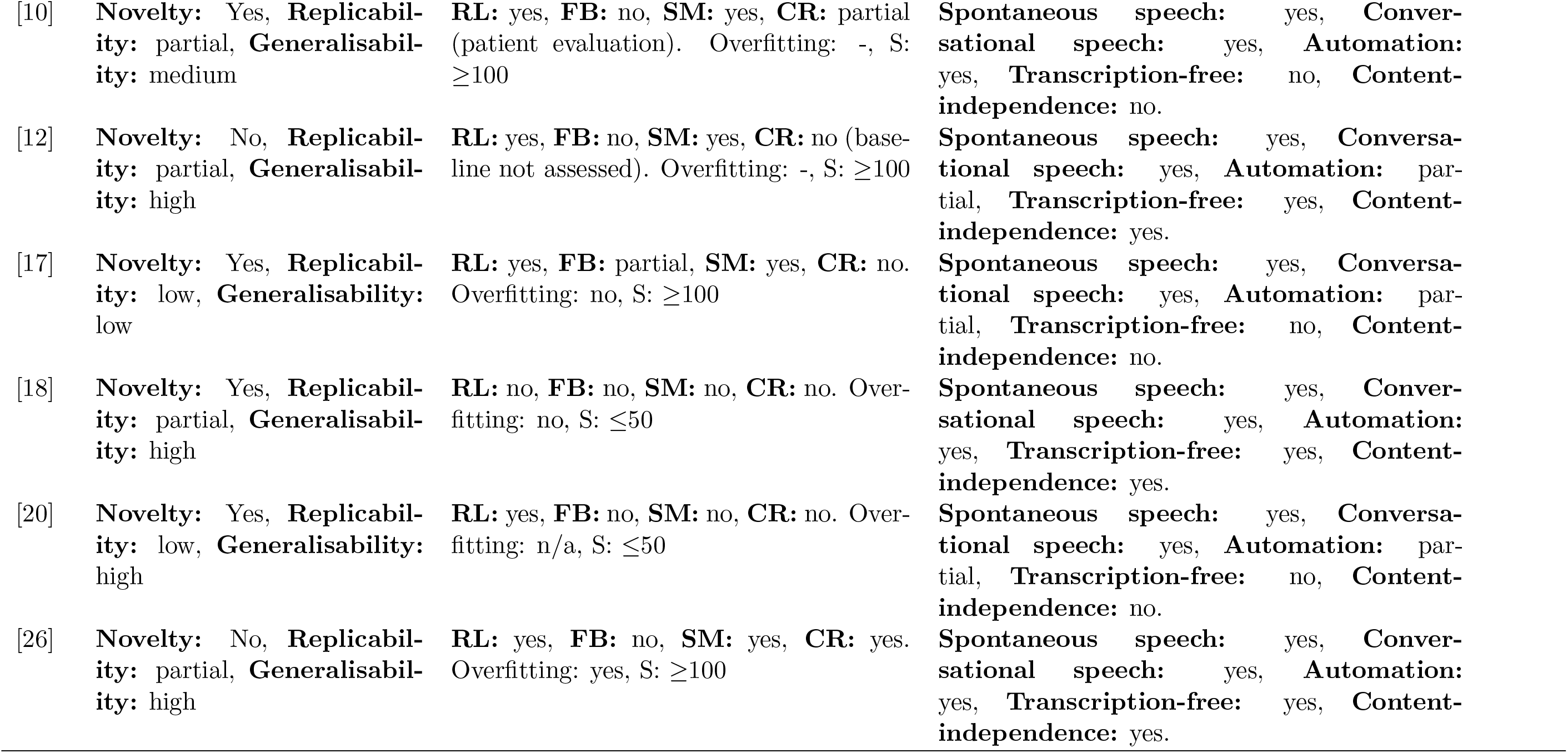
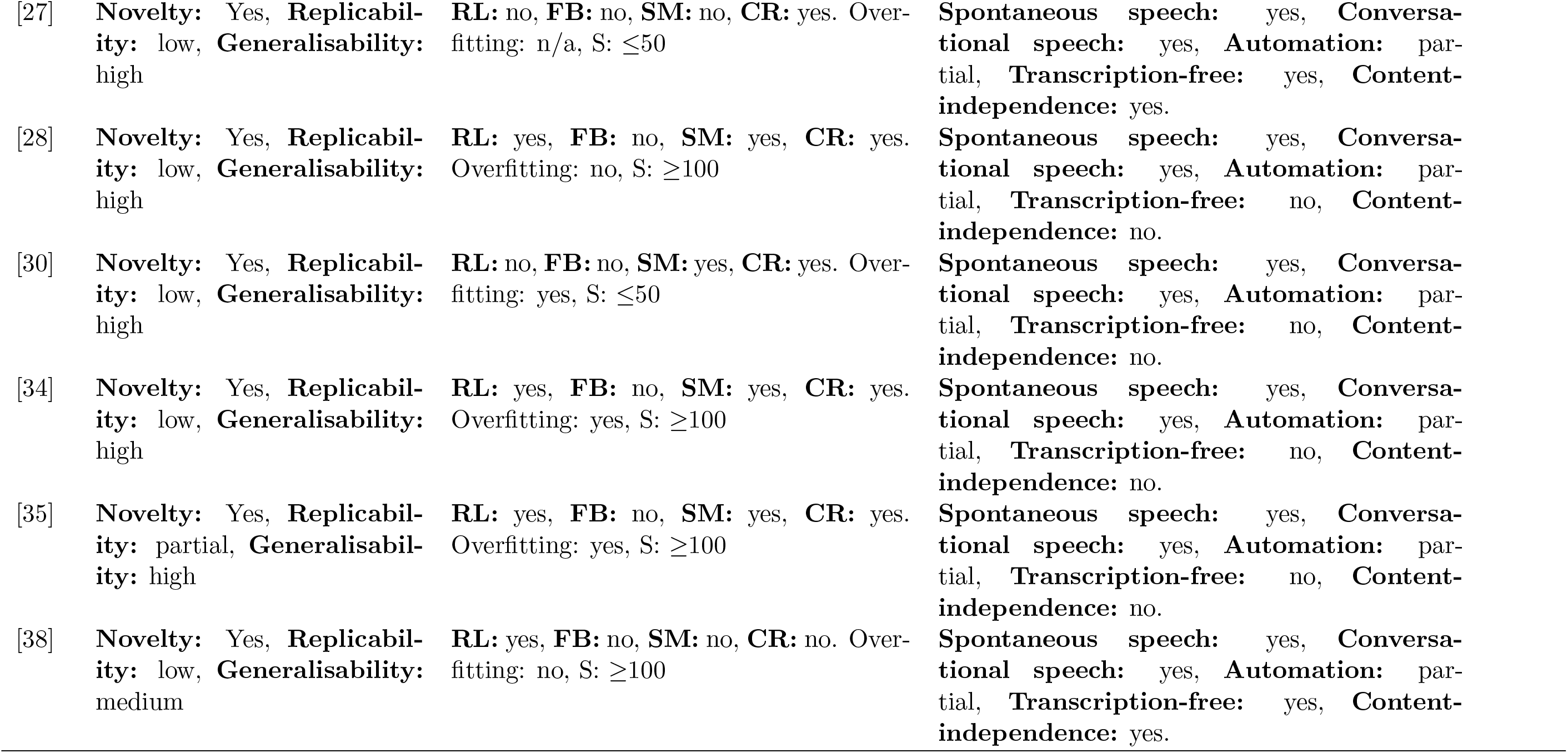
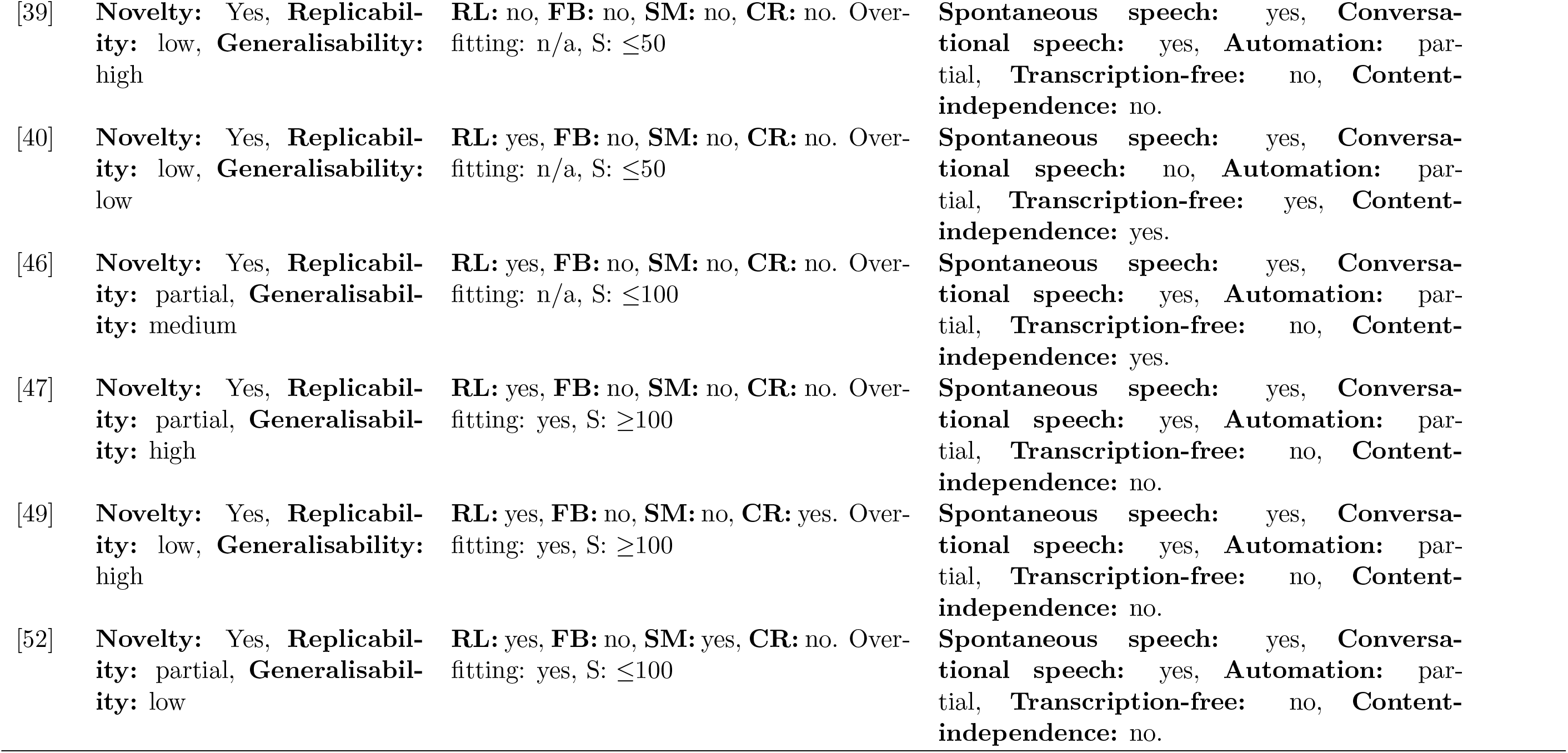
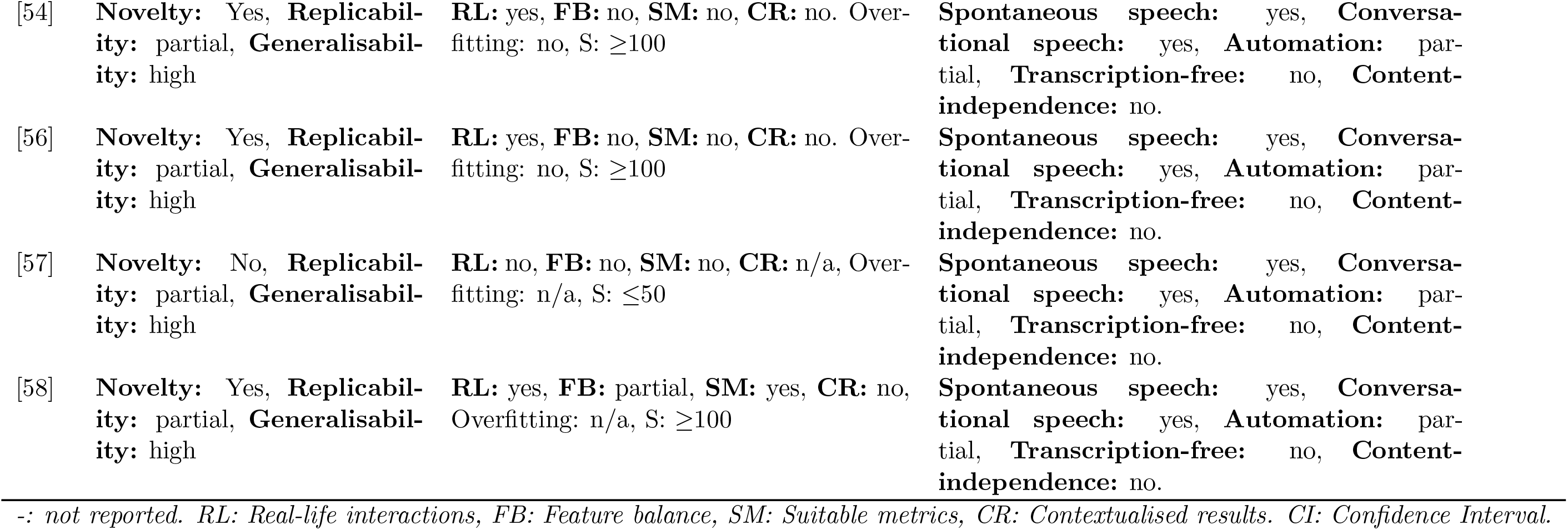
Study assessment.

| ID | Research implications | Risk of bias | Strengths/Limitations |
| --- | --- | --- | --- |
| [3] | **Novelty:** Yes, **Replicability:** partial, **Generalisability:** high | **RL:** partial (structured transcripts), **FB:** no, **SM:** no, **CR:** manual annotation of 5% $\kappa = 0.67$, 95% CI=0.58-0.75; Spearman's $\rho = 0 : 98$, $p < 0.001$ Overfitting: HO but no CV, S: $\leq 50$ | **Spontaneous speech:** yes, **Conversational speech:** yes, **Automation:** partial, **Transcription-free:** no, **Content-independence:** yes. |
| [4] | **Novelty:** Yes, **Replicability:** partial, **Generalisability:** medium | **RL:** yes, **FB:** no, **SM:** n/a, **CR:** manual annotation of 21 phases $\kappa = 0.98$, 21 sequences $\kappa = 0.98$, 21 interaction elements $\kappa = 0.95$. Overfitting: n/a, S: $\leq 100$ | **Spontaneous speech:** yes, **Conversational speech:** yes, **Automation:** no, **Transcription-free:** yes, **Content-independence:** no. |
| [6] | **Novelty:** Yes, **Replicability:** partial, **Generalisability:** medium | **RL:** no, **FB:** no, **SM:** no, **CR:** yes (prior probability of success). Overfitting: CV, S: $\geq 100$ | **Spontaneous speech:** yes, **Conversational speech:** yes, **Automation:** partial, **Transcription-free:** no, **Content-independence:** no. |
| [7] | **Novelty:** Yes, **Replicability:** partial, **Generalisability:** low | **RL:** yes, **FB:** no, **SM:** yes, **CR:** no. Overfitting: n/a, S: $\leq 50$ | **Spontaneous speech:** yes, **Conversational speech:** yes, **Automation:** partial, **Transcription-free:** no, **Content-independence:** no. |
| [8] | **Novelty:** No, **Replicability:** partial, **Generalisability:** low | **RL:** yes, **FB:** no, **SM:** yes, **CR:** no. Overfitting: LOO CV, S: $\leq 100$ | **Spontaneous speech:** yes, **Conversational speech:** yes, **Automation:** partial, **Transcription-free:** video: yes, audio: unclear, **Content-independence:** video: yes, audio: unclear. |
| [10] | **Novelty:** Yes, **Replicability:** partial, **Generalisability:** medium | **RL:** yes, **FB:** no, **SM:** yes, **CR:** partial (patient evaluation). Overfitting: -, S: $\geq 100$ | **Spontaneous speech:** yes, **Conversational speech:** yes, **Automation:** yes, **Transcription-free:** no, **Content-independence:** no. |
| [12] | **Novelty:** No, **Replicability:** partial, **Generalisability:** high | **RL:** yes, **FB:** no, **SM:** yes, **CR:** no (baseline not assessed). Overfitting: -, S: $\geq 100$ | **Spontaneous speech:** yes, **Conversational speech:** yes, **Automation:** partial, **Transcription-free:** yes, **Content-independence:** yes. |
| [17] | **Novelty:** Yes, **Replicability:** low, **Generalisability:** low | **RL:** yes, **FB:** partial, **SM:** yes, **CR:** no. Overfitting: no, S: $\geq 100$ | **Spontaneous speech:** yes, **Conversational speech:** yes, **Automation:** partial, **Transcription-free:** no, **Content-independence:** no. |
| [18] | **Novelty:** Yes, **Replicability:** partial, **Generalisability:** high | **RL:** no, **FB:** no, **SM:** no, **CR:** no. Overfitting: no, S: $\leq 50$ | **Spontaneous speech:** yes, **Conversational speech:** yes, **Automation:** yes, **Transcription-free:** yes, **Content-independence:** yes. |
| [20] | **Novelty:** Yes, **Replicability:** low, **Generalisability:** high | **RL:** yes, **FB:** no, **SM:** no, **CR:** no. Overfitting: n/a, S: $\leq 50$ | **Spontaneous speech:** yes, **Conversational speech:** yes, **Automation:** partial, **Transcription-free:** no, **Content-independence:** no. |
| [26] | **Novelty:** No, **Replicability:** partial, **Generalisability:** high | **RL:** yes, **FB:** no, **SM:** yes, **CR:** yes. Overfitting: yes, S: $\geq 100$ | **Spontaneous speech:** yes, **Conversational speech:** yes, **Automation:** yes, **Transcription-free:** yes, **Content-independence:** yes. |
| [27] | **Novelty:** Yes, **Replicability:** low, **Generalisability:** high | **RL:** no, **FB:** no, **SM:** no, **CR:** yes. Overfitting: n/a, S: $\leq 50$ | **Spontaneous speech:** yes, **Conversational speech:** yes, **Automation:** partial, **Transcription-free:** yes, **Content-independence:** yes. |
| [28] | **Novelty:** Yes, **Replicability:** low, **Generalisability:** high | **RL:** yes, **FB:** no, **SM:** yes, **CR:** yes. Overfitting: no, S: $\geq 100$ | **Spontaneous speech:** yes, **Conversational speech:** yes, **Automation:** partial, **Transcription-free:** no, **Content-independence:** no. |
| [30] | **Novelty:** Yes, **Replicability:** low, **Generalisability:** high | **RL:** no, **FB:** no, **SM:** yes, **CR:** yes. Overfitting: yes, S: $\leq 50$ | **Spontaneous speech:** yes, **Conversational speech:** yes, **Automation:** partial, **Transcription-free:** no, **Content-independence:** no. |
| [34] | **Novelty:** Yes, **Replicability:** low, **Generalisability:** high | **RL:** yes, **FB:** no, **SM:** yes, **CR:** yes. Overfitting: yes, S: $\geq 100$ | **Spontaneous speech:** yes, **Conversational speech:** yes, **Automation:** partial, **Transcription-free:** no, **Content-independence:** no. |
| [35] | **Novelty:** Yes, **Replicability:** partial, **Generalisability:** high | **RL:** yes, **FB:** no, **SM:** yes, **CR:** yes. Overfitting: yes, S: $\geq 100$ | **Spontaneous speech:** yes, **Conversational speech:** yes, **Automation:** partial, **Transcription-free:** no, **Content-independence:** no. |
| [38] | **Novelty:** Yes, **Replicability:** low, **Generalisability:** medium | **RL:** yes, **FB:** no, **SM:** no, **CR:** no. Overfitting: no, S: $\geq 100$ | **Spontaneous speech:** yes, **Conversational speech:** yes, **Automation:** partial, **Transcription-free:** yes, **Content-independence:** yes. |
| [39] | **Novelty:** Yes, **Replicability:** low, **Generalisability:** high | **RL:** no, **FB:** no, **SM:** no, **CR:** no. Overfitting: n/a, S: $\leq 50$ | **Spontaneous speech:** yes, **Conversational speech:** yes, **Automation:** partial, **Transcription-free:** no, **Content-independence:** no. |
| [40] | **Novelty:** Yes, **Replicability:** low, **Generalisability:** low | **RL:** yes, **FB:** no, **SM:** no, **CR:** no. Overfitting: n/a, S: $\leq 50$ | **Spontaneous speech:** yes, **Conversational speech:** no, **Automation:** partial, **Transcription-free:** yes, **Content-independence:** yes. |
| [46] | **Novelty:** Yes, **Replicability:** partial, **Generalisability:** medium | **RL:** yes, **FB:** no, **SM:** no, **CR:** no. Overfitting: n/a, S: $\leq 100$ | **Spontaneous speech:** yes, **Conversational speech:** yes, **Automation:** partial, **Transcription-free:** no, **Content-independence:** yes. |
| [47] | **Novelty:** Yes, **Replicability:** partial, **Generalisability:** high | **RL:** yes, **FB:** no, **SM:** no, **CR:** no. Overfitting: yes, S: $\geq 100$ | **Spontaneous speech:** yes, **Conversational speech:** yes, **Automation:** partial, **Transcription-free:** no, **Content-independence:** no. |
| [49] | **Novelty:** Yes, **Replicability:** low, **Generalisability:** high | **RL:** yes, **FB:** no, **SM:** no, **CR:** yes. Overfitting: yes, S: $\geq 100$ | **Spontaneous speech:** yes, **Conversational speech:** yes, **Automation:** partial, **Transcription-free:** no, **Content-independence:** no. |
| [52] | **Novelty:** Yes, **Replicability:** partial, **Generalisability:** low | **RL:** yes, **FB:** no, **SM:** yes, **CR:** no. Overfitting: yes, S: $\leq 100$ | **Spontaneous speech:** yes, **Conversational speech:** yes, **Automation:** partial, **Transcription-free:** no, **Content-independence:** no. |
| [54] | **Novelty:** Yes, **Replicability:** partial, **Generalisability:** high | **RL:** yes, **FB:** no, **SM:** no, **CR:** no. Overfitting: no, S: $\geq 100$ | **Spontaneous speech:** yes, **Conversational speech:** yes, **Automation:** partial, **Transcription-free:** no, **Content-independence:** no. |
| [56] | **Novelty:** Yes, **Replicability:** partial, **Generalisability:** high | **RL:** yes, **FB:** no, **SM:** no, **CR:** no. Overfitting: no, S: $\geq 100$ | **Spontaneous speech:** yes, **Conversational speech:** yes, **Automation:** partial, **Transcription-free:** no, **Content-independence:** no. |
| [57] | **Novelty:** No, **Replicability:** partial, **Generalisability:** high | **RL:** no, **FB:** no, **SM:** no, **CR:** n/a, Overfitting: n/a, S: $\leq 50$ | **Spontaneous speech:** yes, **Conversational speech:** yes, **Automation:** partial, **Transcription-free:** no, **Content-independence:** no. |
| [58] | **Novelty:** Yes, **Replicability:** partial, **Generalisability:** high | **RL:** yes, **FB:** partial, **SM:** yes, **CR:** no, Overfitting: n/a, S: $\geq 100$ | **Spontaneous speech:** yes, **Conversational speech:** yes, **Automation:** partial, **Transcription-free:** no, **Content-independence:** no. |

*-: not reported. RL: Real-life interactions, FB: Feature balance, SM: Suitable metrics, CR: Contextualised results. CI: Confidence Interval.*

**Table A.11:**
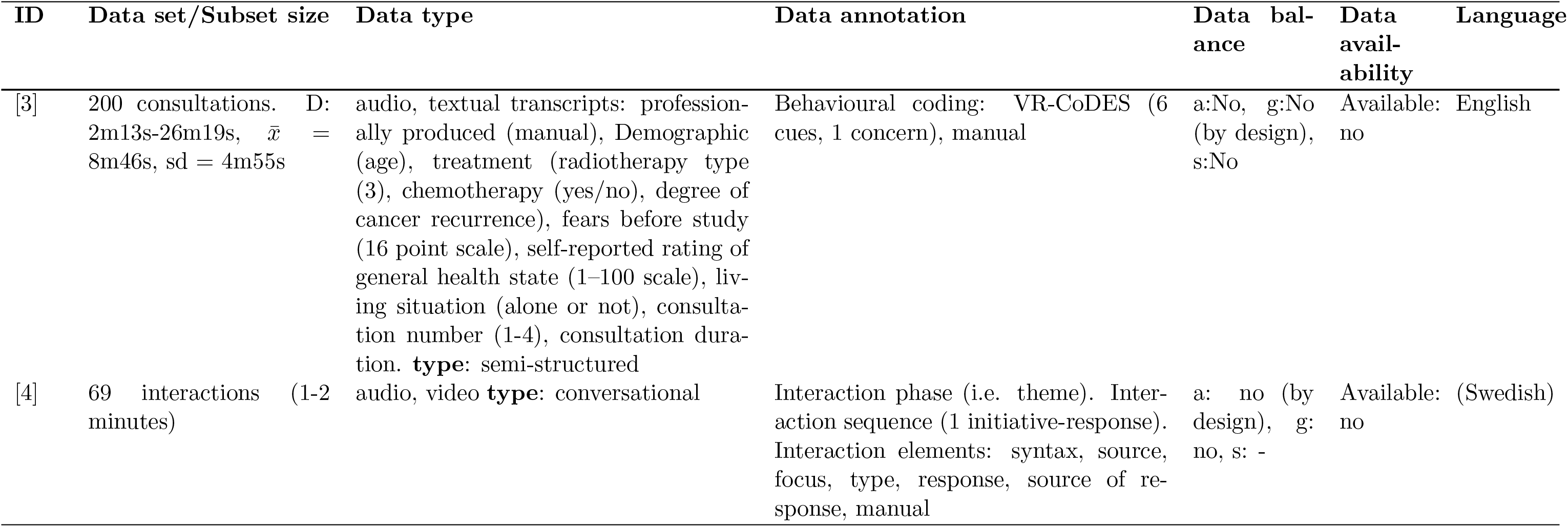
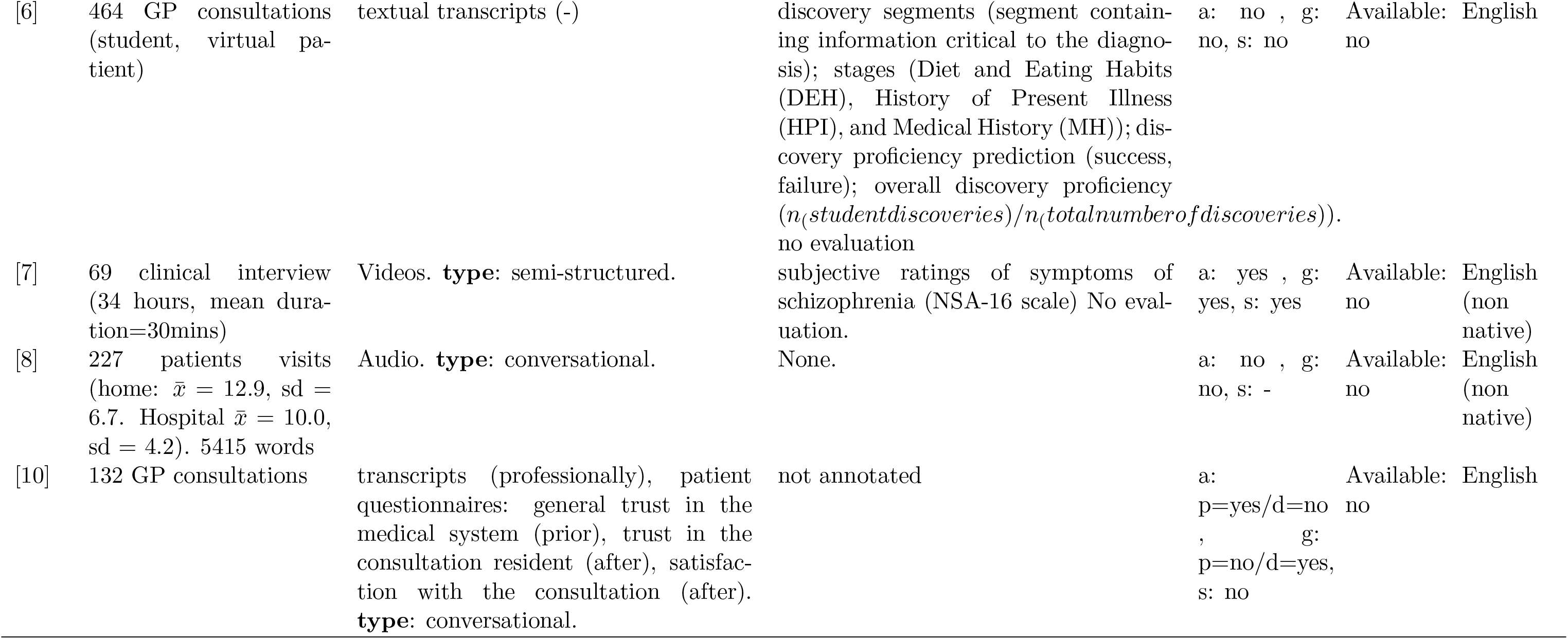
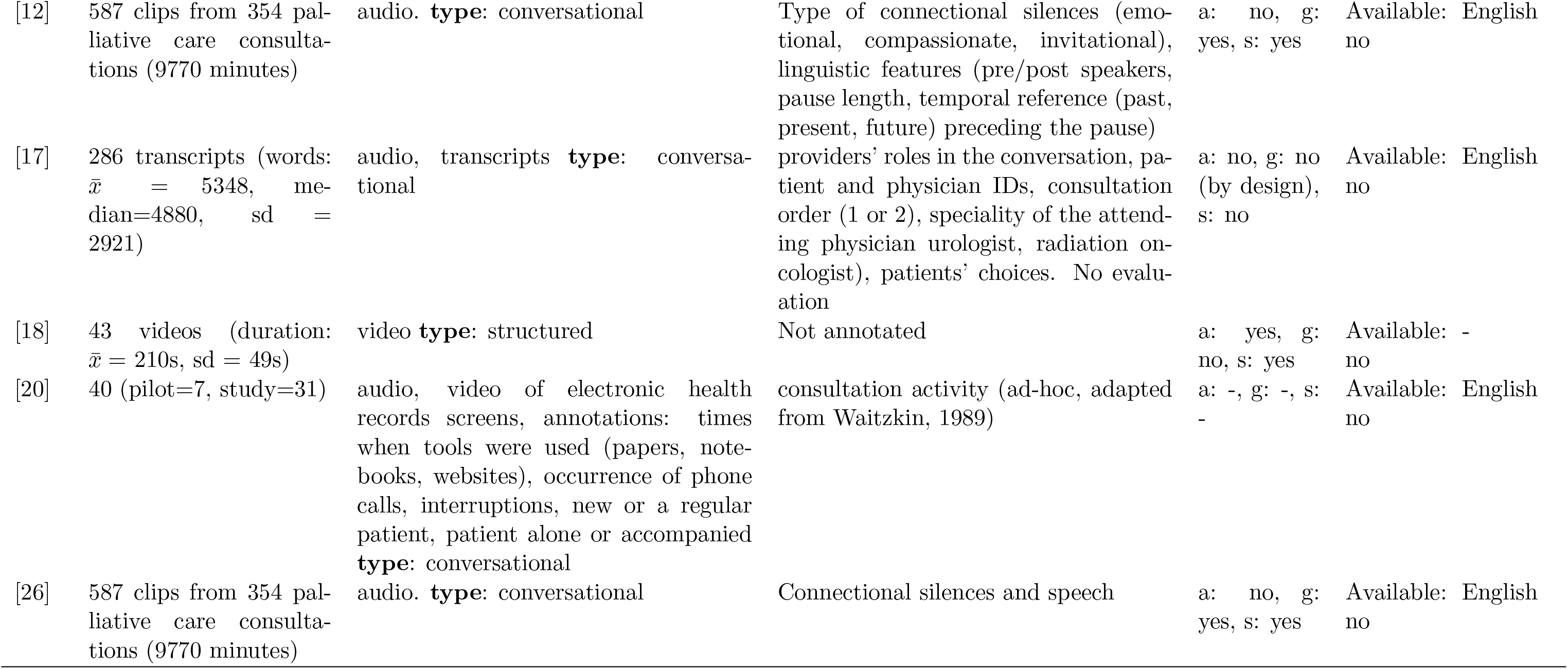
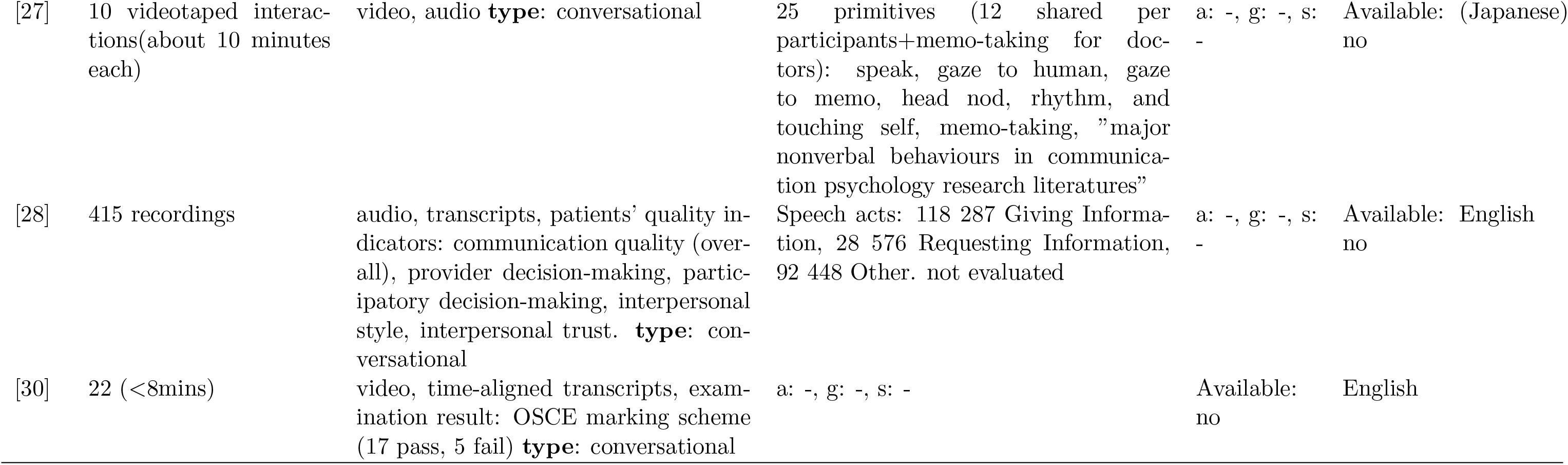
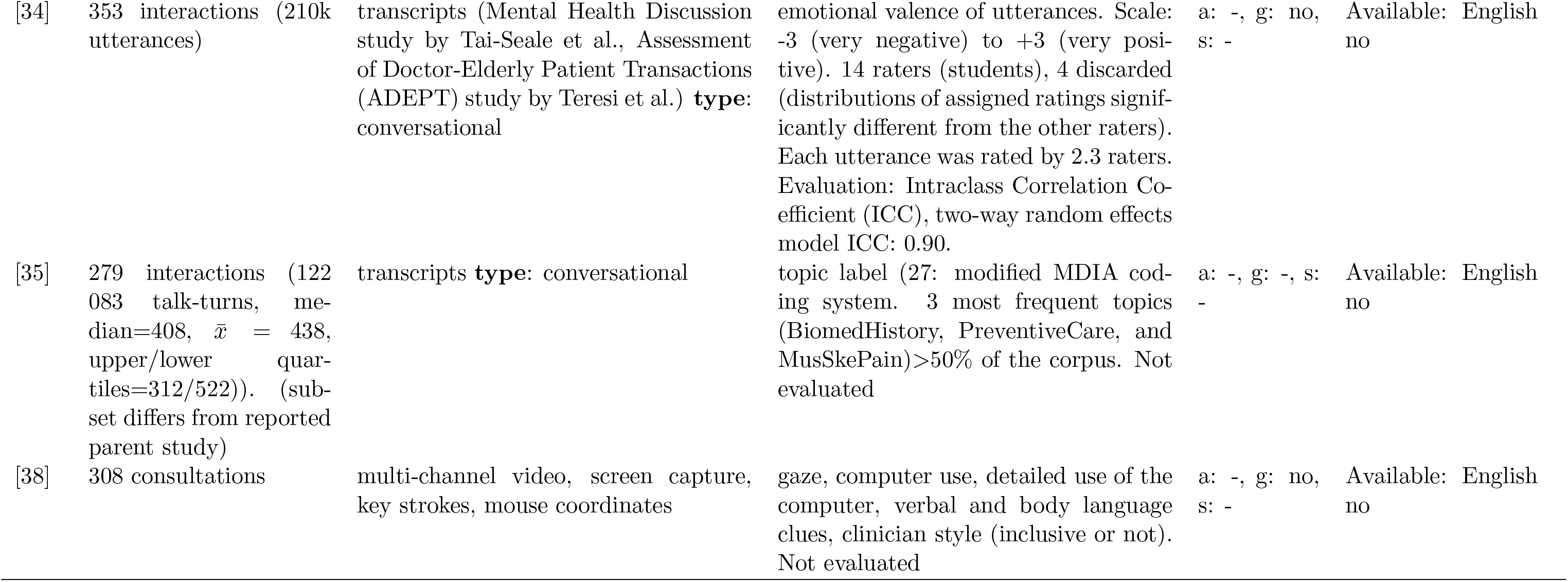
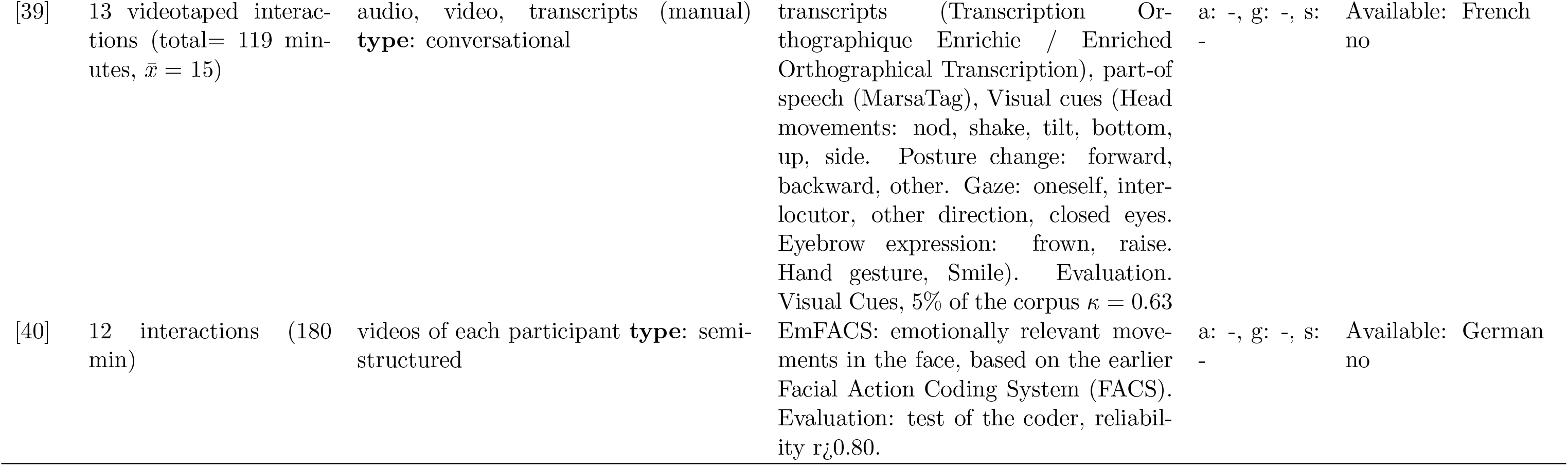
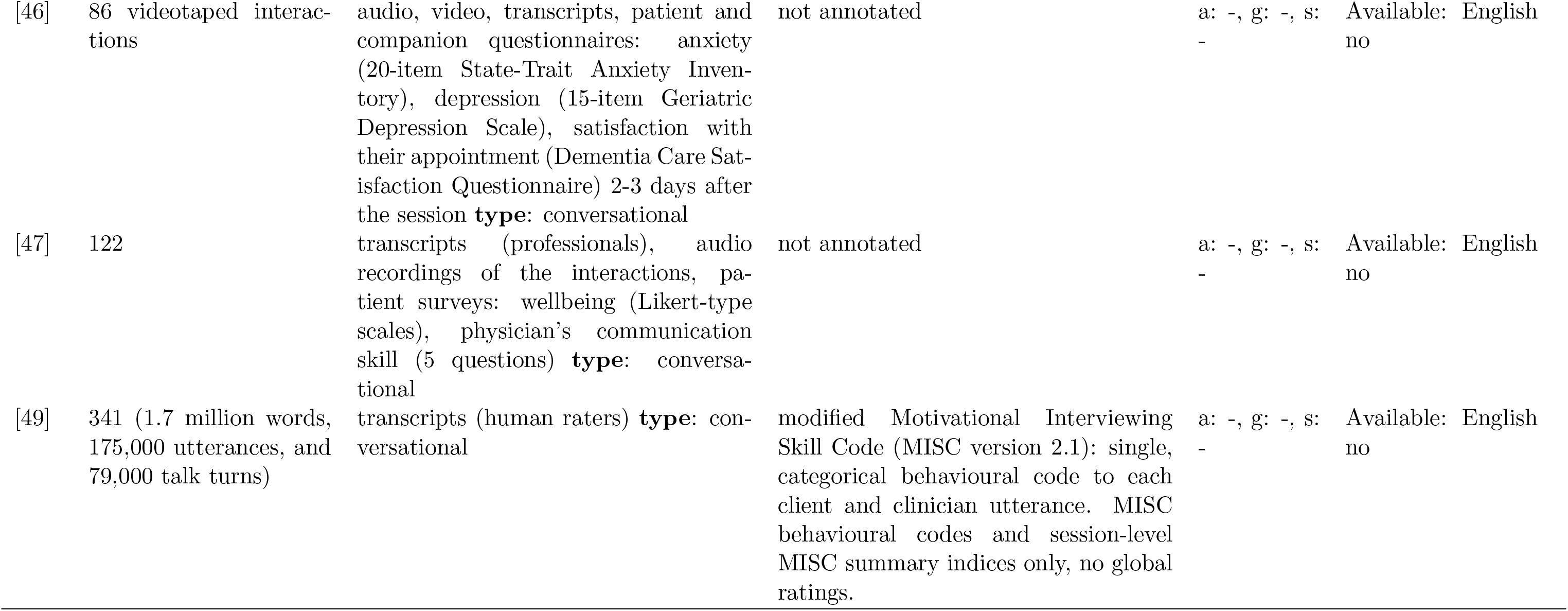
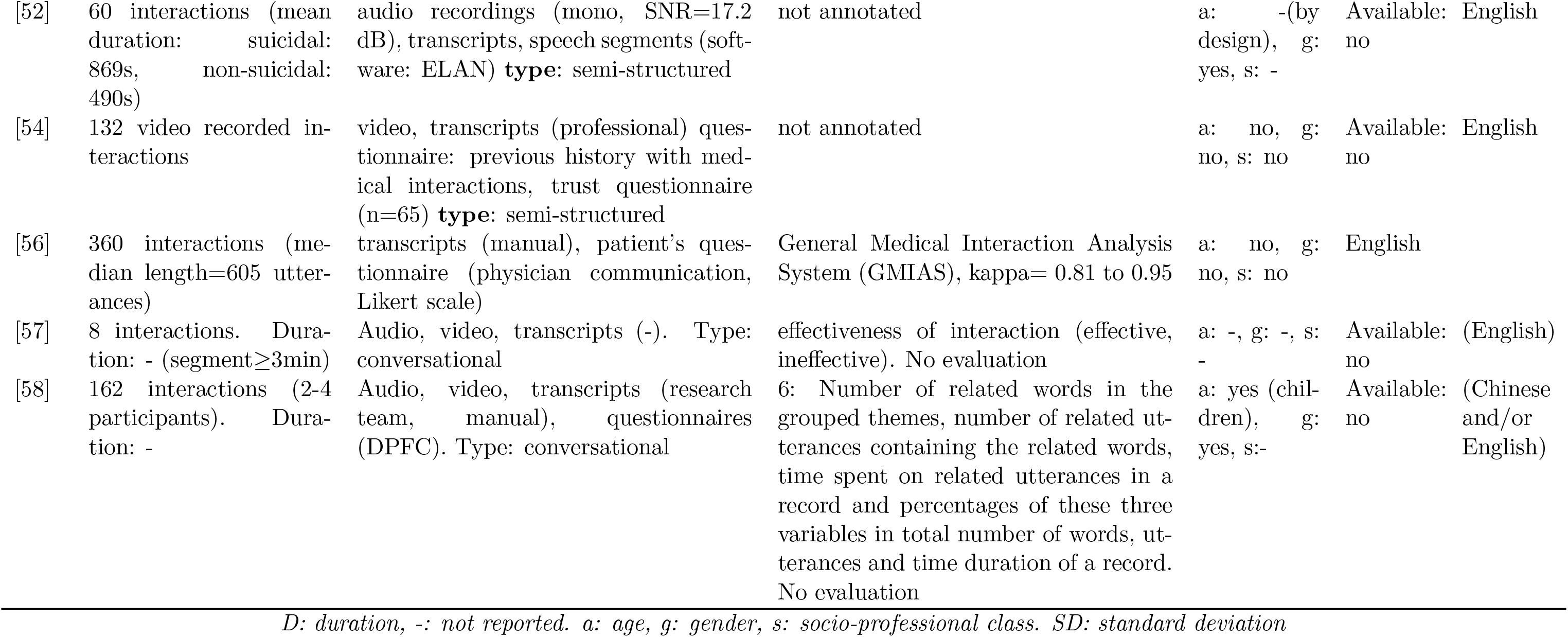
Datasets assessment.

## Footnotes

1 Google Scholar is a specialised search engine for published scientific literature. It is a valuable resource to lookup specific references, e.g. cited articles in a publication.

2 DuckDuckGo is a generic web search engine with a strong focus on keeping user’s privacy. It is an alternative to the more popular web search engine by Google.

3 https://edin.ac/3OIvxVW

